# Clinical Utility of Automatable Prediction Models for Improving Palliative and End-Of-Life Care Outcomes: Towards Routine Decision Analysis Before Implementation

**DOI:** 10.1101/2021.03.27.21254465

**Authors:** Ryeyan Taseen, Jean-François Ethier

## Abstract

**Objective:** To evaluate the clinical utility of automatable prediction models for increasing goals-of-care discussions among hospitalized patients at the end-of-life.

**Materials and Methods:** We developed three Random Forest models and updated the Modified Hospital One-year Mortality Risk model: alternative models to predict one-year mortality (proxy for EOL status) using admission-time data. Admissions from July 2011-2016 were used for training and those from July 2017-2018 were used for temporal validation. We simulated alerts for admissions in the validation cohort and modelled alternative scenarios where alerts lead to code status orders (CSOs) in the EHR. We linked actual CSOs and calculated the expected risk difference (eRD), the number needed to benefit (NNB) and the net benefit (NB) of each model for the patient-centered outcome of a CSO among EOL hospitalizations.

**Results:** Models had a C-statistic of 0.79-0.86 among unique patients. A CSO was documented during 2599 of 3773 hospitalizations at the EOL (68.9%). At a threshold that identified 10% of eligible admissions, the eRD ranged from 5.4% to 10.7% (NNB 5.4-10.9 alerts). Under usual care, a CSO had a 34% PPV for EOL status. Using this to inform the relative cost of FPs, only two models improved NB over usual care. A RF model with diagnostic predictors had the highest clinical utility by either measure, including in sensitivity analyses.

**Discussion:** Automatable prediction models with acceptable temporal validity differed meaningfully in their expected ability to improve patient-centered outcomes over usual care.

**Conclusion:** Decision-analysis should precede implementation of automated prediction models for improving palliative and EOL care outcomes.

## INTRODUCTION

End-of-life (EOL) conversations and shared decision-making between clinical staff and hospitalized patients can improve the quality of EOL care.[1,2] In the hospital setting, these conversations inform goals-of-care (GOC) documentation, particularly code status orders (CSOs), which encode the essential preferences for life-supporting therapy.[3] Hospitalizations are frequent at the EOL, and where the need to plan for future care is matched by the opportunity.[4] However, hospitalized patients with a poor prognosis do not benefit from EOL conversations or GOC documentation as often as they should.[5] Closing this gap is challenged by workload constraints and difficulty in prognostication.[6]

Clinical decision support systems (CDSS) that integrate automated prediction models may help increase the prevalence of GOC discussions by generating computerized alerts for patients with a high risk of mortality.[2,7] The rationale is that physicians, when explicitly alerted to the poor prognosis of a patient in their care, will initiate a discussion about GOC if one is appropriate and has not already occurred.

In the translational pathway of prediction models, an increasingly recognized step is the assessment of clinical utility,[8] which should occur before a prospective evaluation of clinical impact.[9,10] CDSSs, particularly those with machine learning (ML) models, are potentially costly to implement[11,12] and their impact highly subject to local factors,[13] giving reason to assess clinical utility before investing in application. Decision-analytic methods for this assessment using observational data are accessible[10,14–16] but are rarely unused for prediction models prompting palliative and EOL care (PEOLC) interventions, resulting in poor evidence of value.[17,18] A few decision-analyses in this area of research have assessed system-perspective monetary benefit;[19,20] a decision-analytic evaluation of clinical benefits and harms from the perspective of patient-centered quality improvement has remained elusive.

In this study, we evaluated the clinical utility of locally applicable prediction models using a routinely collected measure of GOC discussions, CSOs in the EHR. Our primary objective was to evaluate the clinical utility of a novel ML model compared to a published model[21] and models requiring less types of predictors. In the process, we demonstrate innovative strategies to increase the applicability of simple decision-analytic techniques for assessing the utility of automatable prediction models before implementation.

## MATERIALS AND METHODS

This retrospective study comparing prediction models includes methods for the development, validation, and decision-analytic evaluation of prediction models. We conform to the TRIPOD guidelines[22] for reporting prognostic modelling methods, and to the relevant aspects of the CHEERS guidelines[23] for reporting decision-analytic methods. The study took place at an integrated university hospital network with two sites and about 700 acute care beds in the city of Sherbrooke, Quebec, Canada (details in the supplement). IRB approval was obtained prior to data collection (IRB of the *CIUSSS de l’Estrie – CHUS* #2018-2478).

### Source of data and participants

All predictor data in the study was collected from the institutional data warehouse, which combines EHR and administrative data. All adult hospitalizations admitted to a non-psychiatric service between July 1^st^, 2011 and June 30^th^, 2018 were included in the overall cohort, except for admissions to rarely admitting specialities (e.g., genetics) or admissions with a legal context (e.g., court-ordered). Mortality records were sourced from the Quebec vital statistics registry and considered complete until June 30^th^, 2019 (additional details in supplement).

The overall cohort was split temporally, with a training cohort defined as admissions occurring between July 1^st^, 2011 and June 30^th^, 2016 inclusively, and a testing cohort defined as hospital admissions occurring between July 1^st^, 2017 and June 30^th^, 2018 inclusively. This split was designed to simulate the prospective evaluation of a given model had it been trained with all available data just before midnight on June 30^th^, 2017 and then applied prospectively for one year at our institution. Hospitalizations that occurred between July 1^st^, 2016 and June 30^th^, 2017, inclusively, were excluded to prevent any unrealistic leakage of outcome information between the training and testing cohort.

For the evaluation of clinical utility, our population of interest was all hospitalizations where there was enough time for a GOC discussion to occur and where it was not inappropriate or unnecessary given the information available to a CDSS at the point-of-care. Our selection criteria were hospitalizations with overnight stay that were not admissions in obstetrics or palliative care. The included sample for the evaluation of clinical utility were all such hospitalizations in the testing cohort.

### Prediction models

We developed a ML model using the Random Forest (RF) algorithm that includes administrative, demographic, and diagnostic predictors accessible at the time of hospital admission to predict one-year mortality (RF-AdminDemoDx, 244 predictors). As an alternative strategy, we updated the Modified Hospital One-year Mortality Risk (mHOMR) model[21] for local application (nine predictors). In addition, we specified two simplified versions of the RF-AdminDemoDx model: one where no diagnostic variables were included (RF-AdminDemo, twelve predictors) and one where only four variables – age, sex, admission service and admission type – were included (RF-Minimal). All four prediction models were feasible to operationalize with the existing informatics infrastructure, though had different requirements in terms of data access and implementation (Table 1). Data generation processes were investigated to align retrospectively extracted variables with what would be available within a few minutes of hospital admission. The models were trained with the training cohort, their temporal validity evaluated with the testing cohort and their clinical utility evaluated with the CDSS-eligible cohort. Model development, specification and validation is fully described in the supplement.

**Table 1.** Predictors included in automatable prediction models.

| Variable<sup>a</sup> | Type | Description |
| --- | --- | --- |
| Age<sup>b,c,d</sup> | Integer | Age at admission in full years since birth. |
| ED visits<sup>b,d</sup> | Integer | Visits to the emergency department in the year before admission |
| Ambulance admissions<sup>b,d</sup> | Integer | Admissions to the hospital by ambulance in the year before admission |
| Weeks recently hospitalized<sup>b</sup> | Integer | Full weeks hospitalized in the 90 days before admission |
| Sex<sup>b,c,d</sup> | Factor | Female or Male |
| Living status<sup>b,d</sup> | Factor | Chronic care hospital, Nursing home, Home or Unknown<sup>e</sup> |
| Admission type<sup>b,c</sup> | Factor | Urgent, Semi-urgent, Elective or Obstetric |
| Admission service<sup>b,c,d</sup> | Factor | Cardiac surgery, Cardiology, Critical care, Endocrinology, Family medicine, Gastroenterology, General surgery, Gynecology, Hematology-oncology, Internal medicine, Maxillo-facial surgery, Nephrology, Neurosurgery, Neurology, Obstetrics, Ophthalmology, Orthopedic surgery, Oto-rhino-laryngology, Palliative care, Plastic surgery, Respiriology, Rheumatology, Thoracic surgery, Trauma, Urology or Vascular surgery |
| Admission diagnosis | Binary set | Free-text diagnosis on admission order form mapped to 147 binary variables using regular expressions (see supplement) |
| Comorbidity groups | Binary set | ICD-10 codes from hospital discharge abstracts and ED information systems mapped to 84 binary variables (see supplement) |
| Visible comorbidities | Binary | If a previous hospitalization occurred between five years and six months before admission or if a previous ED visit occurred between six months and two weeks before admission. |
| Flu season<sup>b,f</sup> | Binary | If the current admission is in the month of December, January or February |
| ICU admission<sup>b,d</sup> | Binary | If the current admission is a direct admission to the ICU |
| Urgent 30-day readmission<sup>b,d</sup> | Binary | If the current admission is an urgent readmission within 30 days of a previous discharge |
| Ambulance admission<sup>b,d</sup> | Binary | If the current admission is via ambulance |
| ED admission<sup>d</sup> | Binary | If the current admission is via the ED |

Abbreviations: ED, emergency department; ICD-10, International Statistical Classification of Diseases, Tenth Revision; ICU, intensive care unit.
See supplement for additional details.
<sup>a</sup> All variables except ED admission included in the RF-AdminDemoDx model.
<sup>b</sup> Included in the RF-AdminDemo model.
<sup>c</sup> Included in the RF-Minimal model.
<sup>d</sup> Included in the mHOMR model. For variable transformations and interaction terms, see original specification by Wegier et al.[21]
<sup>e</sup> Unknown if no previous hospitalization between five years and six months before admission.
<sup>f</sup> Models were developed prior to the COVID-19 pandemic; future revisions will likely exclude this variable.

### Perspective

The evaluation of clinical utility was conducted from the perspective of a clinician-led quality improvement team that aims to implement a CDSS to increase the prevalence of GOC discussions for patients with a poor prognosis: a promising initiative[3] for a well-established problem.[5] A necessary component of GOC discussions for hospitalized patients are discussions about code status, the documentation of which had been standardized as CSOs in the institutional EHR since 2015. The documentation of resuscitation preferences for a hospitalized patient with a poor prognosis is a positively-valued, patient-centered outcome in the context of EOL communication[24] and its absence for the same population considered a potentially harmful medical error.[2,25] The main objective of decision analysis was to identify the prediction model that maximized this quality indicator. The secondary objective was evaluating the net benefit[14,15] (NB) of prediction models.

### Alternative strategies

We simulated the operation of a CDSS that uses alternative prediction models for triggering an alert. Conceptually, alerts would suggest discussing GOC, including CPR preferences, if appropriate.[7] The system would generate alerts after midnight for eligible patients admitted the previous day having a predicted risk greater or equal to a certain decisional threshold. Since intervention harm was minimal and time constraints were known to limit GOC discussions,[2,6] we considered the proportion of admissions with an alert, P(Alert), to be the most appropriate criteria for setting model thresholds. We set a P(Alert) of 10% as a point of reference and expected thresholds that identified between 5-20% of CDSS-eligible admissions to be an appropriate range for sensitivity analyses.

The alternative strategies under consideration were the four *mortality alert rules* that resulted from applying to each prediction model a P(Alert)-specific threshold.

### Outcome definitions

Electronic CSOs were linked to hospitalizations in the testing cohort after model development and did not have any role in predictive validation. These orders could convey one of three resuscitation preferences: wants all resuscitation measures (Full code), does not want CPR but wants endotracheal intubation if necessary (DNR/Intubation-OK), and does not want CPR or intubation (DNR/DNI). We considered a CSO to have occurred during a hospitalization if at least one was documented between one week before the admission date and the discharge date, inclusively. The extra week was added to associate a hospitalization with any CSOs documented during observation in the ED prior to hospital admission. The main outcome was a hospitalization with a CSO among those for patients at the EOL, which we defined as death within one year of admission.

### Decision trees

We modelled two decision trees where alerts led to the desired action (Figure 1). The first was based on the conventional assumptions of decision analysis for prediction models,[26,27] where alerts lead to action, and the absence of an alert leads to inaction. The second was a scenario-appropriate adaptation that allowed assessing model utility relative to a strategy of usual care, where alerts lead to action, and the absence of an alert leads to usual care: either action or inaction depending on what had factually occurred for the alert-negative case. For both trees, the benefit to patients of discussing and documenting GOC[2] was attributed to true positives (TPs). The cost of this action is spending clinical time,[2] which was attributed to false positives (FPs). Our valuation procedure and assumptions are further explained in the supplement.

**Figure 1.**
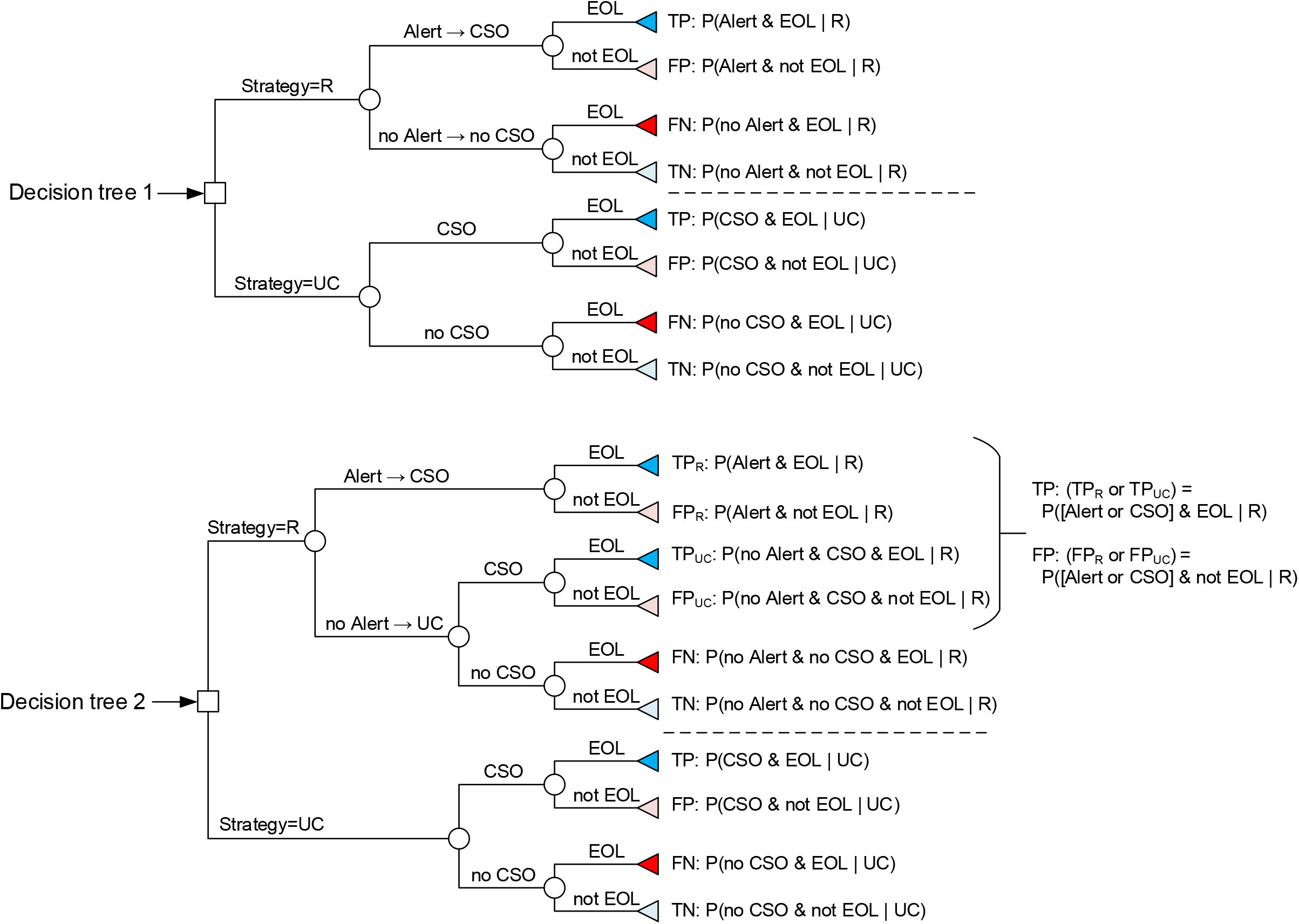
Strategic decision tree models. Abbreviations: CDSS, Clinical decision support system; CSO, code status order; EOL, end-of-life; TP, true positive; FP, false positive; TN, true negative; FN, false negative. Two decision trees modelling the potential outcomes of each hospitalization in the CDSS-eligible cohort under alternative strategies. The strategy of an alert rule, R, implies that a CDSS is implemented and uses R to generate alerts. In the strategy of usual care, UC, CSOs occur as they factually did between July 2017 and July 2018 in the two hospitals of Sherbrooke, Quebec, Canada. In both trees, an alert always implies a CSO. The difference between the two trees is how outcomes unfold in the absence of an alert. In decision tree 1, no alert results in no action (no CSO). In decision tree 2, no alert results in the action that occurred retrospectively in usual care. The first tree models the conventional scenario of decision curve analysis where a prediction rule aims to reduce intervention-related harm, while the second models the scenario of a CDSS that aims to increase a routine good practice that is constrained by time. A TP outcome occurs when a CSO is documented during a hospitalization for a patient who died within one year of admission (EOL status). A FP outcome occurs when a CSO is documented during a hospitalization for a patient who survives more than a year (“not EOL” status). A FN outcome occurs when no CSO is documented during an EOL hospitalization. A TN occurs when no CSO is documented for a “not EOL” hospitalization. The formulas to calculate the expected probability of each outcome for a given strategy are provided to the right of each terminal node.

To distinguish the effect of model-based predictions from the effect of simply generating alerts, we included a fifth model of uniformly random numbers between zero and one. We did not expect alerts from such a model to cause physicians to act in the same way as the validated prediction models, but it would serve to explicit a side-effect of the assumption that all alerts would cause the desired action of a CSO.

### Statistical analysis

We described cohort characteristics stratified by EOL and CSO status. We assessed model discrimination using the C-statistic and its calibration using a calibration plot.[8] To assess construct validity of predictions, we regressed DNR preference against predicted risk in the CDSS-eligible cohort.

Our primary measure of clinical utility was the expected risk difference (eRD) compared to usual care of the main outcome, calculated for each rule as

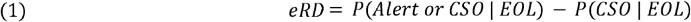

This metric is based on the intention-to-treat estimator[10] and answers the hypothetical question: if every alert based on rule *R* had led to the desired action (a CSO), how many more hospitalizations at the EOL (as a proportion of all hospitalizations at the EOL) would have had a CSO?

For contextualizing the eRD, we calculated the number needed to benefit (NNB):

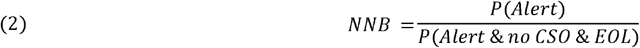

The NNB is the number of alerts needed to increase benefit by one outcome over usual care. It is the reciprocal of the expected benefit among those with an alert: P(*no CSO, EOL* | *Alert*). Conceptually, it incorporates both the number needed to screen, 1/P(*EOL* | *Alert*), and the number needed to treat, 1/P(*no CSO* | *EOL*), as originally described,[16] but it was calculated without assuming conditional independence: the subset of patients at the end-of-life successfully screened with an alert would not necessarily have the same chance of beneficial treatment (the counterfactual outcome in the event of “no CSO”) as the set of all patients at the end-of life.

Our secondary measure of clinical utility was the NB, calculated for each strategy, *S*, as

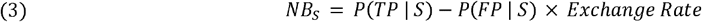

In our scenario, model threshold was based on estimated availability of clinical time, not necessarily patient-provider preference for CSO. This made threshold potentially unsuitable to inform the exchange rate, calculated in conventional decision curve analysis as *Threshold*/(1-*Threshold*).[27] The exchange rate represents the theoretical ratio between the harm of inappropriate inaction (FN) and the harm of inappropriate action (FP), which can be obtained by various means;[14] the threshold method is a convenient simplification in the absence of other utility estimates in the validation set.[27] We calculated a model-independent exchange rate using observed actions of clinicians (i.e., using their *revealed preferences*[14]), who implicitly decide under uncertainty between the harm of inaction and the time-cost of action:

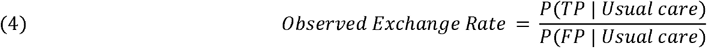

Substituting Equation 4 in Equation 3 results in a NB of zero for the default strategy S = Usual care. We plotted decision curves for both decision trees and for both a threshold-based and observed exchange rate.

We performed a two-way sensitivity analysis[14] between P(Alert) and the exchange rate in subgroups of service type and hospital site. We assessed subgroup heterogeneity using a forest plot of the expected relative risk (the ratio of the terms in the eRD) in relevant inpatient populations.

Consistent with the decision-analytic design, no p-valued significance tests were performed between the alternative strategies.[28] We bootstrapped 95% confidence intervals for all estimates.[8,29] To verify the potential influence of including all hospitalizations in the CDSS-eligible cohort rather than sampling unique patients, we repeated analyses using the first, last, and a random hospitalization per patient. All statistical analyses were performed with R version 3.6.3[30] (relevant extensions and details in the supplement).

## RESULTS

### Sample and model description

The participant flow diagram is presented in Figure 2. Between July 1^st^, 2011 and June 30^th^, 2018, there were 175041 hospitalizations for adults in a non-psychiatric service at our institution (93295 patients). After excluding 76 hospitalizations with rare circumstances, the training cohort included 122860 hospitalizations between July 1^st^, 2011 and June 30^th^, 2016 (70788 patients) and the testing cohort included 26291 hospitalizations between July 1^st^, 2017 and June 30^th^, 2018 (20012 patients). There were 22034 hospitalizations (16490 patients) in the CDSS-eligible cohort. Patient-hospitalization characteristics are presented for the CDSS-eligible cohort in Table 2 (description of other cohorts in the supplement). Prediction models had acceptable temporal validity (Table 3, Figure S1). When sampling over unique patients, the C-statistic ranged from 0.84 to 0.89 in the testing cohort and lowered to 0.79 to 0.86 in the CDSS-eligible cohort. Figure 3 describes EOL process indicators as a function of model-predicted risk; all models had good construct validity for DNR preferences.

**Table 2.** CDSS-eligible cohort characteristics^a^.

|  |  |  | **3 773 hospitalizations at the EOL** |  |
| --- | --- | --- | --- | --- |
|  | **Overall**<br>**(n=22 034)** | **With CSO (n=7 648)** | **2 599 with CSO** | **1 174 without CSO** |
| Age, median (IQR), y | 68 (57-78) | 77 (67-85) | 78 (69-87) | 69 (60-78) |
| Sex |  |  |  |  |
| Male | 11 561 (52) | 3 831 (50) | 1 387 (53) | 665 (57) |
| Female | 10 473 (48) | 3 817 (50) | 1 212 (47) | 509 (43) |
| Hospital site |  |  |  |  |
| A | 13 350 (61) | 3 867 (51) | 1 455 (56) | 909 (77) |
| B | 8 684 (39) | 3 781 (49) | 1 144 (44) | 265 (23) |
| Service type |  |  |  |  |
| Medical | 12 187 (55) | 6 246 (82) | 2 135 (82) | 658 (56) |
| Surgical | 9 266 (42) | 1 023 (13) | 312 (12) | 484 (41) |
| Critical care<sup>b</sup> | 581 (3) | 379 (5) | 152 (6) | 32 (3) |
| Admission type |  |  |  |  |
| Non-elective | 16 780 (76) | 7 298 (95) | 2 525 (97) | 917 (78) |
| Elective | 5 254 (24) | 350 (5) | 74 (3) | 257 (22) |
| Living status at discharge<sup>c</sup> |  |  |  |  |
| Home | 11 205 (51) | 2 663 (35) | 467 (18) | 507 (43) |
| Home with health center (CLSC) liaison | 6 115 (28) | 1 656 (22) | 482 (19) | 404 (34) |
| Short term transitional care | 1 354 (6) | 717 (9) | 201 (8) | 99 (8) |
| Nursing home | 1 612 (7) | 1 143 (15) | 296 (11) | 57 (5) |
| Chronic care hospital | 528 (2) | 454 (6) | 200 (8) | 24 (2) |
| Other<sup>d</sup> | 291 (1) | 151 (2) | 89 (3) | 18 (2) |
| Death in hospital | 929 (4) | 864 (11) | 864 (33) | 65 (6) |
| ED visits<sup>e</sup> |  |  |  |  |
| 0 | 12 786 (58) | 3 457 (45) | 956 (37) | 608 (52) |
| 1-2 | 6 476 (29) | 2 620 (34) | 941 (36) | 383 (33) |
| 3 or more | 2 772 (13) | 1 571 (21) | 702 (27) | 183 (16) |
| Admissions by ambulance<sup>e</sup> |  |  |  |  |
| 0 | 18 547 (84) | 5 432 (71) | 1 644 (63) | 964 (82) |
| 1-2 | 2 891 (13) | 1 760 (23) | 738 (28) | 180 (15) |
| 3 or more | 596 (3) | 456 (6) | 217 (8) | 30 (3) |
| Weeks recently hospitalized<sup>f</sup> |  |  |  |  |
| 0 | 18 601 (84) | 5 925 (77) | 1 774 (68) | 815 (69) |
| 1-2 | 2 617 (12) | 1 237 (16) | 571 (22) | 281 (24) |
| 3 or more | 816 (4) | 486 (6) | 254 (10) | 78 (7) |
| ED admission | 12 711 (58) | 6 329 (83) | 2 155 (83) | 566 (48) |
| Ambulance admission | 7 418 (34) | 4 536 (59) | 1 628 (63) | 297 (25) |
| Urgent 30-day readmission | 2 396 (11) | 1 205 (16) | 596 (23) | 210 (18) |
| ICU admission | 1 036 (5) | 454 (6) | 171 (7) | 51 (4) |
| ICU stay during hospitalization | 3 512 (16) | 1 616 (21) | 538 (21) | 152 (13) |
| Hospital length of stay, median (IQR), d | 4 (2-8) | 7 (4-15) | 9 (4-17) | 4 (2-8) |
| Code status preference<sup>g</sup> |  |  |  |  |
| Full code | 2 323 (11) | 2 323 (30) | 285 (11) | 0 (0) |
| DNR/Intubation-OK | 928 (4) | 928 (12) | 254 (10) | 0 (0) |
| DNR/DNI | 4 397 (20) | 4 397 (57) | 2 060 (79) | 0 (0) |
| Not documented | 14 386 (65) | 0 (0) | 0 (0) | 1 174 (100) |
| Major comorbidities<sup>h</sup> |  |  |  |  |
| Congestive heart failure | 2 758 (13) | 1 728 (23) | 745 (29) | 194 (17) |
| Chronic pulmonary disease | 4 626 (21) | 2 516 (33) | 922 (35) | 265 (23) |
| Dementia | 1 815 (8) | 1 447 (19) | 570 (22) | 77 (7) |
| Metastatic cancer | 1 787 (8) | 848 (11) | 634 (24) | 304 (26) |

Abbreviations: CLSC, *Centre local de services communautaires*; CSO, code status order; DNI, do-not-intubate; DNR, do-not-resuscitate; ED, emergency department; EOL, end of life.
<sup>a</sup> Data given as number (percentage) of hospitalizations unless otherwise indicated. Percentages may not add to 100 due to rounding.
<sup>b</sup> Represent direct admissions to the ICU before a primary non-critical care service could be specified (i.e., the responsible service upon ICU discharge). ICU exposure is more precisely measured with the variables “ICU admission” and “ICU stay during hospitalization”.
<sup>c</sup> See supplement for characteristics of real-time-accessible living status (used for prediction models).
<sup>d</sup> Includes transfer to another hospital, rehabilitation center, palliative care center, or discharge against medical advice.
<sup>e</sup> In the year before admission.
<sup>f</sup> In the 90 days before admission.
<sup>g</sup> Last preference documented during hospitalization if one was documented.
<sup>h</sup> Charlson comorbidities using ICD-10 codes by Quan et al[31] and ascertained using the discharge abstract of index hospitalization and of those in the year before discharge.

**Table 3.** Predictive performance of automatable prediction models for the outcome of one-year mortality.

|  | **RF-AdminDemoDx** | **RF-AdminDemo** | **RF-Minimal** | **mHOMR** |
| --- | --- | --- | --- | --- |
| **Internal validation<sup>a</sup>** |  |  |  |  |
| C-statistic (range) | 0.90 (0.90-0.91) | 0.86 (0.85-0.87) | 0.85 (0.84-0.86) | 0.86 (0.85-0.86) |
| Brier score (range) | 0.068 (0.065-0.073) | 0.079 (0.077-0.083) | 0.082 (0.078-0.084) | 0.081 (0.078-0.085) |
| **External validation<sup>b,c</sup>** |  |  |  |  |
| C-statistic (95% CI) | 0.89 (0.88-0.89) | 0.85 (0.84-0.86) | 0.84 (0.83-0.84) | 0.84 (0.83-0.85) |
| Brier score (95% CI) | 0.074 (0.072-0.076) | 0.084 (0.081-0.086) | 0.086 (0.084-0.089) | 0.086 (0.083-0.088) |
| **CDSS-eligible validation<sup>b,d</sup>** |  |  |  |  |
| C-statistic (95% CI) | 0.86 (0.85-0.87) | 0.81 (0.80-0.82) | 0.79 (0.78-0.80) | 0.80 (0.79-0.81) |
| Brier score (95% CI) | 0.088 (0.085-0.091) | 0.10 (0.097-0.10) | 0.10 (0.10-0.11) | 0.10 (0.099-0.11) |

Abbreviations: CDSS, clinical decision support system.
<sup>a</sup> Internal validity estimated using ten-fold cross-validation in the training cohort (12069-12521 hospitalizations and 7078-7079 patients in each fold). Metrics calculated for each fold after sampling one random hospitalization per patient. Data given as median estimate (range).
<sup>b</sup> Metrics calculated on 1000 two-stage bootstrapped samples as detailed in supplement. Data given as median estimate (95% confidence interval).
<sup>c</sup> Temporal validity estimated in testing cohort (26291 hospitalizations, 20012 patients).
<sup>d</sup> Temporal validity estimated in CDSS-eligible cohort (22034 hospitalizations, 16490 patients).

**Figure 2.**
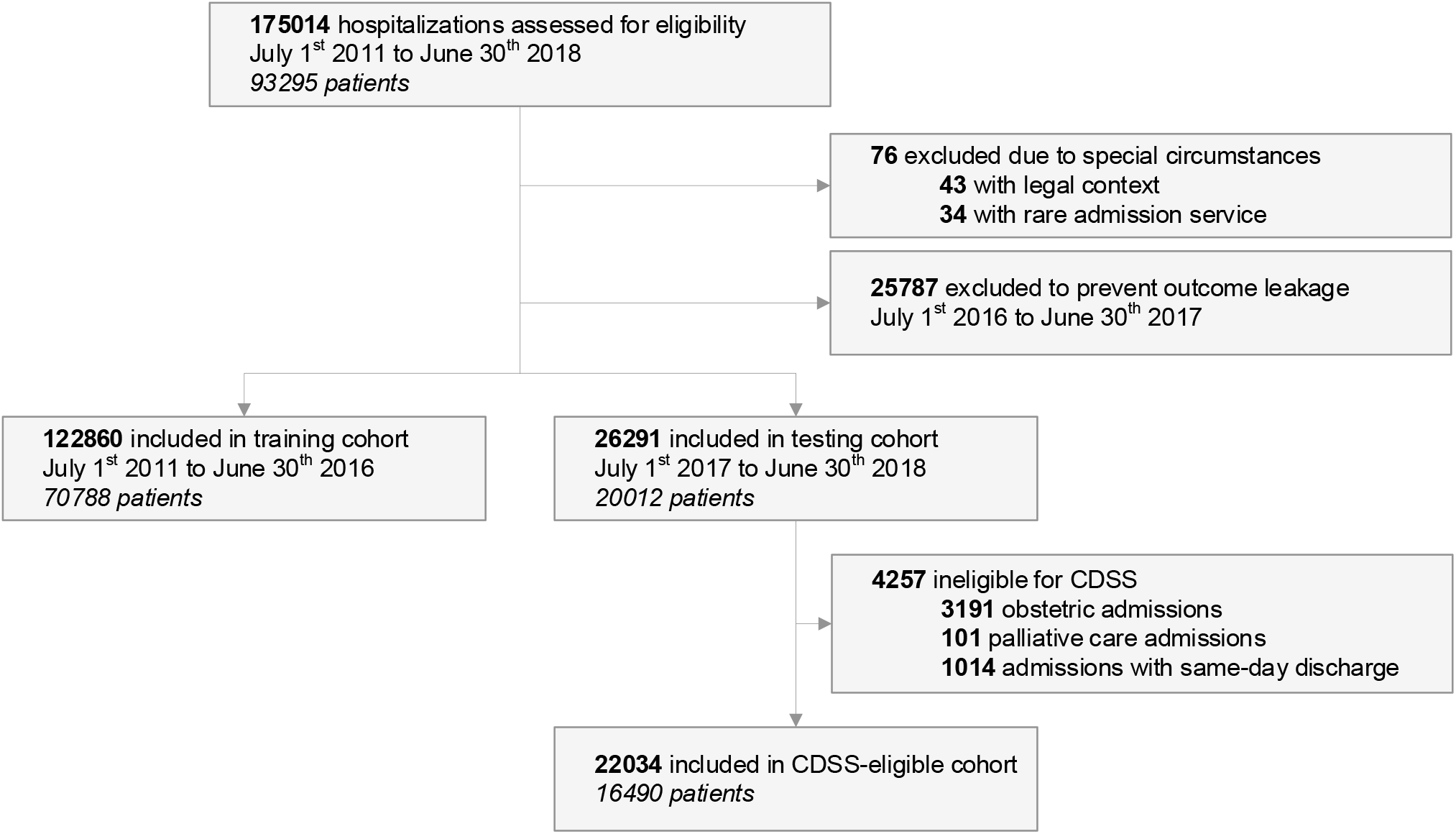
Participant flow diagram. Abbreviations: CDSS, clinical decision support system.

**Figure 3.**
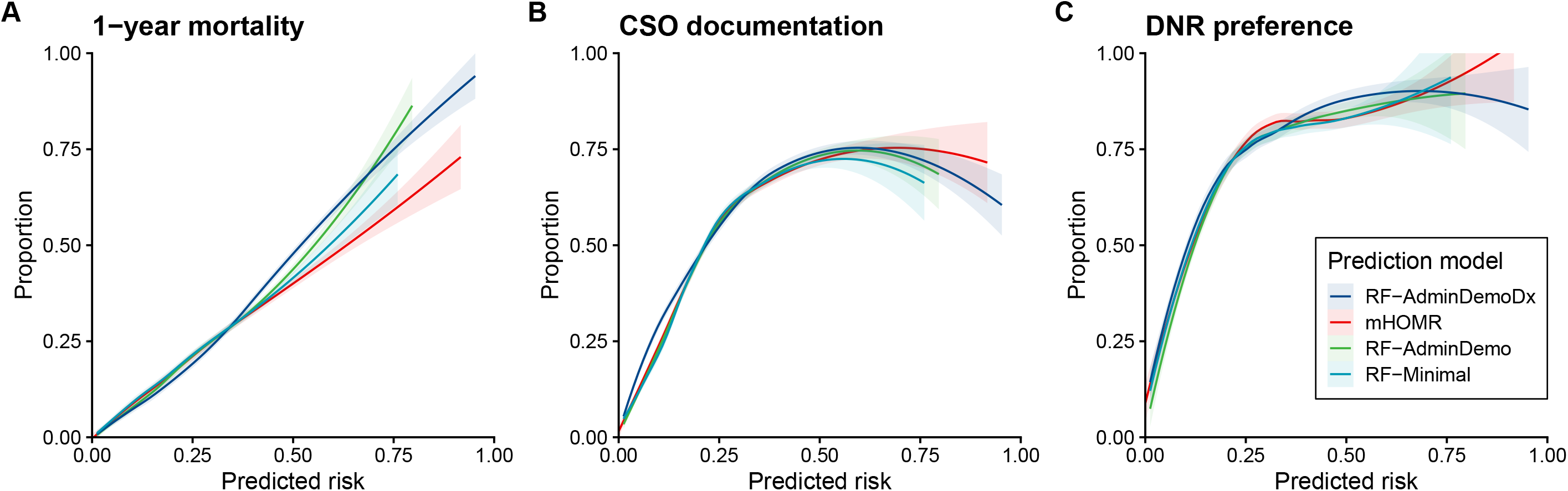
Regression of end-of-life outcome and communication indicators against model-predicted risk. Abbreviations: CSO, code status order; DNR, do-not-resuscitate. Figure caption: Binary variables regressed against predicted risk of a given model then plotted along with 95% CI bands using the loess algorithm. One random hospitalization per patient sampled from the CDSS-eligible cohort before applying regression. Panels A and B include 16490 patients, where 2248 died within one year of hospitalization and 5241 had CSO documentation during that hospitalization. Panel C includes the 5241 patient/hospitalizations with CSO documentation, where 3552 preferred a DNR status in the last CSO documented before discharge. Note that a DNR includes both “DNR/Intubation-OK” and “DNR/DNI”: those without a DNR in Panel C prefer a “Full Code”.

### Code status orders at the end of life

There were 7648 hospitalizations associated with a CSO in the CDSS-eligible cohort (34.7%). Among these, 2599 (34.0%) were associated with death within one-year of admission; clinicians were observed to document a CSO during one hospitalization at the EOL□for every ≈1.9 hospitalizations not at the EOL (observed exchange rate of 2599 TP:5049 FP). On average, clinicians acted as though the harm of FNs was 1.9x as harmful as a FP.

There were 3773 hospitalizations at the EOL in the CDSS-eligible cohort (17.1%). Among these, a CSO was not documented in 1174 cases, meaning a minimal GOC discussion had not been documented for 31.1% of applicable hospitalizations with overnight stays at the EOL. Compared to hospitalizations at the EOL that did have a CSO, these cases were more likely to be elective (OR 9.6 [95% CI, 7.3-12.5]), in surgical specialities (OR 5.1 [95% CI, 4.4-6.1]), for younger patients (mean age 68.3 vs 76.9 years, [95% CI −9.5 to −7.6]), and of shorter duration (mean length of stay 6.5 vs 12.9 days, [95% CI −7.1 to −5.7]).

### Clinical utility

Simulated at a P(Alert) of 10%, each model would have generated on average six alerts per day over one year (Figure 4). At this same level of resource use, the eRD varied between 5.4% and 10.7%, and the NNB between 5.4 and 10.9 alerts (Table 4). The RF-AdminDemoDx model had the highest clinical utility. This model also maximized NB in the decision curves regardless of the decision tree or exchange rate used (Figure 5). When routine clinical actions were considered, only the RF-AdminDemoDx and RF-AdminDemo models could increase value above usual care in the range of desired alert frequency (Figure 5-D).

**Table 4.** Clinical utility of prediction models in the CDSS-eligible cohort.

|  | **RF-AdminDemoDx** | **mHOMR** | **RF-AdminDemo** | **RF-Minimal** | **Random** |
| --- | --- | --- | --- | --- | --- |
| Threshold | 0.478 (0.473-0.483) | 0.461 (0.455-0.467) | 0.465 (0.459-0.470) | 0.422 (0.415-0.424) | 0.898 (0.893-0.902) |
| No. Alerts | 2204 (2204-2207) | 2204 (2204-2208) | 2204 (2204-2207) | 2205 (2204-2229) | 2204 (2204-2207) |
| eRD, % | 10.7 (9.8-11.7) | 5.5 (4.7-6.2) | 6.9 (6.1-7.7) | 5.4 (4.8-6.2) | 3.3 (2.8-3.9) |
| eRR | 1.16 (1.14-1.17) | 1.08 (1.07-1.09) | 1.10 (1.09-1.11) | 1.08 (1.07-1.09) | 1.05 (1.04-1.06) |
| Benefit | 0.1363 (0.1322-0.1407) | 0.1273 (0.1233-0.1318) | 0.1298 (0.1259-0.1346) | 0.1272 (0.1235-0.1318) | 0.1236 (0.1196-0.1282) |
| Harm | 0.1264 (0.1222-0.1311) | 0.1284 (0.1243-0.1330) | 0.1278 (0.1235-0.1323) | 0.1297 (0.1255-0.1347) | 0.1500 (0.1449-0.1554) |
| Net benefit | 0.0099 (0.0080-0.0119) | -0.0011 (-0.0029-0.0006) | 0.0020 (0.0002-0.0039) | -0.0025 (-0.0041 to -0.0006) | -0.0264 (-0.0285 to -0.0243) |
| Standardized net benefit<sup>a</sup> | 0.0578 (0.0465-0.0701) | -0.0062 (-0.0170-0.0037) | 0.0117 (0.0011-0.0224) | -0.0148 (-0.0241 to -0.0036) | -0.1539 (-0.1665 to -0.1428) |
| NNB, Alerts | 5.4 (5.0-6.0) | 10.7 (9.5-12.6) | 8.4 (7.5-9.5) | 10.9 (9.4-12.3) | 17.6 (15.1-21.0) |
| PPV, % | 62.5 (60.2-64.5) | 47.5 (45.3-49.9) | 50.0 (47.9-52.3) | 44.4 (42.4-46.5) | 16.9 (15.4-18.5) |
| NPV, % | 87.9 (87.4-88.4) | 86.3 (85.7-86.7) | 86.5 (86.1-87.0) | 85.9 (85.4-86.4) | 82.9 (82.3-83.3) |
| Sensitivity, % | 36.5 (35.4-37.7) | 27.7 (26.7-28.9) | 29.2 (28.1-30.2) | 26.0 (24.9-27.1) | 9.9 (9.0-10.7) |
| Specificity, % | 95.5 (95.2-95.7) | 93.7 (93.4-93.9) | 94.0 (93.7-94.2) | 93.3 (93.1-93.5) | 90.0 (89.8-90.1) |
| C-statistic<sup>b</sup> | 0.85 (0.84-0.85) | 0.79 (0.78-0.79) | 0.80 (0.79-0.81) | 0.77 (0.77-0.78) | 0.50 (0.49-0.51) |

Abbreviations: eRD, expected risk difference; eRR, expected relative risk; NNB, number needed to benefit; NPV, negative predictive value; PPV, positive predictive value.
All data presented as point estimate (95% CI). Sample size = 22034 hospitalizations; Threshold set such that $P(\text{Alert}) = 10\%$ ; TP CSOs in usual care = 2599 (95% CI, 2512-2698); FP CSOs in usual care = 5049 (95% CI, 4921-5172); Observed exchange rate = 0.515 (0.492-0.540); Prevalence of cases with 1-year mortality = 17.1% (95% CI, 16.7-17.6%); Random alert rule included as a point of reference, not as a strategy under consideration.
<sup>a</sup> Calculated as the net benefit divided by the prevalence of EOL status (1-year mortality), which is the equal to the maximum amount of benefit (if decisions were perfect).
<sup>b</sup> Threshold independent.

**Figure 4.**
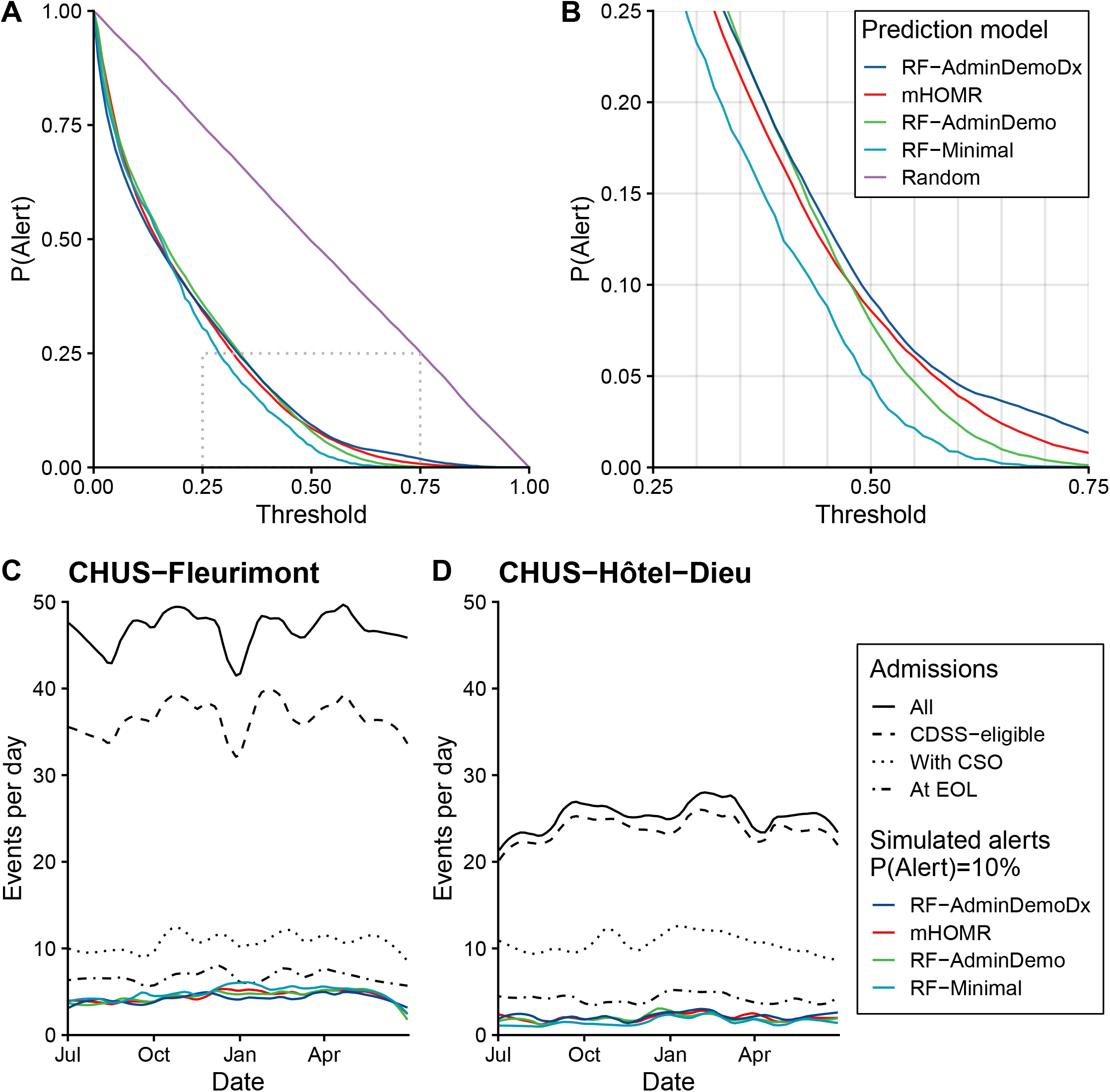
Association between model threshold and proportion of admissions with an alert. Abbreviations: CHUS, Centre Hospitalier Universitaire de Sherbrooke; CSO, code status order; EOL, end-of-life. Panel A: The proportion of CDSS-eligible cases with a predicted risk above the threshold, P(Alert), is plotted against model threshold. A region of interest is outlined where thresholds satisfy reasonable workload demands (5-20% of CDSS-eligible admission). Panel B: The region of interest in Panel A is magnified. Panel C and D: Time-series between July 1^st^, 2017 and June 30^th^, 2018 of daily events stratified by hospital site. The frequency of actual admissions is compared with the frequency of simulated alerts at thresholds satisfying a reference P(Alert) of 10%. “All” refers to all admissions in the testing cohort (excluding pediatric and psychiatric admissions). The loess algorithm was used to smooth day-to-day variations using a span of 0.2 for all curves. CHUS-Fleurimont refers to site A and CHUS-Hôtel-Dieu refers to site B in the main text. Non-CDSS-eligible cases at site A include mostly obstetrical admissions.

**Figure 5.**
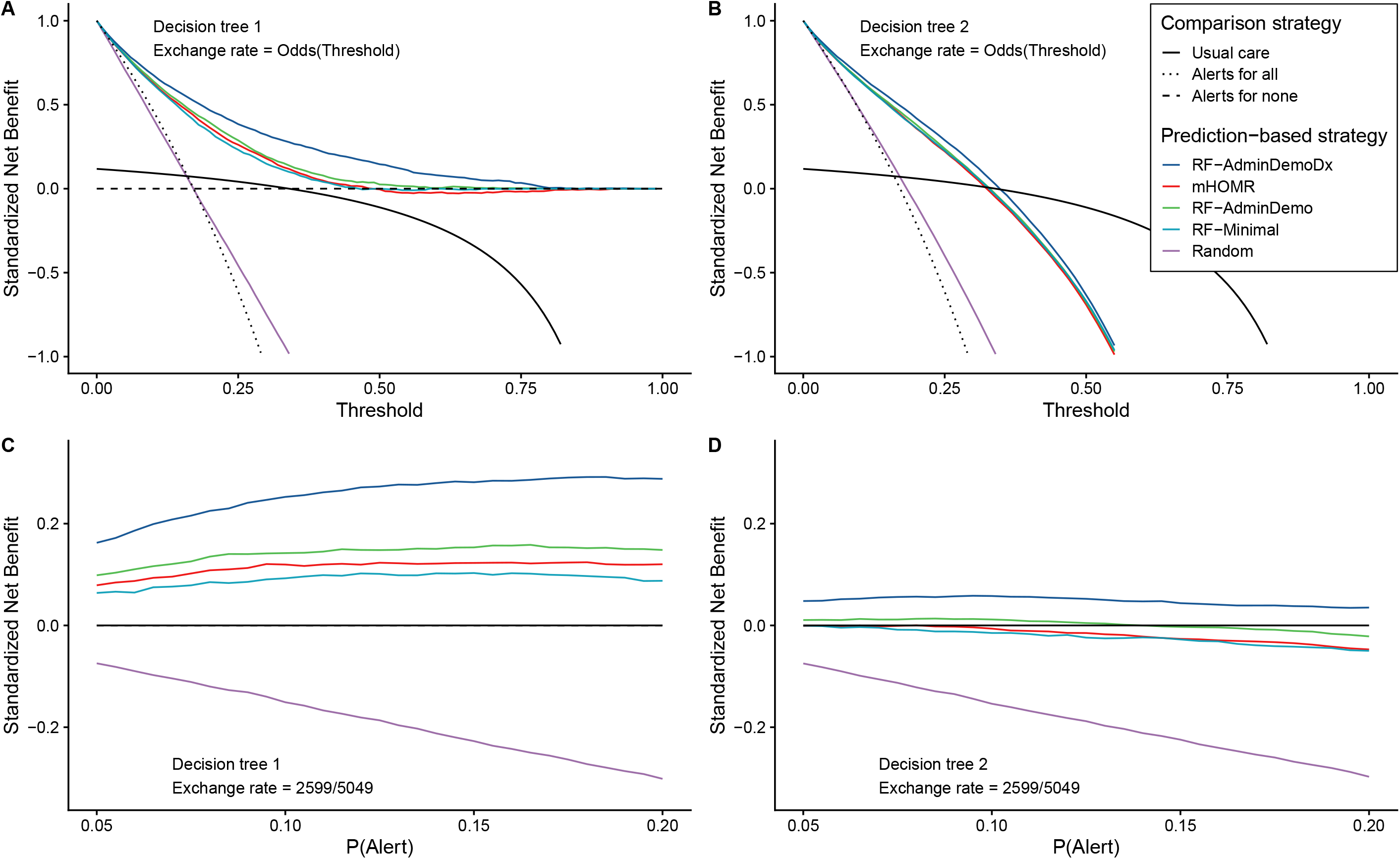
Decision curve analysis. Abbreviations: CSO, code status order; EOL, end of life; FP, false positive; TP, true positive. Decision curves to assess net benefit as a function of either Threshold or P(Alert). Net benefit was standardized by dividing by outcome prevalence (one-year mortality). In the top panels, it is assumed that model threshold is informative of preference between a TP and FP outcome: the exchange rate is calculated as Threshold/(1-Threshold). In the bottom panels, it is instead assumed that observed clinical actions are informative of the exchange rate, which is assigned a constant value: nTP/nFP outcomes in usual care. This value is the odds that a CSO in usual care is a true positive, and as a probability p = nTP/(nTP+nFP), is where the strategy of usual care intersects zero in Panel A. In the left-side panels, the first decision tree is applied to generate TP and FP outcomes, where prediction-based alerts lead to CSO (i.e., “action” or “treatment”) and the absence of an alert leads to no CSO (i.e., “no action” or “no treatment”). In this tree, “Alerts for none” and “Alerts for all” implies “CSO for none” and “CSO for all”, respectively. In the right-side panels, the second decision tree is applied, where alerts lead to CSO, and the absence of an alert leads to usual care: whatever action had factually occurred for a given case. In this tree, “Alerts for all” still implies “CSO for all”, but “Alerts for none” implies “Usual care”. The strategy of “Alerts for all” is a distant outlier in the bottom panels, corresponding to a constant standardized net benefit of around −1.5. The strategy of “Alerts for none” overlaps “Usual care” in panels B to D.

### Sensitivity analysis

The clinical utility of the RF-AdminDemoDx model remained the highest among models in the two-way sensitivity analysis (Figure 6). Subgroup analysis indicated heterogeneity that could influence implementation, including a smaller benefit for all models at site B (Figure S2-S6). Estimates of clinical utility using different sampling strategies did not change the direction or interpretation of results (Table S6-S8, Figure S7-S9).

**Figure 6.**
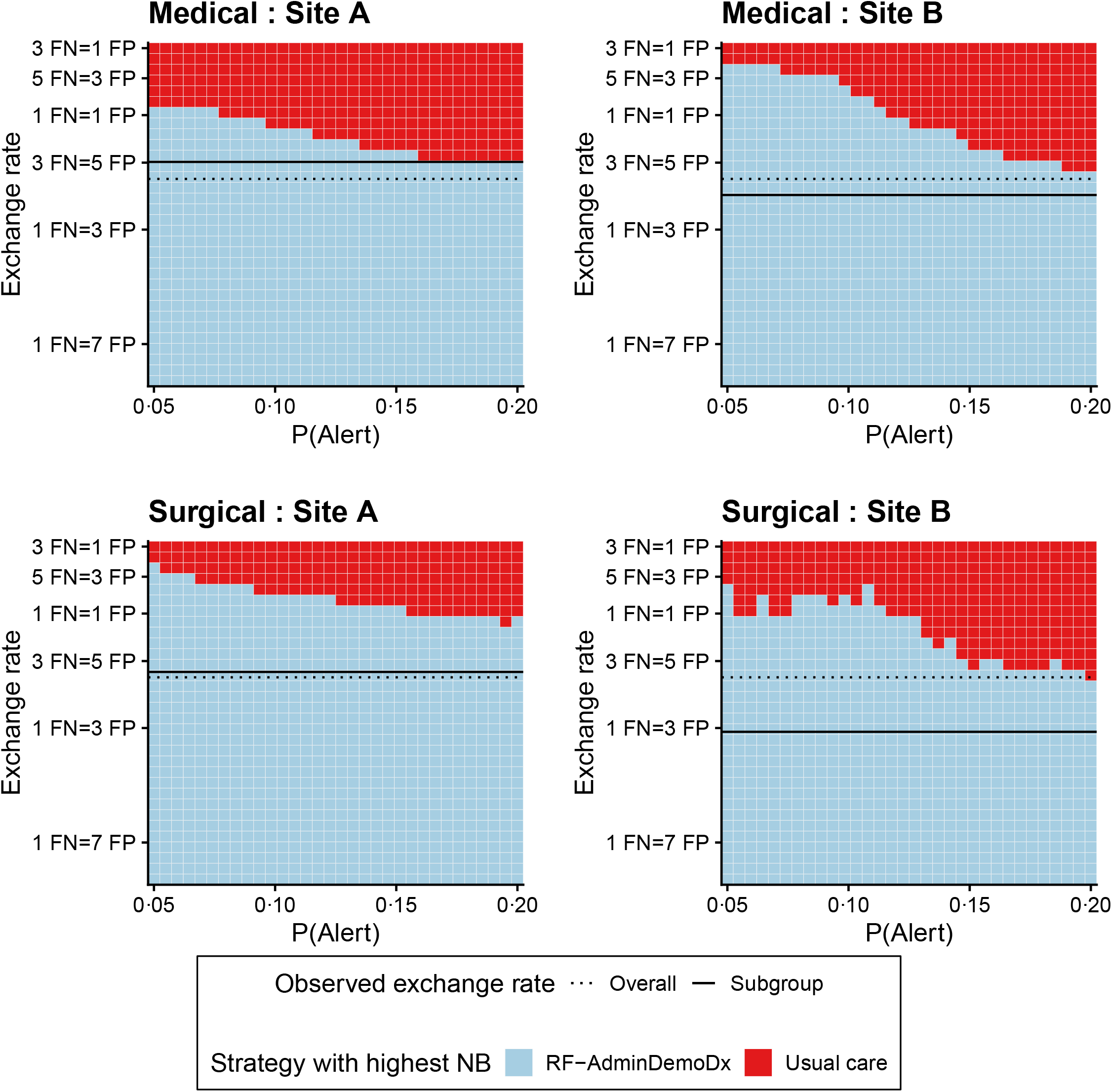
Two-way sensitivity analysis between resource availability and clinical preference. The net benefit (NB) was calculated for each strategy, for each tile and for each plot; the strategy with the highest net benefit is indicated for the corresponding combination of P(Alert) and exchange rate in each hospital site (column) and service type (row). Any ties were resolved by selecting the strategy with the highest benefit, P(TP), or randomly selecting if a tie persisted. A higher exchange rate indicates a higher value to the time-cost of discussing GOC with a patient not at the EOL (worried about FPs), while a lower exchange rate indicates a higher value to the harm of omitting a GOC discussion for a patient at the EOL (worried about FNs). The overall exchange rate (dotted line) was calculated using Equation 4 in the full cohort and the subgroup exchange rate (solid line) corresponds to the result of Equation 4 among a given subgroup. The mHOMR, RF-AdminDemo, and RF-Minimal models are not referenced because they were never a strategy with the highest net benefit.

## DISCUSSION

Improving patient identification for routine PEOLC interventions is a priority for health care stakeholders aiming to reconcile the default policies of life-sustaining therapy with the static truth that all life comes to an end. We performed an up-to-date review of model validation studies in this area of research and provide both a narrative and tabular synthesis of related studies in the supplement. In recent years, there has been a shift from manual screening tools[32] towards automated trigger tools,[33] with the latter shifting from query-based algorithms[34] towards increasingly flexible, but infrastructure-dependant, prediction models.[21,35–38] A challenge with such models is that their usual learning objective, minimizing the error of mortality prediction, is only indirectly related to the clinical objective of maximizing benefit for a resource-limited PEOLC intervention. In this setting of mismatched expectations, predictiveness does not mean usefulness, making it essential to assess clinical utility and not just predictive accuracy.[8–10,16]

In our review of the literature, we did not find any retrospective study evaluating the clinical utility – both benefits and harms – of automatable prediction models for prompting PEOLC interventions. In contrast, almost all studies reported the C-statistic for mortality, and these were generally above 0.8. The context insensitivity of the C-statistic makes it practical for research, but uninformative for practice: more value-based metrics are required to guide decision-makers.[14,17] Prediction models for prompting a PEOLC intervention had varying use-cases for decision support, including GOC discussion, palliative care referral, outpatient follow-up for advance care planning, or hospice referral. The benefits, harms, and resources associated with these actions differ between each other and between health systems; one curve does not fit all.

Strengths of our study included ensuring that retrospectively accessed data represented real-time data and the use of temporal rather than random splitting for validation: simulating prospective application at the point of care. When validated in similar cohorts (not necessarily target population), all models in our study had similar C-statistics as published models (i.e., above 0.8). However, the eRD and NNB for a patient-centered outcome ranged almost twofold, and only two models had a higher NB than usual care with our scenario-appropriate decision tree.

Others have validated the predictive performance of a model, then described physician opinion about the appropriateness of high-risk predictions for intervening.[35,38,39] While informative of construct validity, appropriateness does not represent a model’s usefulness over alternatives. If a mortality alert rule resulted in alerts for every hospitalization – and only hospitalizations – with a DNR in the CDSS-eligible cohort, its PPV for 1-year mortality would be 43.5% (2 314/5 325) and all cases would be appropriate for hypothetical CSO documentation; yet this rule is useless for improving this outcome because it tells clinicians what they already act upon. A similar situation could result from using a model that is highly influenced by terms like “palliative” and “DNR”,[40] or a model that uses historical palliative care consults to predict future consults.[36] Even if alerts correctly predict mortality or benefit, those who would benefit from usual care anyways might be disproportionately identified. More concerningly, those who do not usually benefit may be further marginalized.[7]

Prediction models are often evaluated in biased conditions[41] and rarely compared against routine clinical decision-making.[42] Clinical utility metrics – like an intention-to-treat estimator,[10] the NB,[15] or the NNB[16] – allow for patient-centered comparisons of prediction models with more appropriate assumptions. They can also detect unexpected differences in potential impact, like the difference in utility between our two sites, before any health system investment and exposure to patients. We demonstrated three innovative strategies to increase the applicability of decision analysis for assessing the utility of automatable prediction models.

First, we did not rely on a link between model threshold and clinical preference for net benefit analysis.[27,43] Instead, we linked the threshold to the desired alert frequency, representing resource use, and used other procedures to value outcomes. In doing so, we overcome a limitation of threshold-based NB analysis, which has been remarked as inappropriate for prediction model use-cases that require considering resource availability in addition to patient benefits and harms.[16] Note that the intent behind NB analysis – if not most decision-analytic methods[14] – is that it be adapted and extended to specific scenarios,[27,44] the motivating principle being precisely that off-the-shelf metrics are not necessarily appropriate for all scenarios and stakeholders.[17]

Second, we extended the original decision tree used for decision curve analysis to allow simulating model-*augmented* outcomes (e.g., that no alert can still lead to CSO if clinically appropriate), rather than model-*determined* outcomes (e.g., that no alert will lead to no CSO). We would not want or expect the latter for our use-case. By design, the adapted decision tree results in a more modest estimation of utility, one that accounts for the expected value of routine care: models can only increase benefit if it is there to be increased after applying usual clinical decision-making.

Third, we used empiric rates of TP and FP actions to inform an observed exchange rate. This enabled decision curve analysis while comparing models at the same level of resource use, which was not necessarily the same threshold across models. Among those with a CSO, most patients preferred a DNR when the predicted risk of mortality was above 10-15%, but such a threshold would result in alerts for half of CDSS-eligible admissions. While likely acceptable for patients, who have little to lose and much to gain from a routine GOC discussion, this low threshold could imply unreasonable workloads for clinicians and cause alert fatigue.[12] The observed exchange rate is a simple measure of the benefit-for-time trade-off that limits a good practice with minimal intervention-related harm. It is readily reproducible if practice patterns change over time and we believe insightful about clinical decision-making, noting that physicians may be influenced by an inflated perception of GOC-related cost.[45] This technique could facilitate the clinical utility assessment of other models for improving good practices in time-constrained environments, where utilities cannot be inferred from the model threshold. We used CSOs because they were the only electronic indicator of GOC documentation at our institution, but the same technique could be applied for other standardized indicators of the EOL communication process, like Physician Orders for Life-Sustaining Treatment.[46]

Our reproduction of the mHOMR model did not discriminate one-year mortality as well as in Ontario (c=0.84 vs 0.89),[21] but it was relatively simple to generalize to our institution. We cannot say the same of our ML model, which relies on admission diagnoses in Quebec-local French and would need another free-text mapping to be transportable beyond provincial borders (we report all variable definitions to enable this). However, while the local instance of our ML model is less geographically transportable than mHOMR, it is convincingly more useful for future application at our institution. This finding adds evidence to the recommendation that the pursuit of model generalizability should not be at the expense of local clinical utility.[13]

Our study has several limitations. First, resuscitation preference documentation is an essential but limited measure of EOL communication.[24] We did not measure the quality of the GOC discussions that preceded a CSO nor the concordance of preferences with care received.[5] However, the role of this study was to inform implementation and not substitute a prospective evaluation of clinical impact, where these higher-value patient outcomes should be assessed before long-term adoption.[8] Second, decision analysis requires simplifying assumptions to be practical, like assuming alerts would deterministically lead to action.[10,27] To increase the transparency of these assumptions, we compared prediction models to a random model. Third, due to the COVID-19 pandemic, a repeat validation is likely warranted before local application because models rely on non-causal associations, such as between admission service and death, that may have unexpectedly shifted after systemic reorganization. Finally, although evaluating clinical utility of a prediction model is recommended and provides more value-based metrics than evaluating just predictive performance,[8–10,14–17] more research is required to investigate how well these metrics predict the actual impact of a model-based CDSS. Future studies can refine on decision analysis based on this retroactive feedback, like including model-independent effects from behavioural economic–inspired co-interventions.[47]

## CONCLUSION

An evaluation of clinical utility is recommended after validating a prediction model because metrics of model predictiveness are not informative of value. This is particularly important for mortality prediction models having the use case of automatically prompting a PEOLC intervention, like a GOC discussion. Decision-analytic techniques to assess utility along patient-centered outcomes are feasible for quality improvement teams. They can help discriminate value from hype, calibrate expectations, and provide valuable information before CDSS implementation. As an adjunct to model validation, the routine evaluation of clinical utility could increase the value of automated predictive analytics implemented at the point-of-care.

## Data Availability

The hospitalization data underlying this article cannot be shared publicly due to regulations to protect patient privacy that are overseen by the IRB. All prediction model specifications, decision-analytic model specifications and the observed exchange rate estimates with various sampling procedures are included in the article and its online supplement. The source code for specific aspects of the study, like the data analysis or visualization source code in R or the two-stage bootstrap source code in C++, is available upon request to the corresponding author.

## STATEMENTS

All authors had full access to all the data in the study and accept responsibility to submit for publication.

### Authors’ contributions

*Conceptualization:* RT, JF

*Data curation:* RT

*Formal analysis:* RT

*Funding acquisition:* RT, JF

*Methodology:* RT, JF

*Resources:* RT, JF

*Software:* RT

*Validation:* RT, JF

*Visualization:* RT

*Writing – original draft:* RT

*Writing - review and editing:* RT, JF

## Acknowledgements

We thank Luc Lavoie, associate professor of computer science at the Université de Sherbrooke, for guidance on data management. We thank Paul Farand, MD, MSc, for having led the quality improvement effort that resulted in the migration of paper to electronic code status orders at the *Centre Hospitalier Universitaire de Sherbrooke* and initially suggesting the idea to investigate automated mortality alerts to prompt GOC discussions.

## REFERENCES

1 Detering KM, Hancock AD, Reade MC, et al. The impact of advance care planning on end of life care in elderly patients: randomised controlled trial. The BMJ 2010;340. doi:10.1136/bmj.c1345

2 Bernacki RE, Block SD, American College of Physicians High Value Care Task Force. Communication about serious illness care goals: a review and synthesis of best practices. JAMA Intern Med 2014;174:1994–2003. doi:10.1001/jamainternmed.2014.5271

3 Huber MT, Highland JD, Krishnamoorthi VR, et al. Utilizing the Electronic Health Record to Improve Advance Care Planning: A Systematic Review. Am J Hosp Palliat Med 2018;35:532–41. doi:10.1177/1049909117715217

4 Gill TM, Gahbauer EA, Han L, et al. The role of intervening hospital admissions on trajectories of disability in the last year of life: prospective cohort study of older people. BMJ 2015;350:h2361. doi:10.1136/bmj.h2361

5 Heyland DK, Barwich D, Pichora D, et al. Failure to engage hospitalized elderly patients and their families in advance care planning. JAMA Intern Med 2013;173:778–87. doi:10.1001/jamainternmed.2013.180

6 Lund S, Richardson A, May C. Barriers to advance care planning at the end of life: an explanatory systematic review of implementation studies. PloS One 2015;10:e0116629. doi:10.1371/journal.pone.0116629

7 Porter AS, Harman S, Lakin JR. Power and perils of prediction in palliative care. The Lancet 2020;395:680–1. doi:10.1016/S0140-6736(20)30318-4

8 Steyerberg E. Clinical Prediction Models: A Practical Approach to Development, Validation, and Updating. 2nd ed. Springer International Publishing 2019. doi:10.1007/978-3-030-16399-0

9 Kappen TH, van Klei WA, van Wolfswinkel L, et al. Evaluating the impact of prediction models: lessons learned, challenges, and recommendations. Diagn Progn Res 2018;2:11. doi:10.1186/s41512-018-0033-6

10 Sachs MC, Sjölander A, Gabriel EE. Aim for Clinical Utility, Not Just Predictive Accuracy. Epidemiology 2020;31:359–64. doi:10.1097/EDE.0000000000001173

11 Sendak MP, Balu S, Schulman KA. Barriers to Achieving Economies of Scale in Analysis of EHR Data. A Cautionary Tale. Appl Clin Inform 2017;8:826–31. doi:10.4338/ACI-2017-03-CR-0046

12 Sutton RT, Pincock D, Baumgart DC, et al. An overview of clinical decision support systems: benefits, risks, and strategies for success. Npj Digit Med 2020;3:1–10. doi:10.1038/s41746-020-0221-y

13 Futoma J, Simons M, Panch T, et al. The myth of generalisability in clinical research and machine learning in health care. Lancet Digit Health 2020;2:e489–92. doi:10.1016/S2589-7500(20)30186-2

14 Hunink MGM, Weinstein MC, Wittenberg E, et al. Decision Making in Health and Medicine: Integrating Evidence and Values. 2nd ed. Cambridge: : Cambridge University Press 2014. doi:10.1017/CBO9781139506779

15 Vickers AJ, Calster BV, Steyerberg EW. Net benefit approaches to the evaluation of prediction models, molecular markers, and diagnostic tests. BMJ 2016;352:i6. doi:10.1136/bmj.i6

16 Liu VX, Bates DW, Wiens J, et al. The number needed to benefit: estimating the value of predictive analytics in healthcare. J Am Med Inform Assoc 2019;26:1655–9. doi:10.1093/jamia/ocz088

17 Vickers AJ, Cronin AM. Traditional statistical methods for evaluating prediction models are uninformative as to clinical value: towards a decision analytic framework. Semin Oncol 2010;37:31–8. doi:10.1053/j.seminoncol.2009.12.004

18 Shah NH, Milstein A, Bagley P Steven C. Making Machine Learning Models Clinically Useful. JAMA 2019;322:1351–2. doi:10.1001/jama.2019.10306

19 Adelson K, Lee DKK, Velji S, et al. Development of Imminent Mortality Predictor for Advanced Cancer (IMPAC), a Tool to Predict Short-Term Mortality in Hospitalized Patients With Advanced Cancer. J Oncol Pract 2018;14:e168–75. doi:10.1200/JOP.2017.023200

20 Jung K, Kashyap S, Avati A, et al. A framework for making predictive models useful in practice. J Am Med Inform Assoc Published Online First: 22 December 2020. doi:10.1093/jamia/ocaa318

21 Wegier P, Koo E, Ansari S, et al. mHOMR: a feasibility study of an automated system for identifying inpatients having an elevated risk of 1-year mortality. BMJ Qual Saf 2019;:bmjqs-2018-009285. doi:10.1136/bmjqs-2018-009285

22 Moons KGM, Altman DG, Reitsma JB, et al. Transparent Reporting of a multivariable prediction model for Individual Prognosis Or Diagnosis (TRIPOD): Explanation and Elaboration. Ann Intern Med 2015;162:W1. doi:10.7326/M14-0698

23 Husereau D, Drummond M, Petrou S, et al. Consolidated Health Economic Evaluation Reporting Standards (CHEERS)--explanation and elaboration: a report of the ISPOR Health Economic Evaluation Publication Guidelines Good Reporting Practices Task Force. Value Health J Int Soc Pharmacoeconomics Outcomes Res 2013;16:231–50. doi:10.1016/j.jval.2013.02.002

24 Heyland DK, Dodek P, You JJ, et al. Validation of quality indicators for end-of-life communication: results of a multicentre survey. CMAJ Can Med Assoc J 2017;189:E980–9. doi:10.1503/cmaj.160515

25 Allison TA, Sudore RL. Disregard of patients’ preferences is a medical error: comment on “Failure to engage hospitalized elderly patients and their families in advance care planning.” JAMA Intern Med 2013;173:787. doi:10.1001/jamainternmed.2013.203

26 Cooper GF, Visweswaran S. Deriving the Expected Utility of a Predictive Model When the Utilities Are Uncertain. AMIA Annu Symp Proc 2005;2005:161–5.

27 Vickers AJ, Elkin EB. Decision curve analysis: a novel method for evaluating prediction models. Med Decis Mak Int J Soc Med Decis Mak 2006;26:565–74. doi:10.1177/0272989X06295361

28 Claxton K. The irrelevance of inference: a decision-making approach to the stochastic evaluation of health care technologies. J Health Econ 1999;18:341–64. doi:10.1016/s0167-6296(98)00039-3

29 DiCiccio TJ, Efron B. Bootstrap confidence intervals. Stat Sci 1996;11:189–228. doi:10.1214/ss/1032280214

30 R Core Team. R: A Language and Environment for Statistical Computing. Vienna, Austria: : R Foundation for Statistical Computing 2020. https://www.R-project.org/

31 Quan H, Sundararajan V, Halfon P, et al. Coding Algorithms for Defining Comorbidities in ICD-9-CM and ICD-10 Administrative Data. Med Care 2005;43:1130–9.

32 Walraven C van, McAlister FA, Bakal JA, et al. External validation of the Hospital-patient One-year Mortality Risk (HOMR) model for predicting death within 1 year after hospital admission. CMAJ 2015;187:725–33. doi:10.1503/cmaj.150209

33 Downar J, Wegier P, Tanuseputro P. Early Identification of People Who Would Benefit From a Palliative Approach-Moving From Surprise to Routine. JAMA Netw Open 2019;2:e1911146. doi:10.1001/jamanetworkopen.2019.11146

34 Bush RA, Pérez A, Baum T, et al. A systematic review of the use of the electronic health record for patient identification, communication, and clinical support in palliative care. JAMIA Open 2018;1:294–303. doi:10.1093/jamiaopen/ooy028

35 Major VJ, Aphinyanaphongs Y. Development, implementation, and prospective validation of a model to predict 60-day end-of-life in hospitalized adults upon admission at three sites. BMC Med Inform Decis Mak 2020;20. doi:10.1186/s12911-020-01235-6

36 Murphree DH, Wilson PM, Asai SW, et al. Improving the delivery of palliative care through predictive modeling and healthcare informatics. J Am Med Inform Assoc Published Online First: 21 February 2021. doi:10.1093/jamia/ocaa211

37 Courtright KR, Chivers C, Becker M, et al. Electronic Health Record Mortality Prediction Model for Targeted Palliative Care Among Hospitalized Medical Patients: a Pilot Quasi-experimental Study. J Gen Intern Med 2019;34:1841–7. doi:10.1007/s11606-019-05169-2

38 Parikh RB, Manz C, Chivers C, et al. Machine Learning Approaches to Predict 6-Month Mortality Among Patients With Cancer. JAMA Netw Open 2019;2. doi:10.1001/jamanetworkopen.2019.15997

39 Avati A, Jung K, Harman S, et al. Improving palliative care with deep learning. BMC Med Inform Decis Mak 2018;18:122. doi:10.1186/s12911-018-0677-8

40 Wang L, Sha L, Lakin JR, et al. Development and Validation of a Deep Learning Algorithm for Mortality Prediction in Selecting Patients With Dementia for Earlier Palliative Care Interventions. JAMA Netw Open 2019;2:e196972–e196972. doi:10.1001/jamanetworkopen.2019.6972

41 Wynants L, Calster BV, Collins GS, et al. Prediction models for diagnosis and prognosis of covid-19: systematic review and critical appraisal. BMJ 2020;369:m1328. doi:10.1136/bmj.m1328

42 Liu X, Faes L, Kale AU, et al. A comparison of deep learning performance against health-care professionals in detecting diseases from medical imaging: a systematic review and meta-analysis. Lancet Digit Health 2019;1:e271–97. doi:10.1016/S2589-7500(19)30123-2

43 Pauker SG, Kassirer JP. The Threshold Approach to Clinical Decision Making. N Engl J Med 1980;302:1109– 17. doi:10.1056/NEJM198005153022003

44 Vickers AJ, Cronin AM, Elkin EB, et al. Extensions to decision curve analysis, a novel method for evaluating diagnostic tests, prediction models and molecular markers. BMC Med Inform Decis Mak 2008;8:53. doi:10.1186/1472-6947-8-53

45 Pintova S, Leibrandt R, Smith CB, et al. Conducting Goals-of-Care Discussions Takes Less Time Than Imagined. JCO Oncol Pract 2020;16:e1499–506. doi:10.1200/JOP.19.00743

46 Lee RY, Brumback LC, Sathitratanacheewin S, et al. Association of Physician Orders for Life-Sustaining Treatment With ICU Admission Among Patients Hospitalized Near the End of Life. JAMA 2020;323:950–60. doi:10.1001/jama.2019.22523

47 Manz CR, Parikh RB, Small DS, et al. Effect of Integrating Machine Learning Mortality Estimates With Behavioral Nudges to Clinicians on Serious Illness Conversations Among Patients With Cancer: A Stepped-Wedge Cluster Randomized Clinical Trial. JAMA Oncol 2020;:e204759. doi:10.1001/jamaoncol.2020.4759

