## Supplementary materials for "Clinical Utility of Automatable Prediction Models for Improving Palliative and End-Of-Life Care Outcomes: Towards Routine Decision Analysis Before Implementation"

### Literature review

*Context*

Automated predictive analytics in health care is a rapidly growing field. We performed an up-to-date systematized review to find studies that evaluated the clinical utility of an automatable prediction model for prompting a palliative or end-of-life care (PEOLC) intervention. Our objective was to describe if and how such studies had evaluated clinical utility, as recommended for clinical prediction models.[1,2]

*Methods*

We searched the PubMed database for studies published in any language from inception to March 1st 2021 using the query:

```
(
	(machine learning)
	OR
	(
		(automated OR (real time) OR computerized OR trigger)
		AND
		((prediction model) OR electronic health records[mesh] OR (decision support))
	)
)
AND
(
	(Advance Care Planning) OR "code?status" OR "resuscitation order" OR "palliative care" OR "end?of?life"
	OR "goal#?of?care" OR "serious?illness"
)
```

Our inclusion criteria were prediction model validation studies where the use-case for the model was implementation in a clinical decision support system (CDSS) that automatically identifies patients for a PEOLC intervention. We included prospective and post-implementation studies as long as they included model validation methods (including internal validation, external validation, or an evaluation of clinical utility). To assess the quality of evidence, we appraised the applicability of validated models for real-time application at the point of care using the *participant selection* and *predictor* domains of PROBAST.[3]

*Study selection*

The query returned 334 results, of which two were relevant systematic reviews,[4,5] and after screening abstracts, 33 were initially selected for satisfying the inclusion criteria. The following studies were then excluded:

- Six studies pertaining to query-based algorithms,[6–11] which are decreasingly used in favour of more flexible prediction models.
- Three studies using purchased US Medicare claims data,[12–14] where there was no description of how this data source was accessible for automated predictions at the point of care.
- Two studies of models predicting in-hospital mortality where it was unclear if predictions were intended for earlier palliative care or earlier intensive care.[15,16]
- Two studies where the primary objective was to investigate prognostic factors rather than validate models.[17,18]

*Study characteristics*

We included a final of 20 original research articles and provide a summary in Table S1. There is a climbing interest in the use automated prediction models that use routinely collected data for identifying patients that might benefit from PEOLC interventions. Three studies were published before 2018, three in 2018, six in 2019, five in 2020, and two in the first two months of 2021. Applicable settings were sometimes mixed and included general inpatient populations (9), patients with a specific comorbidity (8; including cancer [5], dementia [1], end-stage liver disease [1], and COVID-19 [1]), general outpatient populations (4), and patients discharged from the ED (2). Five studies reported data after model implementation or application: three pilot studies,[19–21] and two prospective validation studies,[22,23]. Four of these had been considered quality improvement projects by their respective IRBs.[20–23] The actions that models were intended to prompt were sometimes mixed and included palliative care referral (11), goals-of-care discussion (7), outpatient follow-up with ACP (3) and hospice referral (1). There were seven studies that had used a type of external validation (temporal [1], geographic [3], or prospective [3]). Among the six studies in the general inpatient setting that predicted mortality in a timeframe of 6-12 months, four had a C-statistic above 0.8 and two had a C-statistic above 0.9.

*Risk of bias and concerns for applicability in real-time*

Seven of the 15 retrospective validation studies did not adequately describe or correlate retrospective conditions with those an automated CDSS generating predictions at the time and place they would be actionable.[24–30] Eight studies reported model validation metrics in a population that included subgroups where a PEOLC intervention was inappropriate or unnecessary, and could be excluded in real-time without affecting utility. This including patients in obstetrics[19–22,25,30], children,[25,31] or those already receiving palliative care.[19,25,28] While training a model on these cases might prove useful for algorithmic learning, validity should be reported in a subgroup that excludes them, especially when the subsequent application cohort is different.[19,20]

*Evaluation of clinical utility*

None of the studies had assessed the clinical utility of models using decision-analytic methods, including decision curve analysis. One study used decision analysis after model validation, but for quantifying expected financial value from the perspective of reducing system cost, not its clinical value from the perspective of improving patient outcomes.[32]

One study compared model-based predictions with clinical outcomes in usual care: in a clinical utility-related assessment, Parchure et al[33] described rates of inappropriately missing GOC documentation and unmet palliative needs among those with both in-hospital mortality for COVID-19 (outcome-positive) and a predicted risk above a 0.5 threshold for this outcome (high-risk). The study included an assessment of expected benefit among true positives: the benefit of high-risk predictions among outcome-positive cases (9/146 patients with in-hospital mortality could have benefited if every high-risk prediction had led to an appropriate GOC discussion or palliative care referral). However, it did not assess the expected harm of false positives: the harm of high-risk predictions for those that survived to discharge.

All prospective application studies reported the C-statistic as the primary metric to justify the chosen model.[19–21] None reported a decision analysis quantifying the benefits and harms of prediction-based actions relative to usual care before they had proceeded with implementation.

*Conclusion*

Automatable prediction models for improving PEOLC outcomes were not assessed for their clinical utility using decision-analysis. Studies were rarely well coordinated between clinical requirements (e.g., ensuring both expected benefit and expected harms are considered), informatics requirements (e.g., ensuring all required data is accessible in real-time) and methodological requirements (e.g., ensuring models were validated in the target population for a PEOLC intervention, which should exclude pediatric visits at the ED or admissions in obstetrics). Most implemented models were part of quality improvement projects, highlighting the need to consider feasibility and local factors in the translational process of developing, validating, implementing, and evaluating the impact of automatable prediction models.

**Table S1. Validation studies of automatable or automated prediction models intended to prompt a palliative or end-of-life care intervention**

| Reference; Year; Country | Design; Population[a]; Sample size | Model(s) evaluated; Predicted outcome; Validation method | Evaluation metrics[b] (AUROC[c] of best model) | Model-based action; Clinical utility of model[d] | Concerns for applicability at point-of-care[e] |
|---|---|---|---|---|---|
| Escobar et al.;[34] 2015; USA | Retrospective validation; Inpatients surviving to discharge (excluding those for uncomplicated childbirth); n=360036 | 4 LR models; 7/30-day nonelective rehospitalization or mortality; Random split (50/50) | AUROC (0.76), Pseudo-$R^2$ | Outpatient follow-up (implied to include ACP); not reported | .. |
| Adelson et al.;[32] 2017; USA | Retrospective validation; Inpatients with advanced cancer; n=669 | Cox proportional hazards model with proprietary Rothman Index (PeraHealth); 30/60/90/180-day mortality; Random split (70/30) | AUROC (0.74), Sensitivity, PPV, Survival curves | Hospice referral; Potential cost savings if every prompt led to hospice within 48h of admission, but no reporting of patient-centered benefit, or harm of hospice referral for false positives | .. |
| Uneno et al.;[24] 2017; Japan | Retrospective validation; Patients with cancer receiving chemotherapy n=2693 internal validation, n=367 geographic validation | LR model; 1/2/3/4/5/6-month mortality; Geographic validation | AUROC (0.85), Sensitivity, Specificity, PPV, NPV, Accuracy | GOC discussion; Explicitly mention clinical utility not assessed, planned for future study | Exclusion of patients without death records; impossible to reproduce exclusion criteria in real-time. |
| Avati et al.;[25] 2018; USA | Retrospective validation; All patients and inpatients only (including children); n=221284 total and n=10779 inpatients only | DNN model; 3-to-12-month mortality; Random split (80/10/10) | AUROC (0.93 for all patients and 0.87 for inpatients only), AUPRC, Calibration plot, Brier score | Palliative care referral; not reported | Exclusion of patients that died within 3 months of prediction time; impossible to reproduce exclusion criteria in real-time. Did not report validity in target sample excluding unapplicable cases such as those already in obstetrics, palliative care, or children |
| Gensheimer et al.;[35] 2018; USA | Retrospective validation; Patients treated for metastatic cancer; n=12588 | Cox proportional hazards model; Survival curve and mortality within time points from 0.5 to 5 years; Random split (80/20) | AUROC (0.785), Survival curves | GOC discussion; not reported | .. |
| Sahni et al.;[26] 2018; USA | Retrospective validation; Inpatients (excluding non-emergent admissions); n=59848 | 4 models: LR and 3 RF models; 1-year mortality; Random split (80/20) | AUROC (0.86), Calibration plot | ACP (unclear in what setting, presumed to be outpatient); not reported | Uses post-discharge data to evaluate model performance among inpatients; no description of applicability in outpatient setting |
| Beeksma et al.;[27] 2019; The Netherlands | Retrospective validation; Outpatients; n=1234 | LSTM model; life expectancy; Random split (90/10) | Accuracy | Outpatient ACP; not reported | Excludes patients known to have survived more than 5 years. Impossible to reproduce exclusion criteria in real-time |
| Blom et al.;[31] 2019; Sweden | Retrospective validation; Patients discharged from ED (including children); n=65776 internal validation, n=55164 geographic validation | 6 models: LR, RF, SVM, KNN, AB, MLP; 30-day mortality; Geographic split | AUROC (0.95; n.b., cohort included children), Sensitivity, Specificity | Outpatient GOC discussion; not reported, but includes value-focused description of intended use-case from multiple stakeholder perspectives | Did not report validity in target sample excluding unapplicable cases such as children |

| Reference; Year; Country | Design; Population[a]; Sample size | Model(s) evaluated; Predicted outcome; Validation method | Evaluation metrics[b] (AUROC[c] of best model) | Model-based action; Clinical utility of model[d] | Concerns for applicability at point-of-care[e] |
|---|---|---|---|---|---|
| Courtright et al.;[20] 2019; USA[f] | Retrospective validation, CDSS feasibility, and pre-post evaluation; Inpatients (excluding obstetrics, hospice, rehabilitation); n=46305 | LR model; 6-month mortality; Random split (85/15%) | AUROC (0.86 for less selective test cohort), Calibration plot, Sensitivity, Specificity, PPV, NPV, F1-score | Palliative care referral; not reported before application | Did not report validity in target sample excluding unapplicable cases such as in obstetrics |
| Parikh et al.;[36] 2019; USA | Retrospective validation; Outpatients with cancer; n=26525 | 3 models: LR, GBM and RF; 180/500-day mortality; Random split (70/30) | AUROC (0.88), PPV, Calibration plot, Accuracy, Specificity | GOC discussion; not reported | .. |
| Wang et al.;[28] 2019; USA | Retrospective validation; Patients with dementia; n=26921 | LSTM model; 6/12/24 month mortality; Random split (90/10) | AUROC (0.978) | Palliative care referral; not reported | Unclear if ICD codes used for patient inclusion are accessible in real-time. Did not report validity in target sample excluding unapplicable cases such as those already receiving palliative care |
| Wegier et al.;[19] 2019; Canada[f] | Retrospective validation, CDSS feasibility, and pre-post evaluation; Inpatients (excluding psychiatry); n=640022 (based on previously used dataset[37]) | LR model; 1-year mortality; Bootstrap validation | AUROC (0.89 for less selective test cohort), Pseudo-$R^2$, Calibration plot, Sensitivity, Specificity, PPV, NPV, Positive LR, Negative LR | GOC discussion or palliative care referral; not reported, but includes value-focused description of intended use-case from multiple stakeholder perspectives | Did not report validity in target sample excluding unapplicable cases such as in obstetrics |
| Barash et al.;[38] 2020; Israel | Retrospective validation; Patients discharged from ED; n=363635 | GBM; 30-day mortality; Temporal split | AUROC (0.97), Sensitivity, FPR | Palliative care referral; not reported[g] | Could not be assessed[g] |
| Lin et al.;[29] 2020; Taiwan | Retrospective validation; Inpatients with ESLD; n=907 internal validation, n=643 geographic validation | RF and AB models; 30-day and 1-to-9-month mortality; Geographic split | AUROC (0.852) | Palliative care referral; not reported | Unclear if ICD codes used for patient selection are accessible in real-time |
| Major and Aphinyanaphongs;[22] 2020; USA | Retrospective validation, prospective validation, and CDSS feasibility; Inpatients (excluding hospice care); n=128941 | RF model; 60-day mortality; Prospective validation | AUROC (0.87), AUPRC, Calibration plot, PPV | Palliative care referral; harm-to-benefit ratio considered before implementation (preference of 1 TP:3 FP among clinicians) but not applied to evaluate clinical utility | Did not report validity in target sample excluding unapplicable cases such as in obstetrics |
| Manz et al.;[23] 2020; USA | Prospective validation; Patients with cancer; n=24582 | GBM; 180-day mortality; Prospective validation | AUROC (0.89), PPV, NRI compared to standard prognostic indices (ECOG and Elixhauser) | GOC discussions; not reported before application. Compared performance to standard decision-making policies, but did not consider harm-to-benefit ratio when using NRI[39] | .. |
| Parchure et al.;[33]; 2020; USA | Retrospective validation; Inpatients with COVID-19; n=567 | RF model; 20-to-84-hour mortality; Random split (70/30) | AUROC (0.855), Sensitivity, Specificity, Accuracy, AUPRC, Rate of unmet GOC/palliative needs | GOC discussion or palliative care referral; reporting of clinical benefit among cases with in-hospital mortality, but | .. |

| Reference; Year; Country | Design; Population[a]; Sample size | Model(s) evaluated; Predicted outcome; Validation method | Evaluation metrics[b] (AUROC[c] of best model) | Model-based action; Clinical utility of model[d] | Concerns for applicability at point-of-care[e] |
|---|---|---|---|---|---|
| | | | | no reporting or discussion of potential harms | |
| Blanes-Selva et al.;[40] 2021, Spain | Retrospective validation; Inpatients (excluding psychiatry and obstetrics); n=65279 | 5 models: GBM, RF, KNN, SVM, MLP; 1-year mortality; Random split (80/20) | AUROC (0.91), Accuracy, Sensitivity, Specificity, Balanced Error Rate | Palliative care referral; not reported | Unclear if validation sample includes palliative care admissions |
| Guo et al.;[30] 2021; USA | Retrospective validation; Inpatients and outpatients; n=59639 | 4 models: LSTM, RF, DNN, LR; 1-year mortality; Random split (80/10/10) | AUROC (0.92 for outpatients, 0.97 for inpatients), AUPRC, Accuracy, Sensitivity, Specificity, PPV, F1-score | Palliative care referral; not reported | Describes delay of 3-6 months for administrative claims data, but not described if and how this was handled (intended focus of future study). Did not report validity in target sample excluding unapplicable cases such as in obstetrics |
| Murphree et al.;[21] 2021; USA[f] | Retrospective validation, prospective validation, and CDSS feasibility; Inpatients; n=50143 | GBM model; Historic palliative care referral; Random split (80/20) then prospective validation | AUROC (0.91), PPV, Calibration plot | Palliative care referral; not reported before application | Did not report validity in target sample excluding unapplicable cases such as in obstetrics |

Abbreviations: AB, adaptive boosting; ACP, advance care planning; AUPRC, area under the precision recall curve; AUROC, area under the receiver operating characteristic; CDSS, clinical decision support system; DNN, deep neaural network; ESLD, end-stage liver disease; GBM, gradient boosting machine; GOC, goals-of-care; ICD, international classification of disease; KNN, k-nearest neighbours; LR, logistic regression; LSTM, long short-term memory; MLP, multilayer perceptron; NPV, negative predictive value; NRI, net reclassification index; PPV, positive predictive value; RF, random forest; SVM, support vector machine;

[a] Excluding children (age < 18 years) unless otherwise indicated.

[b] Evaluation of model performance or utility. Not included are descriptive metrics (like variable importance measures) or metrics of construct validity (like physician opinion of appropriateness)

[c] Also known as the C-statistic.

[d] Quantitative assessment of benefits and harms of model-based actions relative to a standard policy.

[e] Concerns that validated models do not use data of the target population that is accessible at the intended time and setting of decision support.

[f] Post-application study that included a validation cohort and application cohort. All sample sizes and metrics correspond to assessment in the latest validation cohort.

[g] Only the abstract could be reviewed for this study that was not yet in print at the time of review; full text could not be accessed via institutional library, via request to authors, nor via attempted purchase from journal website, which was impeded by internal server errors over a week, and not fixed before review completion.

### Methods

*Setting*

The study took place at the *Centre Hospitalier Universitaire de Sherbrooke* (CHUS), a public university hospital network in the city of Sherbrooke, Quebec, Canada. It is the only institution offering acute hospitalisation services in the city and is the only tertiary care centre for approximately 1.2 million people in a large catchment area (Eastern Townships),[41] resulting in relatively complete longitudinal data. It is partitioned into two sites, *CHUS-Fleurimont* (site A) and *CHUS-Hôtel-Dieu* (site B). Site A generally offers more specialized services than site B, although the staff for a given speciality cover both.

*Data collection and analysis period*

The start date of July 1st, 2011 coincides with the availability of all necessary predictors in the data warehouse and a large window to use as a training period (five years). The end date of June 30th, 2018 reflects the last date of hospital admissions where the outcome of one-year mortality could be ascertained before the start of analysis in July 2019.

*Mortality outcome*

The binary outcome predicted by all models was the occurrence of death less than 365 days after admission. We considered this timeframe acceptable for approximating EOL status and potential eligibility for palliative interventions.[19,25] Mortality outcome data was collected from the provincial vital statistics registry (*Régie de l'assurance maladie de Québec*) for all patients in the data warehouse with a provincial health number, i.e., official death certificates for all residents of Quebec in the overall cohort (99.5% of hospitalizations). This was cross-referenced with in-hospital mortality records, which includes those for patients with a residence outside of Quebec. Since it was possible for mortality records up until June 30th, 2019 to be missing for a subsequent data extraction in July 2019, death certificates up until that date were re-extracted in October 2020. This increased the prevalence of one-year mortality measured in the testing cohort by 8.2% (308 of 3773 hospitalizations), presumably due to administrative delays. We used the latest mortality data extraction for all analyses and consider it having complete case outcomes.

*mHOMR model*

Wegier et al[19] demonstrated the feasibility of integrating the Modified Hospital One-year Mortality Risk (mHOMR) model into a computerized decision support system (CDSS) for the purpose of motivating discussions about goals of care (GOC). This logistic regression model had been derived from the HOMR model.[37,42] The rationale for modifying the original model was that HOMR included predictors that were coded after hospital discharge and not available for an automated prediction model at admission, including the admission diagnosis, chronic comorbidities and the need for home oxygen. After excluding these predictors, it was feasible to apply the modified model in a CDSS at their institution. The relatively few types of predictors required made it easy to reproduce the mHOMR model at our institution. It was the only mortality prediction model that was both generalizable to our institution and had been tested in a prospective setting for the purpose of prompting a PEOLC–related intervention.

The mHOMR model was reproduced as closely as possible to the published specification. Two categorical variables, the admitting service and living status, were relevelled to align more closely with local practices and to accommodate real time–available data, respectively. All variable transformations (e.g., inverse of the square root of the number of ED visits plus one) and interactions (e.g., between living status and number of ambulance admissions in the last 12 months), were kept as is.

The living status variable was based on the last known value of this variable coded from previous hospitalizations, if present (after simulating a delay, as described later). About half of values were missing and these were imputed as "Unknown", with the assumption that the pattern of missingness – whatever it may be – was transportable to future application at our institution.[43] The living status variable was considered too important for outright exclusion, and its inclusion, even with missingness, improved performance in the training cohort.

All other variable definitions were identical to the original specification. The model was fitted on 1 000 two-stage bootstrap[44] samples of unique patients in the training cohort, where the first stage is the selection of a random patient, with replacement, and the second stage is the selection of a random visit of that patient, with replacement. A simpler technique for fitting a logistic regression model on a large amount of longitudinal data is the selection of a single encounter per patient (e.g., randomly,[42] or the last observed[45]), and discarding the data from all other encounters. In contrast, the two-stage bootstrap procedure allowed considering all hospitalizations for training the model while satisfying the assumption of I.I.D. observations. As was done for the published mHOMR model, the final coefficients were the average bootstrapped coefficients for each variable.

*Machine learning model*
The predictors used by the ML model reflect the same type of information used in the original HOMR model, although sourced and pre-processed differently. In mHOMR, predictors that required diagnostic codes from discharge abstracts had been excluded. While the information from the discharge abstract of an index hospitalization is not available at the time of admission, it may be available for subsequent hospitalizations after some delay due to administrative processing. We surmised that this information, particularly about chronic comorbidities, was important for risk prediction, even if delayed. This was our main reason for developing a new prediction model. Modifications to the original set of variables in HOMR were guided by (1) operational constraints, such as the availability of electronic data in real-time, (2) clinical judgement, such as the grouping of diagnostic codes for personalized risk prediction, and (3) model assumptions, such as the increased flexibility of machine learning algorithms relative to logistic regression.

A total of 244 predictors where included in the final ML model. Among these variables, 147 represented medical terms mapped from a free-text admission diagnosis accessible in real-time from the EHR (Table S2) and 84 represented diagnostic groups mapped from comorbidity information accessible in real-time from administrative databases (Table S3). One variable flagged the absence of accessible comorbidity information. Among the remainder were variables related to demographics (age, sex and living status), previous care utilization (number of weeks hospitalized in the last 90 days, number of admissions by ambulance in the last 12 months and number of ED visits in the last 12 months) and characteristics of the current admission (admission during flu season, admission type, urgent 30-day readmission, admission via ambulance, ICU admission and admitting service). Unlike with the mHOMR model, continuous variables were not transformed before inclusion and no interactions were specified.

Two of these variables were not included in the original HOMR model: if the admission is during the peak months of the local influenza flu season (which we defined as from December to February, inclusively) and the number of full weeks of hospitalisation in the last 90 days. The first variable was motivated by the overall increased prevalence of mortality observed in these months. We reasoned, prior to the COVID-19 pandemic, that including this information could improve the calibration of a machine learning model. The second variable was to add more information on recent care utilization, which we considered important for predicting mortality.

The admission diagnosis was sourced from the free-text transcription of this information in the EHR at the time of admission. A terminology mapping was created manually between this free text variable and a set of medical terms. This was an iterative process conducted by a physician (RT) where the goal was to group common or prognostically relevant expressions without necessarily being exhaustive. Observations in the testing cohort were not included in this process. The resulting mapping is available in Table S2. Each term was included in the model as a binary variable and each hospitalization could have zero or more positive terms. At least one term could be mapped for 90% of admissions in the training cohort. There was no imputation done for admissions without any mapped terms.

We opted to provide the model with relevant diagnostic information from previous visits after simulating an appropriate delay, rather than exclude this information entirely. To inform this delay, the administrative processes that led to electronic data generation were investigated for variables that were not expected to be available at the time of hospital admission. We delayed the visibility of diagnoses from the discharge abstract of a hospitalization by six months after discharge (the usual delay for codification is around three months) and the visibility of the diagnosis from a pre-hospitalization ED stay by two weeks after ED departure (the usual delay for codification is less than one week). Simulating a longer delay than necessary would facilitate the reproducibility of training conditions when the model would be initially applied. Diagnoses coded for a hospitalization that ended less than five years before an index hospitalization were accessible to the index hospitalization (a "lookback" period of five years), and discharge abstracts since January 1$^{st}$, 2007 were available for such a lookback.

The Charlson Comorbidity Index (CCI) was used to group comorbidities in the original HOMR model. The CCI and similar diagnostic grouping algorithms (e.g., Elixhauser comorbidities) have been used in several prediction models for personalized prediction.[22,36,46] While empirically accurate when averaging risk in large cohorts,[45] we did not consider its grouping appropriate for our clinical application. For example, the Charlson comorbidity "Any malignancy, including lymphoma and leukemia, except malignant neoplasm of skin" (Elixhauser comorbidity: "Solid tumor without metastasis") includes both patients with pancreatic cancer (C25) and patients with thyroid cancer (C73), while "Chronic pulmonary disease" (identical Elixhauser comorbidity) includes both patients with chronic obstructive pulmonary disease (J44) and patients with asthma (J45).[47] The diseases in these groups may have similar diagnostic codes, but they do not have similar prognoses.[48] Alternatively, a machine learning model could include diagnoses with little restriction,[25,49] resulting in thousands of distinct diagnostic codes. This strategy might increase predictive performance but the increase in dimensions increases susceptibility to overfitting[50] and requires extra effort to maintain interpretability for our use case.[51,52] We decided to take an intermediate approach, where diagnostic codes where regrouped by a physician (RT) into more prognostically similar subsets of disease: taking advantage of the flexibility of machine learning algorithms but using clinical judgement for every variable added. We surmised that this strategy would yield an acceptable balance between predictive performance and model interpretability. The source of data was considered in the grouping, with a distinction made between (1) diagnoses coded from a pre-hospitalization ED stay (diagnostic codes from ED stays without subsequent hospitalizations could not be extracted), (2) diagnoses coded from the "Principal diagnosis explaining admission" field of discharge abstracts and (3) all diagnoses coded from discharge abstracts (including the principal diagnosis). As an illustration, the prognostic significance of a pulmonary embolism is different if "pulmonary embolism" was the reason for a hospitalization via the ED one month ago, versus if "pulmonary embolism" was mentioned as a past event on a discharge abstract one year ago. ED-associated codes originated from the local instance of the *Système d'information de gestion des urgences* database[53] while discharge abstract–related codes originated from the local instance of the *Maintenance et exploitation des données pour l'étude de la clientèle hospitalière* database[54] (both of which were accessible via the institutional data warehouse). The resulting groupings for each source are available in Table S3.

The codification of the living status variable followed the same timeline as the discharge abstract codification and its visibility was also delayed for six months after discharge. Like for the mHOMR model, the value was imputed as "Unknown" when there was no living status visible for a given hospitalization.

Real-time access from provincial databases was not deemed feasible for the home oxygen variable, and so it was inferred from the ICD-10 code for dependence on supplemental oxygen (Z99.8) from discharge abstracts, subject to the same six-month delay. Archive personnel confirmed the specificity of this code for indicating the mention of home oxygen on the discharge abstracts of our institution.

The ML model was trained with the Random Forest (RF) algorithm.[55] This method was chosen as we found more complex machine learning algorithms (e.g., gradient tree boosting) had modest performance gain, a finding similar to that of others[36], but required more effort to minimize overfitting. Trees were grown using the probability forest method.[56] The number of trees was set to 512 and the number of variables to try per split was set at the default setting of 15 (the floor of the square root of the number of predictors; this was done after ensuring neighbouring values were not more appropriate using a random split in the training cohort). Factor variables were kept as is (rather than one-hot encoded) and partitioned after ordering by outcome.[57(p310)] The sampling algorithm was modified such that the in-bag sample of each tree was selected using the two-stage bootstrap procedure described above.

*Model validation*

We assessed the predictive validity of the models for the outcome of one-year mortality by analyzing usual metrics: discrimination using the C-statistic and calibration with using a calibration plot. We also report the Brier score as an overall metric of validity, although note its limitations.[58] Internal validity is reported to provide an indication of model reproducibility had we not employed temporally external validation, which is more representative of actual performance at the point of care.[59] Note that we did not use internal validation to test optimal variable selection; all variables were either prespecified (originating from HOMR or mHOMR) or based on clinical judgement with reproducible definitions.

K-fold cross-validation (k = 10) was used to estimate the internal validity of the models.[50] Folds were constructed by randomly splitting the set of unique patients into ten groups, then rejoining all hospitalizations to their corresponding patient. This prevented the possibility of leaking a given patient's information between training and testing sets for the $k^{th}$ model. The reported performance measures were obtained by calculating a given statistic in each fold after selecting one random hospitalization per patient.

External validity was estimated in 1 000 two-stage bootstrap resamples of the testing cohort. Note that a given model was trained with the training cohort once and applied to the testing cohort once; each iteration of the bootstrap resampled from the predictions fixed to each observation in the testing cohort. The median value was used as the point estimate and the bias-corrected and accelerated (BCa) method[60] was used to construct 95% confidence intervals.

*Valuation of outcomes*

There are various benefits that can be attributed to the discussion of goals-of-care (GOC) among patients at the end-of-life (EOL), including an improved quality of life, reduced use of aggressive therapy near death, improved adjustment to loss by family, and care that is generally more consistent with patient preferences.[61] Conversely, the absence of GOC discussion in the same population can have the opposite, harmful effects,[61] and could even be considered a medical error of omission.[62] The main cost of discussing GOC is clinician time,[61] a constrained resource.[63]

We used the documentation of code status orders (CSO), encoding the basic preferences for life sustaining therapy, as a process measure of GOC discussions. When a CSO had been documented, it was assumed that a minimal discussion about GOC had occurred. If no CSO had been documented during a hospitalization, then a GOC discussion had not been documented as per hospital policy (CSO documented in the EHR and all other details documented in the paper chart).

A useful prediction model would maximize the benefit of discussing and documenting GOC for patients at the EOL (true positive CSOs) while minimizing the cost of relatively inefficient resource allocation, i.e., when this action occurs for patients not at the EOL (false positive CSOs). We used the recommended[64–67] methods described by Vickers[68] to value these outcomes on the same scale, relative to the benefit of true positives. The value of a true positive is one beneficial outcome, representing the various benefits of a GOC discussion for a patient at the EOL. The value of a false positive is one harmful outcome that is equal in magnitude to a beneficial outcome multiplied by a TP:FP exchange rate.[69,70]

Clinical prediction models are usually intended to be used by clinicians, who incorporate risk and patient preferences to advise patients about proceeding or not with a given intervention (like biopsy or not biopsy).[70] In our context of quality improvement, however, the intention of a prediction model is not to ultimately advise *patients* on some course of action – e.g., between a full code and a DNR: we leave that decision-making between clinicians and patients – but to advise *clinicians* on some course of action: initiate a GOC discussion for a patient at risk of being at the EOL. In this scenario, the exchange rate represents the acceptable number of patients who are at the EOL with whom to discuss GOC (nTP) in exchange for doing so with patients who are not at the EOL (nFP); the exchange rate is equal to nTP:nFP. An equivalent interpretation is the acceptable odds, $p/(1-p)$, of a patient being at the EOL ($p$ = probability of being at the EOL) in order to act in favor of discussing GOC; $p$ = nTP/(nTP+nFP). For example, if it were considered acceptable to have ten GOC discussions if at least one would be for a patient at the EOL, then the exchange rate is 1:9. Equivalently stated, it would be acceptable to spend the time having a GOC discussion with a patient who had at least a 10% chance (p = 1/10) of being at the EOL. In expected utility theory,[63] this would imply that the harm of <u>in</u>action is nine times worse than the harm of action: a rational decision-maker would be expected to act if the chance that <u>in</u>action leads to harm was greater than 10%.

*Decision-analytic assumptions*

To simplify our analysis, we assumed that alerts would deterministically cause the action of CSO documentation (after an appropriate GOC discussion). In reality, it is unlikely that all alerts would be heeded, but this assumption allows us to estimate the maximum benefit attributable to alerts.

While we simulated counterfactual changes in CSO status based on simulated alerts, we did not alter any association between cases and factual EOL outcomes. Communication about GOC and EOL preferences can be associated with increases in life expectancy,[61] but we assumed our definition of EOL status, one-year mortality, to be relatively robust to any such effects. We did not expect that alerts leading to CSO documentation would shorten life to less than a year for those that would have otherwise lived longer or prolong life to more than a year for those that would have otherwise died before.

*Confidence intervals of clinical utility estimates*

A non-parametric bootstrap with 1000 resamples of the CDSS-eligible cohort was used to generate the 95% confidence intervals for all clinical utility estimates (adjusted with the BCa procedure). All intermediate calculation steps were repeated in each bootstrap, including the threshold that resulted from a desired P(Alert) of 10% and the observed exchange rate using Equation 4.

*Statistical software extensions*

The *tidyverse* collection of packages[71] was used for data inspection and preparation. The *ranger* package[72] was used to train the Random Forest models. The two-stage bootstrap described previously was implemented in C++ and linked to R using the *Rcpp* package.[73] The BCa bootstrap procedure was implemented using the source code of the *mediate* function of the *mediation* package.[74] The *ggplot2, cowplot* and *forestplot* packages[75–77] were used for data visualization and generating plots.

**Table S2. Admission diagnosis–mapped variables included in RF-AdminDemoDx model[a]**

<table>
<tr><th>Variable</th><th>Free-text expression[b]</th><th>ICD-9 expression[c]</th></tr>
<tr><td>Abcess</td><td>abces|abcede|collection</td><td></td></tr>
<tr><td>Abdominal pain</td><td>(douleur|dlr)(?:.+)(abdo|epigast)</td><td>789[.]0</td></tr>
<tr><td>Acute coronary syndrome</td><td>(syn(?:.+)\bcoro)|((angor|angine)(?:.+)instable)|(coronarien)|(stemi)|(s(\W)?( )?t(\W)?( )?e(\W)?( )?m(\W)?( )?i)|(\bsca\b)|(infarct(?:.+)myocard)|infarctus|(hemodynamie)|(\bptca\b)|coronaro</td><td>410[.]|411[.]((9[^0])|[^9])</td></tr>
<tr><td>Acute leukemia</td><td>leucemie|\blma\b|\blla\b</td><td>20[458][.][09]</td></tr>
<tr><td>Alcohol</td><td>\broh\b|alcool|ethyli|delirium tremen</td><td></td></tr>
<tr><td>Anemia</td><td>anemie|anemique</td><td>285[.]9|280[.]9</td></tr>
<tr><td>Aneurism</td><td>anevrism|anevrysm</td><td></td></tr>
<tr><td>Angina</td><td>(angine)|(angor)|\bmcas\b|maladie(?:.+)cardiaque</td><td>411[.]90|413[.]|429[.]21|414[.]0</td></tr>
<tr><td>Aortic aneurism</td><td>\baaa\b|anevrism(?:.+)aorte</td><td>441[.]4</td></tr>
<tr><td>Appendicitis</td><td>app[ea]ndicit|appendicect|appendiculaire</td><td>54[013][.]9</td></tr>
<tr><td>ARDS</td><td>\bards\b</td><td></td></tr>
<tr><td>Arrhythmia</td><td>arythmi|arrythmi|(\bcve\b)|(cardioversion)</td><td>427[.][479]|426[.]7</td></tr>
<tr><td>Arthropathy</td><td>arthro|tendon|articula|coxalgie|ankylose</td><td>715[.]</td></tr>
<tr><td>Ascites</td><td>\bascit</td><td>789[.]5</td></tr>
<tr><td>Aspiration pneumonia</td><td>(aspiration)</td><td>507[.]0</td></tr>
<tr><td>Asthma</td><td>asthm[ea]|broncho(\W)?spas[tm]</td><td>493[.]|519.1</td></tr>
<tr><td>Atrial fibrillation</td><td>(\bfa(p)?\b)|(\bflutter\b)|(fibri(?:.+)aur\w+)</td><td>427[.]3</td></tr>
<tr><td>Bariatric</td><td>obesite(?:.+)(severe|morbid)</td><td></td></tr>
<tr><td>Benign tumor</td><td></td><td>2[12][0-9][.]</td></tr>
<tr><td>Biliary colic</td><td>colique(?:.+)biliair|cholelithias|cholestase</td><td>574[.]2</td></tr>
<tr><td>Bladder cancer</td><td>((cancer)|carcinome|(\bneo)|(masse)|(\bca\b))(?:.+)(vessie|vesical|urothelial|ureteral)</td><td></td></tr>
<tr><td>Brain cancer</td><td>(\bgbm\b)|(glioblasto)|astrocytom|\bgliome</td><td>191[.]</td></tr>
<tr><td>Brain hemorrhage</td><td>(\bhip\b)|((hematom|hemor)(?:.+)(\bft\b|occip|cerebral|front|parietal|temporal|cerebell|fosse|parench|thalam|cortical|(intra(.)?(ventric|cranien))))|((\bavc\b|\bacv\b|accident cerebro)(?:.+)hemor)</td><td>431[.]</td></tr>
<tr><td>Brain injury</td><td>(\bhsd\b)|(hematom|hemor)(?:.+)(dural)|\btcc\b|(trauma|contusion)(?:.+)(\bcranien|cerebra)</td><td>432[.]1|85[14][.]0</td></tr>
<tr><td>Brain lesion</td><td>(masse|lesion|tumeur)(?:.+)(cerebral|frontal|cerebell|parietal|occipital|temporal)</td><td></td></tr>
<tr><td>Breast cancer</td><td>((cancer)|(carcinome)|(\badenoca)|(\badenok)|(\bneo)|(ad(n?)k)|(\bca\b))(?:.+)(\bsein\b|mammaire|canalaire|lobulaire)</td><td>174[.]9</td></tr>
<tr><td>Bronchiectasis</td><td>bronchiect</td><td></td></tr>
<tr><td>Bronchitis</td><td>bronchite|bronchiolite|surinfection bronchiq</td><td></td></tr>
<tr><td>C. Difficile</td><td>\bc(?:.+)diff</td><td></td></tr>
<tr><td>Cancer</td><td>(cancer)|(\badenoca)|(\badenok)|(\bneo)|(\bad(n)?k\b)|neoplasi|\bcarcino|anaplasi|sarcome|(\bca\b)|hodgkin|(\blnh\b)|(lymphome)|\blma\b|melanome|leucemie</td><td></td></tr>
<tr><td>Carotid stenosis</td><td>st(?:.+)(carotid|\bci[gd]\b)|endarterect</td><td>433[.]12</td></tr>
<tr><td>Cellulitis</td><td>\bcellulit</td><td>682[.][67]</td></tr>
<tr><td>Chemotherapy</td><td>(\bcotx\b)|(chimio)|(\bcia\b)</td><td></td></tr>
<tr><td>Chest pain</td><td>\bdrs\b|douleur(?:.+)(thora|retro(\W)?stern)</td><td>786[.]5[01]</td></tr>
<tr><td>Cholangitis</td><td>cholangit</td><td>576[.]</td></tr>
<tr><td>Cholecystitis</td><td>cholecystit|cholecystect</td><td>575[.]</td></tr>
<tr><td>Choledocholithiasis</td><td>choledocho|\bcpre\b|(calcul|lithias)(?:.+)(choledo)</td><td>574[.]5</td></tr>
<tr><td>Chronic leukemia</td><td>(synd\w+(?:.+)(myelo|lympho))|(waldenstrom)|\bllc\b</td><td>20[45][.]1</td></tr>
</table>

<table>
<tr><th>Variable</th><th>Free-text expression[b]</th><th>ICD-9 expression[c]</th></tr>
<tr><td>Cirrhosis</td><td>\bcirrho|varice(?:.+)oeso|hemorr(?:.+)(varic|oeso)|\bencephalop(?:.+)hepat</td><td>571[.]5|572[.]21|456[.]1</td></tr>
<tr><td>Colitis</td><td>\bcolite|pancolite</td><td></td></tr>
<tr><td>Colorectal cancer</td><td>((cancer)|(\badenoca)|(\badenok)|(\bneo)|(ad(n?)k)|(tumeur)|(masse)|(\bca\b))(?:.+)(coliq|\bcaec|sigmoid|\bcolo|rect|appendice|\bgrele\b)</td><td>15[34][.]</td></tr>
<tr><td>Conduction abnormality</td><td>bloc(?:.+)([23]e|complet|\ba[. -]?v\b|auricul)|\bb[.]?a[.]?v\b|mobitz|bradycard|\bb[.]?b[.]?g\b|(\bsick sinus\b)|(\bsss\b)|maladie(?:.+)sinus|(\bpause(s)?\b)|(tachy(?:.*)brady)</td><td>426[.][0-689]</td></tr>
<tr><td>COPD</td><td>(mpoc\b)|(malad\w+ pulm\w+ obstr\w+ chron\w+)|(m(\W)?( )?p(\W)?( )?o(\W)?( )?c)|emphysem</td><td></td></tr>
<tr><td>Cytopenia</td><td>pancytopeni|bicytopeni</td><td></td></tr>
<tr><td>Delirium</td><td>(\bdelire\b)|(delirium)|confusion|((alteration|perte|etat)(?:.+)conscience)|\bagitation|\baec\b</td><td>293[.]|307[.]9|348[.]3|331[.]0</td></tr>
<tr><td>Dementia</td><td>\bdemenc|alzheimer|(\bt(?:.+)(cognitif|comportement))|\btnc\b|\berrance\b</td><td>29[04][.]|310[.]1</td></tr>
<tr><td>Diabetes</td><td>diabet|(\bd[bm]\b)|hyperglycemie|hypoglycemie|etat hyper[ ]?(gly|osm)</td><td>250[.]09|251[.]2|790[.]6|250[.]</td></tr>
<tr><td>Dialysis</td><td>irct|irc terminal|\bdialyse</td><td></td></tr>
<tr><td>Diarrhea</td><td>(diarrh)</td><td></td></tr>
<tr><td>Disk disorder</td><td>hernie(?:.+)(disc|[dls]\d)|discectomie|discoidect|discopathie|discarthro|radiculopathie|lombo-sciatalgie</td><td>722[.]|724[.][3]</td></tr>
<tr><td>Diverticular disease</td><td>diverticul(it|os|ai)</td><td>562[.]</td></tr>
<tr><td>DVT</td><td>emboli(?:.+)pulm|\bt[vp]p\b|t(h)?rombophleb|thrombose veineuse|phlebite|embolie massive|ep massive</td><td>415[.]1|451[.][28]</td></tr>
<tr><td>Dysphagia</td><td>\bachalasie|\bdysphagi</td><td>787[.]2</td></tr>
<tr><td>Dyspnea</td><td>\bdyspne</td><td>799[.]0|786[.]03</td></tr>
<tr><td>Ear disorder</td><td>mastoidit|vertige|\boto|labyrinth|cholesteatom|neuron(?:.+)vestibul|\bsurdite\b|\bmeniere\b</td><td>38[0-9][.]|780[.]42</td></tr>
<tr><td>Electrolytes</td><td>(hypo|hyper)(na|ca|k|mg|mag)|deshydrat</td><td>276[.]1|276[.]0|276[.]8|276[.]7|275[.]42</td></tr>
<tr><td>Endocarditis</td><td>endocardit</td><td>424[.]9|421[.]01</td></tr>
<tr><td>ENT cancer</td><td>((cancer)|(\badenoca)|(\badenok)|(\bneo)|(ad(n?)k)|(\bca\b))(?:.+)(\borl\b|larynx|pharynx|langue)</td><td>161[.]</td></tr>
<tr><td>Enteritis</td><td>enterit|\bileite</td><td></td></tr>
<tr><td>EOL care</td><td>(palliati)|soins confort|phase terminal</td><td></td></tr>
<tr><td>Electrophysiology study</td><td>(e p s)|(electrophysio)|(\beps\b)|\bdefib|\bpace(?:.+)defib|pace(.)?maker|\bpace\b|(\bpmp\b)</td><td></td></tr>
<tr><td>Esophageal cancer</td><td>((cancer)|(\badenoca)|(\badenok)|(\bneo)|(ad(n?)k)|(\bca\b))(?:.+)(oesophag)</td><td>150[.]9</td></tr>
<tr><td>Esophageal varices</td><td>varice(?:.+)oeso|hemorr(?:.+)(varic|oeso)</td><td>456[.]1</td></tr>
<tr><td>Eye</td><td>oeil|\bretin|ophthalm</td><td></td></tr>
<tr><td>Falls</td><td>\bchute(s)?\b|t(?:.+)marche\b</td><td>781[.]2</td></tr>
<tr><td>Febrile neutropenia</td><td>(neutro|pancytopenie)(?:.+)febr|fievre(?:.+)neutro</td><td>288[.]02</td></tr>
<tr><td>Fertility</td><td>fertilit</td><td></td></tr>
<tr><td>Fracture</td><td>(fractur)|(\bfx\b)|(\b[#]\b)|rofi</td><td></td></tr>
<tr><td>Gastric cancer</td><td>((cancer)|(\badenoca)|(\badenok)|(\bneo)|(ad(n?)k)|(\bca\b))(?:.+)(gastr|estomac)</td><td>151[.]9</td></tr>
<tr><td>Gastritis</td><td>gastrit</td><td>535[.]5</td></tr>
<tr><td>GI bleed</td><td>\bhd[hb]\b|rectorragie|melena|hemorr(?:.+)digest</td><td>578[.]|569[.]31</td></tr>
<tr><td>Guillain Barré syndrome</td><td>guil(?:.+)bar|mill(?:.+)fis|\bcidp\b</td><td>357[.]0</td></tr>
<tr><td>Gynecological disorder</td><td>\badenomyos|ligature|\bendometrios|endometrite|\bhysterect</td><td>62[56][.]|620[.]2|617[.]0|621[.]3</td></tr>
<tr><td>Heart failure</td><td>(\bins\w* cardia)|(def\w+ cardiaq)|(dysf\w+(?:.+)(\bvg\b|ventricul|diastol|systol))|(\bcmp\b)|(ic(?:.+)(decompens|diastol|systol))|surcharge|(oedem(?:.+)(pulm|poumon))|(\boap\b)|anasarque</td><td>414[.][89]|428[.][19]|(518[.](4|812))|425[.]4|429[.]9|276[.]60</td></tr>
<tr><td>Hemoptysis</td><td>hemoptys</td><td>786[.]3</td></tr>
</table>

<table>
<tr><th>Variable</th><th>Free-text expression[b]</th><th>ICD-9 expression[c]</th></tr>
<tr><td>Hepatic failure</td><td>\bins(?:.+)hepatiq</td><td>573[.]81</td></tr>
<tr><td>Hepatitis</td><td>hepatit</td><td>573[.]3|571[.]1</td></tr>
<tr><td>Hip fracture</td><td>([#]|fx|fracture|rofi)(?:.+)(hanche|femur|femoral|troch|intertro|(bas)(?:.+)cervic|peri(-?)pro|(sous)(?:.+)capi)</td><td></td></tr>
<tr><td>Hypertension</td><td>hypertensi|\bhta\b</td><td></td></tr>
<tr><td>Infection</td><td>(\b(sur)?infect)|\bfievre|febrile|hypertherm|temperature</td><td></td></tr>
<tr><td>Inguinal hernia</td><td>hernie(s)? ing</td><td>550[.]</td></tr>
<tr><td>Intestinal ischemia</td><td>((enterite|colite)(?:.+)ischemique)|((ischemie|angine)(?:.+)(mesenter|intestin))</td><td>557[.][09]</td></tr>
<tr><td>Intestinal polyp</td><td>polyp(?:.+)(coliq|\bcaec|sigmoid|\bcolo|rect|appendice|\bgrele\b|intestin|\bileo)</td><td>211[.]3</td></tr>
<tr><td>Intoxication</td><td>(poly)?intox|sevrage (dro[gq]ue|medicam)|toxicom|abus drogue</td><td>977[.]|291[.]|30[345][.]|292[.]0</td></tr>
<tr><td>Joint prosthesis</td><td>\bpth\b|prot\w+ tot\w+</td><td></td></tr>
<tr><td>Liver cancer</td><td>cholangiocarc|carcinome hepat|hepato(-?)carcinome|hepatome|\bchc\b</td><td></td></tr>
<tr><td>Loss of autonomy</td><td>(pert\w+ ((d'autonomie)|(dautono\w+|autono\w+)))|deconditionnement|epuisement</td><td></td></tr>
<tr><td>Lower leg fracture</td><td>([#]|fx|fracture|rofi)(?:.+)(cheville|tibia|rotule|mal(l)?(eol|[.])|perone|jambe|genou|pied|patella|maisonneuve)</td><td></td></tr>
<tr><td>Lumbar pelvis fracture</td><td>([#]|fx|fracture|burst)(?:.+)(pelvi|pubi|\bacetab|bassin|\bl(\W)?[1-5]|lombaire|sacrum)</td><td>80[58][.]</td></tr>
<tr><td>Lung cancer</td><td>((cancer)|(\badenoca)|(\badenok)|(\bneo)|(ad(n?)k))(?:.+)(poumon|pulmon|\bl[si][gd]\b|lobe (inf|sup|moy))</td><td>16[23][.]</td></tr>
<tr><td>Lung mass</td><td>(masse|nodule|lesion|tumeur)(?:.+)(poumon|pulmon|\bl[si][gd]\b|lobe (inf|sup|moy))|\bbtt\b|biopsie(?:.+)trans(?:.+)thorac</td><td></td></tr>
<tr><td>Lymphoma</td><td>lymphome|\blnh\b</td><td>202[.]</td></tr>
<tr><td>Melanoma</td><td>\bmelanom</td><td>172[.]</td></tr>
<tr><td>Meningitis</td><td>meningit|encephalit</td><td>32[0-4][.]</td></tr>
<tr><td>Metastasis</td><td>metasta[st]|carcinomatose|epanchement(?:.+)neo|\bst(?:.+)\b(4|iv)(.)?\b|mesotheliom</td><td>19[789][.]</td></tr>
<tr><td>Inflammatory bowel disease</td><td>(crohn)|(colite ulc)</td><td>555[.]|556[.]9</td></tr>
<tr><td>Multiple myeloma</td><td>(myelom\w+ multip)</td><td>203[.]</td></tr>
<tr><td>Osteomyelitis</td><td>osteomyelit|(plaie|pied)(?:.+)(infect|diabet)</td><td>730[.]2</td></tr>
<tr><td>Osteoporosis</td><td>osteoporo</td><td>733[.][01]</td></tr>
<tr><td>Other hernia</td><td>hernie (inc|omb)</td><td>55[1-3][.]</td></tr>
<tr><td>Pancreatic cancer</td><td>((cancer)|(\badenoca)|(\badenok)|(\bneo)|(ad(n?)k)|(\bca\b))(?:.+)(pancrea)</td><td>157[.]9</td></tr>
<tr><td>Pancreatic mass</td><td>(masse|tumeur|lesion)(?:.+)pancrea(s|tique)</td><td></td></tr>
<tr><td>Pancreatitis</td><td>pancreatite</td><td>577[.]0</td></tr>
<tr><td>Parkinson’s</td><td>parkinson</td><td>332[.]0</td></tr>
<tr><td>Perforation</td><td>perforation(?:.+)(intestin|grele|colon|colique|caec|jeju|duoden|ilea)|(ulcere|ulcus|divertic|chole|appendic|)\bperfore|pneumoperit</td><td></td></tr>
<tr><td>Pericardial effusion</td><td>epanchement(?:.+)pericard</td><td></td></tr>
<tr><td>Pericarditis</td><td>(myo|peri)(?:.*)cardite</td><td>423[.]91</td></tr>
<tr><td>Pleural effusion</td><td>\bepanchement(?:.+)(pleural|pleureux)</td><td>511[.][0-9]</td></tr>
<tr><td>Pneumonia</td><td>(pneumonie)|empyeme</td><td>48[0123568][.]</td></tr>
<tr><td>Pneumothorax</td><td>pneumothora|\bptx\b</td><td>512[.]</td></tr>
<tr><td>Pregnancy</td><td>travail|(cesarien)|(grossess)|(\d+ semain)|(rupt\w+ prematu\w+)|(\brppm\b)|(rciu)|(placenta)|(accouche)|(post([- ])?partum)|(gestation)|fertilit</td><td>6[3-7][0-9]|[vV]22[.]2</td></tr>
<tr><td>Prolapsus</td><td>\bprolap(a)?sus|(relachement|relaxation)(?:.+)(pelvien)|(cysto|recto|colpo|entero)(?:.*)cele\b|procidence|\bcolpo</td><td>618[.]|788[.]3</td></tr>
<tr><td>Prostate cancer</td><td>((cancer)|(carcinome)|(\badenoca)|(\badenok)|(\bneo)|(ad(n?)k)|(\bca\b))(?:.+)(prostat)|prostatect</td><td>185[.]</td></tr>
</table>

<table>
<tr><th>Variable</th><th>Free-text expression[b]</th><th>ICD-9 expression[c]</th></tr>
<tr><td>Pulmonary fibrosis</td><td>(fibrose)(?:.+)(pulm)|\bfpi\b</td><td>515[.]</td></tr>
<tr><td>Pulmonary hypertension</td><td>((hypertension|htn) pulm)|\bhtp\b</td><td></td></tr>
<tr><td>PVD</td><td>(ische(?:.+)((\bgr)|(membre)|(pied)|(mollet)))|(claudic)|(((\bins(?:.+))|(isch(?:.+)))((art)|(vascul)|(mi[dg])))|(amputat)|\bfrein\b|(\b(arterio|angio)(\b|graph|plast))</td><td>443[.]9|440[.]9|459[.]932|785[.]4</td></tr>
<tr><td>PVD and gangrene</td><td></td><td>785[.]4</td></tr>
<tr><td>PVD and arterial insufficiency</td><td></td><td>443[.]9|440[.]9</td></tr>
<tr><td>PVD and ischemia</td><td></td><td>459[.]932</td></tr>
<tr><td>Reanimation</td><td>\bacr\b|arret cardi|reanimation</td><td>427[.]5|798[.]1</td></tr>
<tr><td>Renal failure</td><td>(\bi(\W)?r(\W)?a\b)|(\bi(\W)?r(\W)?c\b)|(\bins(?:.*) renal)</td><td>58[1-3][.]|584[.]90|586[.]9</td></tr>
<tr><td>Respiratory failure</td><td>((\bins|detresse|difficulte)(?:.+)respirat)|\bdesatur|hypoxemi</td><td>786[.]01|786[.]0\b</td></tr>
<tr><td>Seizures</td><td>status(?:.+)(epile|convuls|partiel)|(epilep(s|t)i)</td><td>780[.]3|345[.]9</td></tr>
<tr><td>Sepsis</td><td>(sepsis)|(\bseptiqu)</td><td>038[.][0-489]|790[.]7</td></tr>
<tr><td>Severe</td><td>\bsevere\b</td><td></td></tr>
<tr><td>Shock</td><td>\bchoc\b(?! ana)(?! hem)(?! hypo)</td><td>785[.]5</td></tr>
<tr><td>Spondylopathy</td><td>stenose(?:.+)(foram|spinal|lombaire|cervical|[cdtls]\d)|myelopathie|laminect|compression medul(l)?aire|foraminect|spondylolisthesi</td><td></td></tr>
<tr><td>Stroke</td><td>(\bavc\b)|(a v c)|(acc(?:.+)cerebr(?:.+))|(\bacv\b)|(a(\W)?v(\W)?c)|\bavc\b|acc(?:.+)vasc(?:.+)cere(?:.+)</td><td>436[.]9|784[.]3|342[.]</td></tr>
<tr><td>Subarachnoid hemorrhage</td><td>(hematom|hemor)(?:.+)arachnoid|\bhsa\b</td><td>430[.]90</td></tr>
<tr><td>Syncope or orthostatic hypotension</td><td>\bsyncop|tension(?:.+)orthosta|lipoth[yi]mi</td><td>780[.]2|458[.]0|458[.]9</td></tr>
<tr><td>Tachycardia</td><td>(\bt(\W)?s(\W)?v)|(tachycardie)|\bt(\W)?v\b|arythmie(?:.+)ventric|(\bt(\W)?a(\W)?p)</td><td>427[.][0-2]</td></tr>
<tr><td>Tamponade</td><td>tamponnade|tamponade</td><td></td></tr>
<tr><td>Thyroid cancer</td><td>((cancer)|carcinome|(\bneo)|(masse)|nodule|tumeur|(\bca\b))(?:.+)(thyroid|folliculaire|papillaire)|thyroidect</td><td>193[.]</td></tr>
<tr><td>Transient ischemic attack</td><td>(\bi(\W)?c(\W)?t\b)|((isch|paresie)(?:.+)transitoir)|amauros(?:.+)fuga</td><td>435[.]</td></tr>
<tr><td>Tonsillitis</td><td>amygdalite|amygdalect</td><td></td></tr>
<tr><td>Trauma</td><td>trauma|laceration|plaie|(luxation)|(rupture)</td><td>95[89][.]|86[0-9][.]</td></tr>
<tr><td>Trigeminal neuralgia</td><td>nevralgie(?:.+)trijum</td><td></td></tr>
<tr><td>Tumor</td><td>(tumeur)|(nodule)|(masse)|(lesion)|(adenome)|meningiome|adenopathie</td><td></td></tr>
<tr><td>Urinary lithiasis</td><td>calcul|lithias|colique neph|coraliform|nephrostomie|\bjj\b</td><td></td></tr>
<tr><td>Urinary retention</td><td>retention</td><td>788[.]2</td></tr>
<tr><td>Urological procedure</td><td>\brtutv\b|\brtuv\b|hbp|\brtup\b|\bhp\b|\baps\b|prostate|\bhbp\b|hematurie</td><td></td></tr>
<tr><td>Urinary tract infection</td><td>(\bpna\b)|(pyelonephrit)|(p n a)|(urosepsis)|(\bcystite)|(infec\w+ urinair)</td><td>590[.][18]|599[.]0|595[.]9|601</td></tr>
<tr><td>Valve prosthesis</td><td>bentall|tiron(?:.+)david|(\brvm\b)|(\brva\b)</td><td></td></tr>
<tr><td>Valve regurgitation</td><td>(\bins(?:.+)|regurg(?:.+))(mitral|aort)</td><td>424[.][01]1</td></tr>
<tr><td>Valve stenosis</td><td>(\bstenos(?:.+)(mitral|aort|\bao\b))|\bsa\b</td><td>424[.]13|39[45][.]0</td></tr>
<tr><td>Virus</td><td>\bsag\b|(synd(?:.+)(gripp|viral))|grippe|grippal|virus|rhume|influenz|\bi(\W)?v(\W)?r(\W)?s\b</td><td>465[.]91|487[.]1|079[.]9</td></tr>
<tr><td>Weight loss or fatigue</td><td>perte de poids|(\bdeg\b)|(((dim\w+)|(det\w+))(?:.+)(etat g))|faiblesse|etat(?:.+)general|\basthenie</td><td>783[.]2|780[.]7</td></tr>
</table>

Abbreviations: ARDS, acute respiratory distress syndrome; COPD, chronic obstructive pulmonary disease; DVT, deep venous thrombosis; ED, emergency department; ENT, ear, nose, throat; EOL, end-of-life; GI, gastro-intestinal; ICU, intensive care unit; PVD, peripheral vascular disease.

[a] Admission diagnoses are transcribed from the physician-written order form into the EHR by administrative clerks at the time of admission. The EHR has a list of standard ICD-9 codes that may be attached to the admission diagnosis. When they are selected, both the code and its standard label are concatenated with the free-text admission diagnosis by the EHR. The resulting, real-time–accessible variable is an unstandardized string of characters that was

preprocessed to make all letters lowercase and to strip accents. Each row defines a binary variable included as a predictor in the RF-AdminDemoDx model. For each variable, if either expression matches the preprocessed string, the variable value is true (or 1), otherwise it is false (or 0).
[b] Regular expressions are intended for use with admission diagnoses in french, including terms and abbreviations that are specific to medical language in Quebec.
[c] Regular expressions that match ICD-9 codes embedded in admission diagnosis string.

**Table S3. Comorbidity diagnosis–mapped variables included in the RF-AdminDemoDx model[a]**

| Variable[b] | ED diagnosis[c] | Summary diagnosis[d] | Principal diagnosis[e] |
|---|---|---|---|
| Atrial fibrillation | I48 | I48 | |
| Acute respiratory failure | J960 | J960 | |
| Alcohol-related diagnosis | F10 E52 G621 I426 K292 Z721 | F10 E52 G621 I426 K292 Z721 | |
| Anasarca | E877 R60 | E877 R60 | |
| Anemia | | D5[1-3] D649 D638 | |
| Recent angina | I20 | | |
| Anticoagulation | | Z921 | |
| Aortic stenosis | | I350 | |
| Ascites | R18 | R18 | |
| Asthma | J45 | J45 | |
| Breast cancer | | C50 Z853 | |
| Bronchiectasis | J47 | J47 | |
| Cachexia | R64 R53 | R64 R54 | |
| CAD | | I20 I25 Z955 | |
| Chemotherapy-related diagnosis 1 | | Z926 | |
| Chemotherapy-related diagnosis 2 | Z511 D700 | Z511 | |
| Chest cancer 1 | | Z85[12] C3[0-39] | |
| Chest cancer 2 | C34 | C3[4-8] | |
| CHF | | I099 I11 I13 I255 I42[05679] I43 I50 P290 | |
| CHF-related admission | I099 I11 I13 I255 I42[05679] I43 I50 P290 | | I099 I11 I13 I255 I42[05679] I43 I50 P290 |
| Chronic respiratory failure | J96[19] | J96[19] | |
| CNS cancer | C71 | C71 | |
| COPD/emphysema | J44[01] J43 | J44 J43 | |
| CVD | G4[56] H340 I6[0-9] | G4[56] H340 I6[0-9] | |
| Dementia | | F0[0-3] F051 G30 G31 | |
| Malnutrition | | E4[36] | |
| Depression | | F204 F31[345] F32 F33 F341 F412 F432 | |
| Dialysis | | Z992 Z49 | |
| Diabetes-related diagnosis 1 | | E1[0-4][09] | |
| Diabetes-related diagnosis 2 | E1[0-4] G632 H360 N083 | E1[0-4][1-8] G632 H360 N083 | |
| Endocrine cancer | | C7[3-5] | |
| Falls | R296 W06 W1[89] R268 | R296 W06 W1[89] | |
| Frailty | R64 R53 R296 W06 W1[89] R268 | E4[36] R64 R54 R296 W06 W1[89] | |
| GI cancer 1 | | Z850 C1[789] C20 | |
| GI cancer 2 | C18 | C1[56] C2[4] | |
| GI cancer 3 | | C2[12356] | |
| GU cancer 1 | | C5[124] C560 C6[1467] Z85[45] | |
| GU cancer 2 | C57 C61 | C5[3] C6[02358] | |
| GU cancer 3 | | C5[57] C56[1-9] | |
| Hematological cancer 1 | | C8[12358] Z85[67] C921 C9[16] | |
| Hematological cancer 2 | | C8[46] C9[04] | |
| Hematological cancer 3 | C8[1-9] C9[0-6] | C9[35] C92[04589] | |
| Home oxygen | | Z998 | |
| Crohn's disease | K50 | | K50 |
| Interstitial lung disease | | J84 | |
| Hepatic disease 1 | | B18 K70[01239] K71[3457] K7[34] K759 K76[2-489] Z944 K86[01] | |
| Hepatic disease 2 | K7[1-6] Z944 | K704 K711 K72 K758 K76[57] | |
| Hepatic disease risk factors | | I85[09] I864 I98[23] K766 | |
| Metastatic solid cancer | | C7[789] C80 | |
| Past MI | | I21 I22 I252 | |
| Recent MI | I21 I22 I252 | | |
| Musculoskeletal cancer | | C4[015-9] | |
| Obesity-hypoventilation syndrome | | E662 | |
| Obstetrics-related diagnosis | O Z37 | | O Z37 |
| ENT cancer | | C0[1-9] C1[0-4] | |
| Palliative | | Z515 | |
| Paralysis | | G041 G114 G80[12] G8[12] G83[0-49] | |

| Variable[b] | ED diagnosis[c] | Summary diagnosis[d] | Principal diagnosis[e] |
|---|---|---|---|
| Past pulmonary embolism | | | I26 |
| Respiratory-related admission | | | J4[0-467] J684 J70[13] |
| Pneumonia admission | | | J1[2-8] |
| Pseudomonas-related diagnosis | | B965 J151 | |
| Psychiatric disorder | | F2[02-589] F302 F31[25] | |
| Pulmonary hypertension | I27 | I27 | |
| Peripheral vascular disease | I7[01] I73[189] I771 I79[02] K55[189] Z95[89] | I7[01] I73[189] I771 I79[02] K55[189] Z95[89] | |
| Recent abdominal pain | R104 | | |
| Recent anemia | D649 | | |
| Recent back pain | M545 | | |
| Recent cancer-related diagnosis | C50 Z853 C767 | | |
| Recent chest pain | R074 | | |
| Recent colitis | A099 | | |
| Recent complication | T8 | | |
| Recent GI bleed | K922 | | |
| Recent hip fracture | S72 | | |
| Recent interstitial nephritis | N10 | | |
| Recent intestinal occlusion | K56 | | |
| Recent pulmonary embolism | I26 | | |
| Recent perforation | K578 | | |
| Recent pneumonia | J1[2-8] | | |
| Recent urinary tract infection | N390 | | |
| Renal disease 1 | N189 | I12 I131 N03[2-7] N05[2-7] N18[1238] N19 Z940 | |
| Renal disease 2 | | N18[045] N250 | |
| Skin cancer | C44 | C4[34] | |
| Transplantation-related diagnosis | | Z94[0-4] | |
| Valvular disease | | I0[5-8] I3[4-9] Z95[2-4] | |

Abbreviations: CAD, coronary artery disease; CHF, congestive heart failure; CNS, central nervous system; COPD, chronic pulmonary obstructive disease, CVD, cerebrovascular disease; ED, emergency department; ENT, ear, nose, throat; GI, gastro-intestinal; GU, genito-urinary; MI, myocardial infarction.

[a] Diagnoses given as space-delimited regular expressions intended to be applied to period-stripped ICD-10 codes. Each row defines a binary variable included as a predictor in the RF-AdminDemoDx model. For each row, if any of the expressions match any of their respectively sourced ICD-10 codes (ED, summary or principal), the variable value is true (or 1), otherwise it is false (or 0).

[b] Numbered variables are categorized by prognostic similarity; higher numbers generally indicate diagnoses with worse prognosis.

[c] Diagnoses associated with an ED visit that led to hospitalization between six months and two weeks before index admission.

[d] Any diagnosis on a hospital discharge abstract between five years and six months before index admission.

[e] Principal diagnosis on a hospital discharge abstract between five years and six months before index admission.

**Table S4. Characteristics of hospitalizations between July 2011 and July 2016 (Training cohort)[a]**

| | Overall (n=122 860) | Vital status one year after admission | |
|---|---|---|---|
| | | Dead (n=18 541) | Alive (n=104 319) |
| Age, median (IQR), y | 64 (45-76) | 75 (64-84) | 62 (40-74) |
| Sex | | | |
| Male | 55 695 (45) | 9 981 (54) | 45 714 (44) |
| Female | 67 165 (55) | 8 560 (46) | 58 605 (56) |
| Hospital site | | | |
| A | 79 868 (65) | 11 260 (61) | 68 608 (66) |
| B | 42 992 (35) | 7 281 (39) | 35 711 (34) |
| Service type | | | |
| Medical | 59 763 (49) | 13 469 (73) | 46 294 (44) |
| Surgical | 45 085 (37) | 4 561 (25) | 40 524 (39) |
| Critical care[b] | 1 479 (1) | 500 (3) | 979 (1) |
| Obstetrics | 16 533 (13) | 11 (0) | 16 522 (16) |
| Admission type | | | |
| Non-elective | 83 125 (68) | 16 771 (90) | 66 354 (64) |
| Elective | 24 136 (20) | 1 764 (10) | 22 372 (21) |
| Obstetrics | 15 599 (13) | 6 (0) | 15 593 (15) |
| Real-time–accessible living status[c] | | | |
| Home | 58 214 (47) | 11 046 (60) | 47 168 (45) |
| Nursing home | 1 938 (2) | 688 (4) | 1 250 (1) |
| Chronic care hospital | 952 (1) | 414 (2) | 538 (1) |
| Unknown | 61 756 (50) | 6 393 (34) | 55 363 (53) |
| Living status at discharge | | | |
| Home | 61 055 (50) | 5 401 (29) | 55 654 (53) |
| Home with CLSC liaison | 42 590 (35) | 4 699 (25) | 37 891 (36) |
| Short term transitional care | 6 574 (5) | 1 439 (8) | 5 135 (5) |
| Nursing home | 4 589 (4) | 1 087 (6) | 3 502 (3) |
| Chronic care hospital | 2 226 (2) | 857 (5) | 1 369 (1) |
| Other[d] | 1 161 (1) | 393 (2) | 768 (1) |
| Death in hospital | 4 665 (4) | 4 665 (25) | 0 (0) |
| ED visits[e] | | | |
| 0 | 73 817 (60) | 7 796 (42) | 66 021 (63) |
| 1-2 | 35 847 (29) | 6 842 (37) | 29 005 (28) |
| 3 or more | 13 196 (11) | 3 903 (21) | 9 293 (9) |
| Admissions by ambulance[e] | | | |
| 0 | 105 595 (86) | 12 979 (70) | 92 616 (89) |
| 1-2 | 14 439 (12) | 4 432 (24) | 10 007 (10) |
| 3 or more | 2 826 (2) | 1 130 (6) | 1 696 (2) |
| Weeks recently hospitalized[f] | | | |
| 0 | 105 957 (86) | 12 690 (68) | 93 267 (89) |
| 1-2 | 12 700 (10) | 4 157 (22) | 8 543 (8) |

| | Overall (n=122 860) | Vital status one year after admission | |
|---|---|---|---|
| | | **Dead (n=18 541)** | **Alive (n=104 319)** |
| 3 or more | 4 203 (3) | 1 694 (9) | 2 509 (2) |
| Visible comorbidities[c] | 67 865 (55) | 13 602 (73) | 54 263 (52) |
| Admission during flu season[c] | 30 280 (25) | 4 736 (26) | 25 544 (24) |
| ED admission | 62 448 (51) | 12 956 (70) | 49 492 (47) |
| Ambulance admission | 36 333 (30) | 9 368 (51) | 26 965 (26) |
| Urgent 30-day readmission | 11 493 (9) | 3 810 (21) | 7 683 (7) |
| ICU admission | 3 173 (3) | 811 (4) | 2 362 (2) |
| ICU stay during hospitalization | 17 119 (14) | 3 368 (18) | 13 751 (13) |
| Length of stay, median (IQR), d | 3 (2-7) | 7 (3-14) | 3 (2-6) |
| Major comorbidities[g] | | | |
| Congestive heart failure | 12 202 (10) | 3 769 (20) | 8 433 (8) |
| Chronic pulmonary disease | 22 181 (18) | 5 709 (31) | 16 472 (16) |
| Dementia | 7 324 (6) | 2 459 (13) | 4 865 (5) |
| Metastatic cancer | 8 618 (7) | 5 192 (28) | 3 426 (3) |

Abbreviations: CLSC, *Centre local de services communautaires*; DNI, do-not-intubate; DNR, do-not-resuscitate; ED, emergency department; EOL, end of life.

[a] Data given as number (percentage) of hospitalizations unless otherwise indicated. Percentages may not add to 100 due to rounding.

[b] Represent direct admissions to the ICU before a primary non–critical care service could be specified (i.e., the responsible service upon ICU discharge). ICU exposure is more precisely measured with the variables "ICU admission" and "ICU stay during hospitalization".

[c] See description in Table 1 (main text).

[d] Includes transfer to another hospital, to a rehabilitation center, to a palliative care center, or discharge against medical advice.

[e] In the year before admission.

[f] In the 90 days before admission.

[g] Charlson comorbidities using ICD-10 codes by Quan et al[47] and ascertained using the discharge abstract of index hospitalization and of those in the year before discharge.

**Table S5. Characteristics of hospitalizations between July 2017 and July 2018 (Testing cohort)[a]**

| | Overall (n=26 291) | Vital status one year after admission | |
|---|---|---|---|
| | | Dead (n=3 968) | Alive (n=22 323) |
| Age, median (IQR), y | 65 (48-76) | 75 (65-85) | 63 (43-74) |
| Sex | | | |
| Male | 12 176 (46) | 2 170 (55) | 10 006 (45) |
| Female | 14 115 (54) | 1 798 (45) | 12 317 (55) |
| Hospital site | | | |
| A | 17 054 (65) | 2 441 (62) | 14 613 (65) |
| B | 9 237 (35) | 1 527 (38) | 7 710 (35) |
| Service type | | | |
| Medical | 12 721 (48) | 2 952 (74) | 9 769 (44) |
| Surgical | 9 773 (37) | 811 (20) | 8 962 (40) |
| Critical care[b] | 621 (2) | 204 (5) | 417 (2) |
| Obstetrics | 3 176 (12) | 1 (0) | 3 175 (14) |
| Admission type | | | |
| Non-elective | 17 733 (67) | 3 613 (91) | 14 120 (63) |
| Elective | 5 607 (21) | 354 (9) | 5 253 (24) |
| Obstetrics | 2 951 (11) | 1 (0) | 2 950 (13) |
| Real-time–accessible living status[c] | | | |
| Home | 12 852 (49) | 2 300 (58) | 10 552 (47) |
| Nursing home | 1 293 (5) | 451 (11) | 842 (4) |
| Chronic care hospital | 297 (1) | 111 (3) | 186 (1) |
| Unknown | 11 849 (45) | 1 106 (28) | 10 743 (48) |
| Living status at discharge | | | |
| Home | 12 452 (47) | 1 005 (25) | 11 447 (51) |
| Home with CLSC liaison | 8 878 (34) | 899 (23) | 7 979 (36) |
| Short term transitional care | 1 410 (5) | 306 (8) | 1 104 (5) |
| Nursing home | 1 622 (6) | 355 (9) | 1 267 (6) |
| Chronic care hospital | 535 (2) | 227 (6) | 308 (1) |
| Other[d] | 339 (1) | 121 (3) | 218 (1) |
| Death | 1 055 (4) | 1 055 (27) | 0 (0) |
| ED visits[e] | | | |
| 0 | 15 827 (60) | 1 646 (41) | 14 181 (64) |
| 1-2 | 7 471 (28) | 1 396 (35) | 6 075 (27) |
| 3 or more | 2 993 (11) | 926 (23) | 2 067 (9) |
| Admissions by ambulance[e] | | | |
| 0 | 22 695 (86) | 2 759 (70) | 19 936 (89) |
| 1-2 | 2 986 (11) | 955 (24) | 2 031 (9) |
| 3 or more | 610 (2) | 254 (6) | 356 (2) |
| Weeks recently hospitalized[f] | | | |
| 0 | 22 664 (86) | 2 720 (69) | 19 944 (89) |
| 1-2 | 2 775 (11) | 896 (23) | 1 879 (8) |

| | Overall (n=26 291) | Vital status one year after admission | |
|---|---|---|---|
| | | Dead (n=3 968) | Alive (n=22 323) |
| 3 or more | 852 (3) | 352 (9) | 500 (2) |
| Past discharge abstract accessible[c] | 14 410 (55) | 2 932 (74) | 11 478 (51) |
| Admission during flu season[c] | 6 517 (25) | 1 061 (27) | 5 456 (24) |
| ED admission | 13 268 (50) | 2 814 (71) | 10 454 (47) |
| Ambulance admission | 7 592 (29) | 2 005 (51) | 5 587 (25) |
| Urgent 30-day readmission | 2 521 (10) | 851 (21) | 1 670 (7) |
| ICU admission | 1 082 (4) | 245 (6) | 837 (4) |
| ICU stay during hospitalization | 3 588 (14) | 719 (18) | 2 869 (13) |
| Length of stay, median (IQR), d | 3 (2-7) | 7 (3-14) | 3 (2-6) |
| Code status preference[g] | | | |
| Full code | 2 331 (9) | 285 (7) | 2 046 (9) |
| DNR/Intubation-OK | 937 (4) | 263 (7) | 674 (3) |
| DNR/DNI | 4 505 (17) | 2 159 (54) | 2 346 (11) |
| Not documented | 18 518 (70) | 1 261 (32) | 17 257 (77) |
| Major comorbidity[h] | | | |
| Congestive heart failure | 2 826 (11) | 960 (24) | 1 866 (8) |
| Chronic pulmonary disease | 4 834 (18) | 1 235 (31) | 3 599 (16) |
| Dementia | 1 836 (7) | 660 (17) | 1 176 (5) |
| Metastatic cancer | 1 891 (7) | 1 020 (26) | 871 (4) |

Abbreviations: CLSC, *Centre local de services communautaires*; DNI, do-not-intubate; DNR, do-not-resuscitate; ED, emergency department; EOL, end of life.

[a] Data given as number (percentage) of hospitalizations unless otherwise indicated. Percentages may not add to 100 due to rounding.

[b] Represent direct admissions to the ICU before a primary non–critical care service could be specified (i.e., the responsible service upon ICU discharge). ICU exposure is more precisely measured with the variables "ICU admission" and "ICU stay during hospitalization".

[c] See description in Table 1 (main text).

[d] Includes transfer to another hospital, to a rehabilitation center, to a palliative care center, or discharge against medical advice.

[e] In the year before admission.

[f] In the 90 days before admission.

[g] Last preference documented during hospitalization if one was documented.

[h] Charlson comorbidities using ICD-10 codes by Quan et al.[47] and ascertained using discharge abstract of index hospitalization and of those in the year before discharge.

**Table S6. Clinical utility of prediction models with a random hospitalization per patient**

| | RF-AdminDemoDx | mHOMR | RF-AdminDemo | RF-Minimal | Random |
|---|---|---|---|---|---|
| Threshold | 0.423 (0.417-0.431) | 0.414 (0.407-0.420) | 0.413 (0.405-0.418) | 0.398 (0.394-0.411) | 0.899 (0.892-0.903) |
| No. Alerts | 1649 (1637-1664) | 1649 (1638-1664) | 1649 (1637-1664) | 1651 (1641-1675) | 1649 (1638-1665) |
| eRD, % | 9.1 (8.0-10.3) | 5.7 (4.7-6.5) | 6.3 (5.3-7.4) | 5.4 (4.5-6.3) | 3.0 (2.3-3.7) |
| eRR | 1.13 (1.11-1.14) | 1.08 (1.06-1.09) | 1.09 (1.07-1.10) | 1.07 (1.06-1.09) | 1.04 (1.03-1.05) |
| Benefit | 0.1111 (0.1062-0.1155) | 0.1064 (0.1017-0.1105) | 0.1073 (0.1026-0.1115) | 0.1061 (0.1012-0.1104) | 0.1027 (0.0979-0.1071) |
| Harm | 0.1064 (0.1016-0.1111) | 0.1092 (0.1041-0.1139) | 0.1076 (0.1026-0.1124) | 0.1099 (0.1047-0.1147) | 0.1286 (0.1225-0.1344) |
| Net benefit | 0.0047 (0.0028-0.0067) | -0.0027 (-0.0048 to -0.0012) | -0.0004 (-0.0023-0.0016) | -0.0038 (-0.0058 to -0.0021) | -0.0258 (-0.0284 to -0.0236) |
| Standardized net benefit[a] | 0.0347 (0.0210-0.0492) | -0.0202 (-0.0356 to -0.0083) | -0.0027 (-0.0164-0.0113) | -0.0282 (-0.0423 to -0.0155) | -0.1895 (-0.2061 to -0.1738) |
| NNB, Alerts | 8.0 (7.1-9.3) | 12.9 (11.3-16.0) | 11.6 (10.1-14.0) | 13.5 (11.9-16.8) | 24.6 (20.0-33.1) |
| PPV, % | 56.9 (54.4-59.3) | 40.9 (38.4-43.0) | 44.2 (41.8-46.6) | 39.4 (37.1-41.6) | 13.9 (12.3-15.7) |
| NPV, % | 91.2 (90.7-91.7) | 89.4 (88.9-89.9) | 89.8 (89.3-90.3) | 89.2 (88.8-89.8) | 86.4 (85.9-87.0) |
| Sensitivity, % | 41.8 (40.1-43.3) | 30.0 (28.4-31.6) | 32.4 (30.8-34.2) | 28.9 (27.4-30.6) | 10.2 (9.0-11.4) |
| Specificity, % | 95.0 (94.7-95.3) | 93.2 (92.9-93.4) | 93.5 (93.3-93.8) | 93.0 (92.7-93.2) | 90.0 (89.8-90.2) |
| C-statistic[b] | 0.86 (0.85-0.87) | 0.80 (0.79-0.81) | 0.81 (0.80-0.82) | 0.79 (0.78-0.80) | 0.50 (0.49-0.51) |

Abbreviations: eRR, expected relative risk, eRD, expected risk difference; NNB, number needed to benefit; NPV, negative predictive value; PPV, positive predictive value.
Data presented as point estimate (95% CI). For all models: Sample size = 16490 patients; Threshold set such that *P*(Alert) = 10%; TP CSOs in usual care = 1627 (1546-1700); FP CSOs in usual care = 3614 (3507-3723); Observed exchange rate = 0.450 (0.424-0.474); Prevalence of cases at the EOL = 13.6% (13.1%-14.1%); Random alert rule included as a point of reference, not as a strategy under consideration.

[a] Calculated as the net benefit divided by the prevalence of EOL status (1-year mortality).

[b] Threshold independent.

**Table S7. Clinical utility of prediction models with the first hospitalization per patient**

| | RF-AdminDemoDx | mHOMR | RF-AdminDemo | RF-Minimal | Random |
|---|---|---|---|---|---|
| Threshold | 0.403 (0.395-0.410) | 0.391 (0.385-0.398) | 0.385 (0.379-0.391) | 0.394 (0.391-0.404) | 0.899 (0.894-0.905) |
| No. Alerts | 1649 (1637-1665) | 1649 (1637-1664) | 1649 (1637-1665) | 1661 (1642-1676) | 1649 (1638-1664) |
| eRD, % | 10.3 (9.1-11.5) | 6.2 (5.2-7.2) | 6.4 (5.3-7.4) | 6.3 (5.3-7.3) | 3.3 (2.7-4.1) |
| eRR | 1.15 (1.13-1.17) | 1.09 (1.08-1.11) | 1.09 (1.08-1.11) | 1.09 (1.08-1.11) | 1.05 (1.04-1.06) |
| Benefit | 0.1045 (0.1002-0.1090) | 0.0992 (0.0949-0.1036) | 0.0995 (0.0949-0.1037) | 0.0993 (0.0949-0.1036) | 0.0954 (0.0913-0.0999) |
| Harm | 0.0987 (0.0945-0.1036) | 0.1006 (0.0964-0.1058) | 0.0991 (0.0950-0.1042) | 0.1011 (0.0967-0.1061) | 0.1188 (0.1134-0.1247) |
| Net benefit | 0.0059 (0.0038-0.0079) | -0.0014 (-0.0034-0.0002) | 0.0003 (-0.0016-0.0019) | -0.0018 (-0.0038 to -0.0001) | -0.0234 (-0.0257 to -0.0212) |
| Standardized net benefit[a] | 0.0445 (0.0288-0.0592) | -0.0109 (-0.0255-0.0014) | 0.0024 (-0.0120-0.0143) | -0.0135 (-0.0291 to -0.0008) | -0.1766 (-0.1926 to -0.1615) |
| NNB, Alerts | 7.4 (6.6-8.3) | 12.2 (10.5-14.5) | 11.8 (10.3-14.2) | 12.0 (10.5-14.6) | 22.6 (18.3-28.8) |
| PPV, % | 53.1 (50.5-55.6) | 39.1 (36.6-41.5) | 40.8 (38.1-43.0) | 38.2 (35.8-40.4) | 12.8 (11.2-14.6) |
| NPV, % | 91.2 (90.7-91.6) | 89.6 (89.1-90.1) | 89.8 (89.3-90.3) | 89.5 (89.0-90.0) | 86.7 (86.2-87.3) |
| Sensitivity, % | 40.1 (38.4-41.8) | 29.5 (27.8-31.1) | 30.8 (29.0-32.4) | 29.0 (27.2-30.5) | 9.7 (8.5-10.9) |
| Specificity, % | 94.6 (94.3-94.9) | 93.0 (92.7-93.2) | 93.2 (92.9-93.4) | 92.8 (92.6-93.1) | 89.9 (89.8-90.1) |
| C-statistic[b] | 0.85 (0.84-0.86) | 0.79 (0.78-0.80) | 0.80 (0.79-0.81) | 0.79 (0.78-0.80) | 0.49 (0.48-0.51) |

Abbreviations: eRR, expected relative risk, eRD, expected risk difference; NNB, number needed to benefit; NPV, negative predictive value; PPV, positive predictive value.

Data presented as point estimate (95% CI). For all models: Sample size = 16490 patients; Threshold set such that *P*(Alert) = 10%; TP CSOs in usual care = 1500 (1436-1574); FP CSOs in usual care = 3653 (3539-3765); Observed exchange rate = 0.411 (0.388-0.437); Prevalence of cases at the EOL = 13.2% (12.8%-13.8%); Random alert rule included as a point of reference, not as a strategy under consideration.

[a] Calculated as the net benefit divided by the prevalence of EOL status (1-year mortality).

[b] Threshold independent.

**Table S8. Clinical utility of prediction models with the last hospitalization per patient**

| | RF-AdminDemoDx | mHOMR | RF-AdminDemo | RF-Minimal | Random |
|---|---|---|---|---|---|
| Threshold | 0.439 (0.432-0.446) | 0.432 (0.425-0.438) | 0.433 (0.427-0.439) | 0.399 (0.394-0.410) | 0.899 (0.893-0.904) |
| No. Alerts | 1649 (1637-1665) | 1649 (1638-1665) | 1649 (1638-1665) | 1651 (1639-1672) | 1649 (1637-1664) |
| eRD, % | 7.4 (6.4-8.5) | 4.7 (3.8-5.6) | 5.3 (4.5-6.4) | 4.6 (3.8-5.4) | 2.4 (1.8-3.0) |
| eRR | 1.10 (1.08-1.11) | 1.06 (1.05-1.07) | 1.07 (1.06-1.08) | 1.06 (1.05-1.07) | 1.03 (1.02-1.04) |
| Benefit | 0.1178 (0.1135-0.1230) | 0.1140 (0.1094-0.1187) | 0.1149 (0.1106-0.1197) | 0.1139 (0.1093-0.1190) | 0.1108 (0.1062-0.1156) |
| Harm | 0.1151 (0.1102-0.1203) | 0.1180 (0.1132-0.1235) | 0.1169 (0.1119-0.1222) | 0.1189 (0.1138-0.1241) | 0.1394 (0.1331-0.1460) |
| Net benefit | 0.0027 (0.0008-0.0047) | -0.0040 (-0.0059 to -0.0021) | -0.0019 (-0.0036 to -0.0001) | -0.0049 (-0.0068 to -0.0031) | -0.0286 (-0.0312 to -0.0260) |
| Standardized net benefit[a] | 0.0194 (0.0056-0.0327) | -0.0287 (-0.0416 to -0.0150) | -0.0138 (-0.0258 to -0.0007) | -0.0350 (-0.0483 to -0.0223) | -0.2037 (-0.2190 to -0.1879) |
| NNB, Alerts | 9.6 (8.5-11.4) | 15.3 (13.0-19.1) | 13.4 (11.3-16.1) | 15.4 (13.2-19.1) | 30.0 (24.3-42.3) |
| PPV, % | 61.1 (58.6-63.7) | 44.5 (42.2-47.0) | 47.8 (45.6-50.4) | 42.3 (39.7-44.5) | 13.9 (12.3-15.7) |
| NPV, % | 91.2 (90.7-91.6) | 89.3 (88.8-89.8) | 89.7 (89.2-90.2) | 89.1 (88.6-89.5) | 85.9 (85.3-86.5) |
| Sensitivity, % | 43.5 (41.9-45.1) | 31.7 (30.2-33.1) | 34.0 (32.6-35.6) | 30.1 (28.5-31.5) | 9.9 (8.8-11.1) |
| Specificity, % | 95.5 (95.2-95.8) | 93.5 (93.3-93.8) | 93.9 (93.7-94.2) | 93.3 (93.0-93.5) | 90.0 (89.8-90.2) |
| C-statistic[b] | 0.87 (0.86-0.88) | 0.81 (0.80-0.82) | 0.82 (0.82-0.83) | 0.80 (0.79-0.81) | 0.50 (0.48-0.51) |

Abbreviations: eRR, expected relative risk, eRD, expected risk difference; NNB, number needed to benefit; NPV, negative predictive value; PPV, positive predictive value.

Data presented as point estimate (95% CI). For all models: Sample size = 16490 patients; Threshold set such that *P*(Alert) = 10%; TP CSOs in usual care = 1772 (1698-1853); FP CSOs in usual care = 3626 (3512-3735); Observed exchange rate = 0.489 (0.461-0.518); Prevalence of cases at the EOL = 14.1% (13.6%-14.6%); Random alert rule included as a point of reference, not as a strategy under consideration.

[a] Calculated as the net benefit divided by the prevalence of EOL status (1-year mortality).

[b] Threshold independent.

**Figure S1. Calibration plot of prediction models in testing cohort**

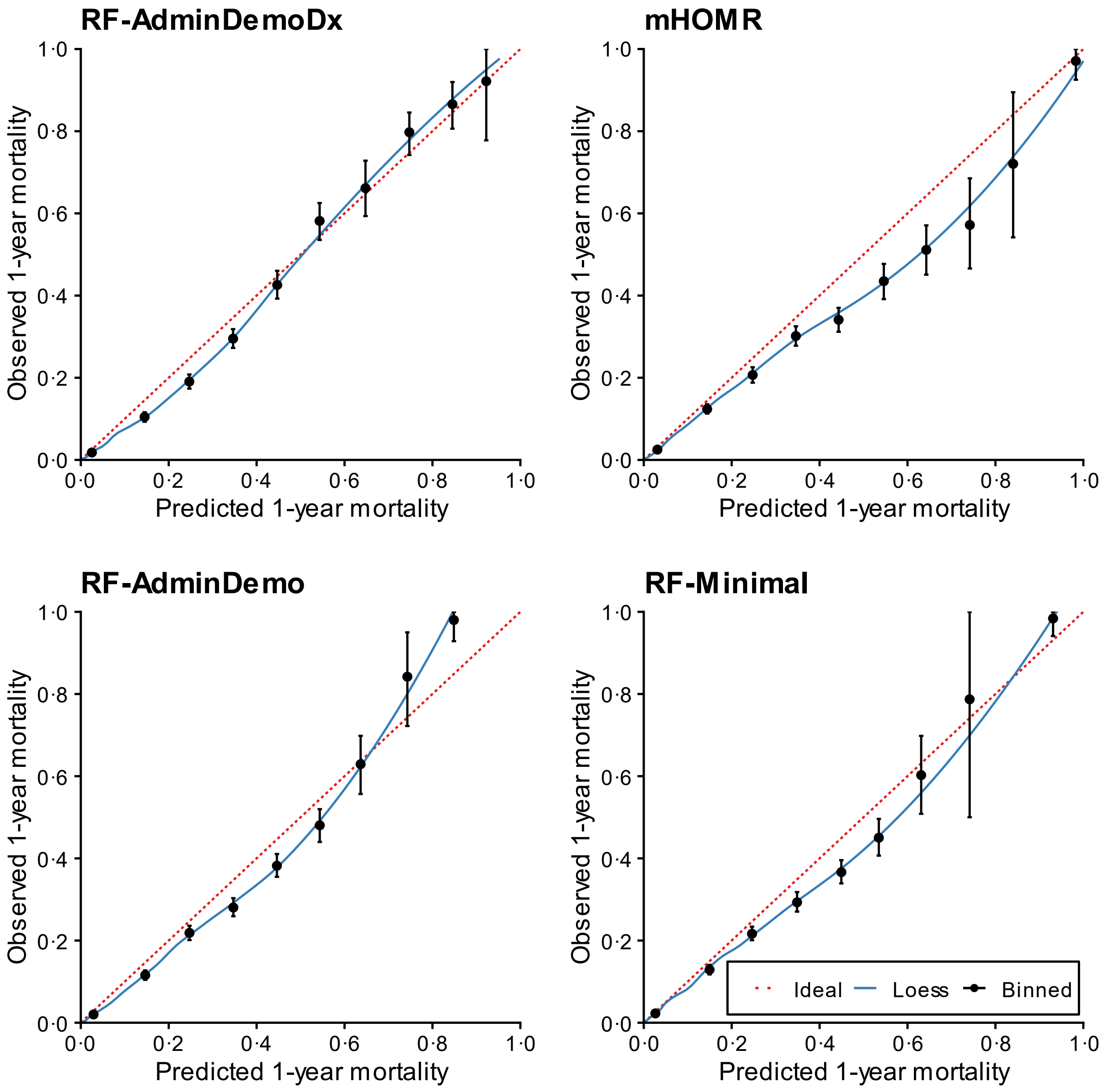

Proportion with the outcome (y-axis) in ten binned groups (x-axis). Bins with less than five observations removed. Binned estimates generated with 1 000 two-stage bootstrap resamples of the testing cohort; mean predicted probability and mean observed proportion over bootstraps plotted on x- and y-axis, respectively. Error bars (95% CI) plotted using corresponding percentiles for bootstraps in each bin. Non-parametric curve generated by selecting one random hospitalization per patient and applying the loess algorithm (*span* set to 0.4) to individual observed outcome (0 or 1) vs. individual predicted probability.

**Figure S2. Forest plot of clinical utility in relevant subgroups of hospitalized patients (RF-AdminDemoDx)**

**RF-AdminDemoDx**

| | **E[CSO\|EOL] Usual care (%)** | **E[CSO\|EOL] CDSS (%)** | **Alerts (NNB)** | **eRR (95% CI)** |
|---|---|---|---|---|
| **Age** | | | | |
| Age < 65 y | 437/876 (50) | 634/876 (72) | 618 (3.1) | 1.45 (1.38-1.53) |
| Age ≥ 65 y | 2162/2897 (75) | 2370/2897 (82) | 1586 (7.6) | 1.10 (1.08-1.11) |
| **Sex** | | | | |
| Female | 1212/1721 (70) | 1397/1721 (81) | 1039 (5.6) | 1.15 (1.13-1.18) |
| Male | 1387/2052 (68) | 1607/2052 (78) | 1165 (5.3) | 1.16 (1.14-1.18) |
| **Hospital site** | | | | |
| A | 1455/2364 (62) | 1798/2364 (76) | 1523 (4.4) | 1.24 (1.21-1.26) |
| B | 1144/1409 (81) | 1206/1409 (86) | 681 (11.0) | 1.05 (1.04-1.07) |
| **Admission type** | | | | |
| Elective | 74/331 (22) | 157/331 (47) | 169 (2.0) | 2.12 (1.85-2.62) |
| Non-elective | 2525/3442 (73) | 2847/3442 (83) | 2035 (6.3) | 1.13 (1.11-1.14) |
| **Service type** | | | | |
| Medical | 2135/2793 (76) | 2350/2793 (84) | 1662 (7.7) | 1.10 (1.09-1.12) |
| Surgical | 312/796 (39) | 495/796 (62) | 435 (2.4) | 1.59 (1.49-1.71) |
| **ICU stay** | | | | |
| Yes | 538/690 (78) | 573/690 (83) | 286 (8.2) | 1.07 (1.04-1.09) |
| No | 2061/3083 (67) | 2431/3083 (79) | 1918 (5.2) | 1.18 (1.16-1.20) |
| **Length of stay** | | | | |
| 1-2 days | 354/708 (50) | 500/708 (71) | 424 (2.9) | 1.41 (1.34-1.50) |
| 3-6 days | 698/1143 (61) | 841/1143 (74) | 701 (4.9) | 1.20 (1.17-1.24) |
| ≥ 7 days | 1547/1922 (80) | 1663/1922 (87) | 1079 (9.3) | 1.07 (1.06-1.09) |
| **Comorbidity** | | | | |
| Chronic pulmonary disease | 922/1187 (78) | 990/1187 (83) | 741 (10.9) | 1.07 (1.06-1.09) |
| Congestive heart failure | 745/939 (79) | 786/939 (84) | 525 (12.8) | 1.06 (1.04-1.07) |
| Dementia | 570/647 (88) | 592/647 (91) | 295 (13.4) | 1.04 (1.02-1.06) |
| Metastatic cancer | 634/938 (68) | 795/938 (85) | 785 (4.9) | 1.25 (1.21-1.30) |
| Any of the above | 1999/2657 (75) | 2226/2657 (84) | 1601 (7.1) | 1.11 (1.10-1.13) |
| None of the above | 600/1116 (54) | 778/1116 (70) | 603 (3.4) | 1.30 (1.25-1.35) |
| **Overall** | **2599/3773 (69)** | **3004/3773 (80)** | **2204 (5.4)** | **1.16 (1.14-1.17)** |

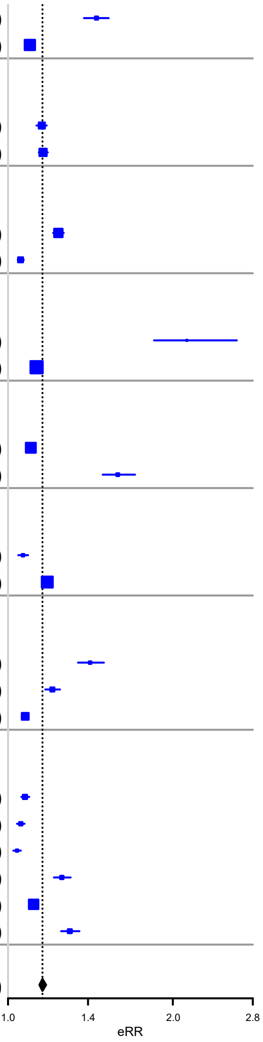

Abbreviations: CDSS, clinical decision support system; CSO, code status order; EOL, end of life; NNB, number needed to benefit.
The term E[CSO | EOL] is the expected number of CSOs among patients at the EOL for a given strategy using decision tree 2 and is presented as n(CSO & EOL)/n(EOL). The difference in this proportion between "Usual care" and "CDSS" is the eRD. Horizontal error bars (95% CI) generated with 1000 bootstrap resamples of hospitalizations in each subgroup.

**Figure S3. Forest plot of clinical utility in relevant subgroups of hospitalized patients (mHOMR)**

**mHOMR**

| | E[CSO\|EOL] Usual care (%) | E[CSO\|EOL] CDSS (%) | Alerts (NNB) | eRR (95% CI) |
|---|---|---|---|---|
| **Age** | | | | |
| Age < 65 y | 437/876 (50) | 467/876 (53) | 163 (5.4) | 1.07 (1.05-1.10) |
| Age ≥ 65 y | 2162/2897 (75) | 2338/2897 (81) | 2041 (11.6) | 1.08 (1.07-1.10) |
| **Sex** | | | | |
| Female | 1212/1721 (70) | 1282/1721 (74) | 832 (11.9) | 1.06 (1.05-1.07) |
| Male | 1387/2052 (68) | 1523/2052 (74) | 1372 (10.1) | 1.10 (1.08-1.12) |
| **Hospital site** | | | | |
| A | 1455/2364 (62) | 1636/2364 (69) | 1576 (8.7) | 1.12 (1.11-1.14) |
| B | 1144/1409 (81) | 1169/1409 (83) | 628 (25.1) | 1.02 (1.01-1.03) |
| **Admission type** | | | | |
| Elective | 74/331 (22) | 94/331 (28) | 117 (5.8) | 1.27 (1.16-1.43) |
| Non-elective | 2525/3442 (73) | 2711/3442 (79) | 2087 (11.2) | 1.07 (1.06-1.09) |
| **Service type** | | | | |
| Medical | 2135/2793 (76) | 2300/2793 (82) | 1812 (11.0) | 1.08 (1.07-1.09) |
| Surgical | 312/796 (39) | 346/796 (43) | 194 (5.7) | 1.11 (1.08-1.15) |
| **ICU stay** | | | | |
| Yes | 538/690 (78) | 552/690 (80) | 372 (26.6) | 1.03 (1.01-1.04) |
| No | 2061/3083 (67) | 2253/3083 (73) | 1832 (9.5) | 1.09 (1.08-1.11) |
| **Length of stay** | | | | |
| 1-2 days | 354/708 (50) | 415/708 (59) | 366 (6.0) | 1.17 (1.13-1.22) |
| 3-6 days | 698/1143 (61) | 771/1143 (67) | 747 (10.2) | 1.10 (1.08-1.13) |
| ≥ 7 days | 1547/1922 (80) | 1619/1922 (84) | 1091 (15.2) | 1.05 (1.04-1.06) |
| **Comorbidity** | | | | |
| Chronic pulmonary disease | 922/1187 (78) | 979/1187 (82) | 918 (16.1) | 1.06 (1.05-1.08) |
| Congestive heart failure | 745/939 (79) | 775/939 (83) | 518 (17.3) | 1.04 (1.03-1.06) |
| Dementia | 570/647 (88) | 592/647 (91) | 384 (17.5) | 1.04 (1.02-1.06) |
| Metastatic cancer | 634/938 (68) | 700/938 (75) | 376 (5.7) | 1.10 (1.08-1.13) |
| Any of the above | 1999/2657 (75) | 2133/2657 (80) | 1554 (11.6) | 1.07 (1.06-1.08) |
| None of the above | 600/1116 (54) | 672/1116 (60) | 650 (9.0) | 1.12 (1.09-1.15) |
| **Overall** | **2599/3773 (69)** | **2805/3773 (74)** | **2204 (10.7)** | **1.08 (1.07-1.09)** |

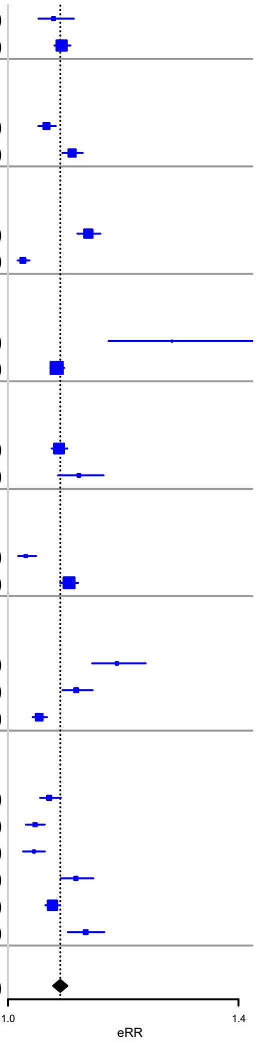

See caption for Figure S2.

**Figure S4. Forest plot of clinical utility in relevant subgroups of hospitalized patients (RF-AdminDemo)**

**RF-AdminDemo**

| | E[CSO\|EOL] Usual care (%) | E[CSO\|EOL] CDSS (%) | Alerts (NNB) | eRR (95% CI) |
|---|---|---|---|---|
| **Age** | | | | |
| Age < 65 y | 437/876 (50) | 540/876 (62) | 397 (3.9) | 1.24 (1.19-1.29) |
| Age ≥ 65 y | 2162/2897 (75) | 2321/2897 (80) | 1807 (11.4) | 1.07 (1.06-1.09) |
| **Sex** | | | | |
| Female | 1212/1721 (70) | 1311/1721 (76) | 837 (8.5) | 1.08 (1.07-1.10) |
| Male | 1387/2052 (68) | 1550/2052 (76) | 1367 (8.4) | 1.12 (1.10-1.14) |
| **Hospital site** | | | | |
| A | 1455/2364 (62) | 1698/2364 (72) | 1604 (6.6) | 1.17 (1.14-1.19) |
| B | 1144/1409 (81) | 1163/1409 (83) | 600 (31.6) | 1.02 (1.01-1.03) |
| **Admission type** | | | | |
| Elective | 74/331 (22) | 119/331 (36) | 115 (2.6) | 1.61 (1.41-1.88) |
| Non-elective | 2525/3442 (73) | 2742/3442 (80) | 2089 (9.6) | 1.09 (1.07-1.10) |
| **Service type** | | | | |
| Medical | 2135/2793 (76) | 2297/2793 (82) | 1748 (10.8) | 1.08 (1.07-1.09) |
| Surgical | 312/796 (39) | 407/796 (51) | 286 (3.0) | 1.30 (1.24-1.38) |
| **ICU stay** | | | | |
| Yes | 538/690 (78) | 555/690 (80) | 340 (20.0) | 1.03 (1.02-1.05) |
| No | 2061/3083 (67) | 2306/3083 (75) | 1864 (7.6) | 1.12 (1.10-1.14) |
| **Length of stay** | | | | |
| 1-2 days | 354/708 (50) | 437/708 (62) | 374 (4.5) | 1.23 (1.18-1.30) |
| 3-6 days | 698/1143 (61) | 804/1143 (70) | 749 (7.1) | 1.15 (1.12-1.19) |
| ≥ 7 days | 1547/1922 (80) | 1620/1922 (84) | 1081 (14.8) | 1.05 (1.04-1.06) |
| **Comorbidity** | | | | |
| Chronic pulmonary disease | 922/1187 (78) | 989/1187 (83) | 880 (13.1) | 1.07 (1.06-1.09) |
| Congestive heart failure | 745/939 (79) | 779/939 (83) | 530 (15.6) | 1.05 (1.03-1.06) |
| Dementia | 570/647 (88) | 590/647 (91) | 405 (20.2) | 1.04 (1.02-1.05) |
| Metastatic cancer | 634/938 (68) | 703/938 (75) | 388 (5.6) | 1.11 (1.08-1.14) |
| Any of the above | 1999/2657 (75) | 2145/2657 (81) | 1533 (10.5) | 1.07 (1.06-1.09) |
| None of the above | 600/1116 (54) | 716/1116 (64) | 671 (5.8) | 1.19 (1.15-1.23) |
| **Overall** | **2599/3773 (69)** | **2861/3773 (76)** | **2204 (8.4)** | **1.10 (1.09-1.11)** |

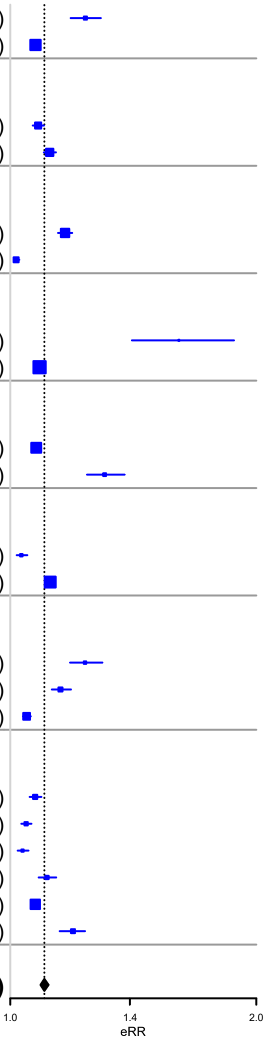

See caption for Figure S2.

**Figure S5. Forest plot of clinical utility in relevant subgroups of hospitalized patients (RF-Minimal)**

**RF-Minimal**

| | E[CSO\|EOL] Usual care (%) | E[CSO\|EOL] CDSS (%) | Alerts (NNB) | eRR (95% CI) |
|---|---|---|---|---|
| **Age** | | | | |
| Age < 65 y | 437/876 (50) | 473/876 (54) | 233 (6.5) | 1.08 (1.06-1.11) |
| Age ≥ 65 y | 2162/2897 (75) | 2329/2897 (80) | 1972 (11.8) | 1.08 (1.07-1.09) |
| **Sex** | | | | |
| Female | 1212/1721 (70) | 1282/1721 (74) | 768 (11.0) | 1.06 (1.04-1.07) |
| Male | 1387/2052 (68) | 1520/2052 (74) | 1437 (10.8) | 1.10 (1.08-1.11) |
| **Hospital site** | | | | |
| A | 1455/2364 (62) | 1654/2364 (70) | 1749 (8.8) | 1.14 (1.12-1.16) |
| B | 1144/1409 (81) | 1148/1409 (81) | 456 (114.0) | 1.00 (1.00-1.01) |
| **Admission type** | | | | |
| Elective | 74/331 (22) | 79/331 (24) | 31 (6.2) | 1.07 (1.02-1.14) |
| Non-elective | 2525/3442 (73) | 2723/3442 (79) | 2174 (11.0) | 1.08 (1.07-1.09) |
| **Service type** | | | | |
| Medical | 2135/2793 (76) | 2293/2793 (82) | 1816 (11.5) | 1.07 (1.06-1.09) |
| Surgical | 312/796 (39) | 351/796 (44) | 219 (5.6) | 1.12 (1.09-1.17) |
| **ICU stay** | | | | |
| Yes | 538/690 (78) | 548/690 (79) | 346 (34.6) | 1.02 (1.01-1.03) |
| No | 2061/3083 (67) | 2254/3083 (73) | 1859 (9.6) | 1.09 (1.08-1.11) |
| **Length of stay** | | | | |
| 1-2 days | 354/708 (50) | 414/708 (58) | 388 (6.5) | 1.17 (1.13-1.22) |
| 3-6 days | 698/1143 (61) | 769/1143 (67) | 753 (10.6) | 1.10 (1.08-1.13) |
| ≥ 7 days | 1547/1922 (80) | 1619/1922 (84) | 1064 (14.8) | 1.05 (1.04-1.06) |
| **Comorbidity** | | | | |
| Chronic pulmonary disease | 922/1187 (78) | 977/1187 (82) | 834 (15.2) | 1.06 (1.05-1.08) |
| Congestive heart failure | 745/939 (79) | 764/939 (81) | 375 (19.7) | 1.03 (1.01-1.04) |
| Dementia | 570/647 (88) | 583/647 (90) | 316 (24.3) | 1.02 (1.01-1.04) |
| Metastatic cancer | 634/938 (68) | 710/938 (76) | 399 (5.2) | 1.12 (1.09-1.15) |
| Any of the above | 1999/2657 (75) | 2126/2657 (80) | 1431 (11.3) | 1.06 (1.05-1.08) |
| None of the above | 600/1116 (54) | 676/1116 (61) | 774 (10.2) | 1.13 (1.10-1.16) |
| **Overall** | **2599/3773 (69)** | **2802/3773 (74)** | **2205 (10.9)** | **1.08 (1.07-1.09)** |

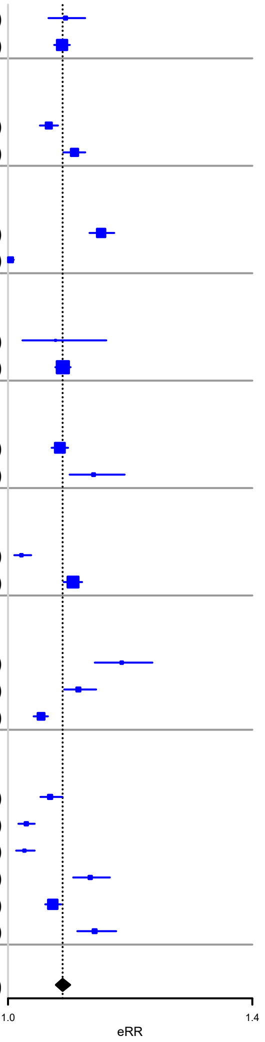

See caption for Figure S2.

**Figure S6. Forest plot of clinical utility in relevant subgroups of hospitalized patients (Random)**

**Random**

| | E[CSO\|EOL] Usual care (%) | E[CSO\|EOL] CDSS (%) | Alerts (NNB) | eRR (95% CI) |
|---|---|---|---|---|
| **Age** | | | | |
| Age < 65 y | 437/876 (50) | 477/876 (54) | 894 (22.4) | 1.09 (1.06-1.13) |
| Age ≥ 65 y | 2162/2897 (75) | 2247/2897 (78) | 1310 (15.4) | 1.04 (1.03-1.05) |
| **Sex** | | | | |
| Female | 1212/1721 (70) | 1261/1721 (73) | 1054 (21.5) | 1.04 (1.03-1.05) |
| Male | 1387/2052 (68) | 1463/2052 (71) | 1150 (15.1) | 1.05 (1.04-1.07) |
| **Hospital site** | | | | |
| A | 1455/2364 (62) | 1545/2364 (65) | 1385 (15.4) | 1.06 (1.05-1.08) |
| B | 1144/1409 (81) | 1179/1409 (84) | 819 (23.4) | 1.03 (1.02-1.04) |
| **Admission type** | | | | |
| Elective | 74/331 (22) | 104/331 (31) | 535 (17.8) | 1.41 (1.27-1.63) |
| Non-elective | 2525/3442 (73) | 2620/3442 (76) | 1669 (17.6) | 1.04 (1.03-1.05) |
| **Service type** | | | | |
| Medical | 2135/2793 (76) | 2207/2793 (79) | 1206 (16.8) | 1.03 (1.03-1.04) |
| Surgical | 312/796 (39) | 361/796 (45) | 945 (19.3) | 1.16 (1.12-1.22) |
| **ICU stay** | | | | |
| Yes | 538/690 (78) | 557/690 (81) | 344 (18.1) | 1.04 (1.02-1.05) |
| No | 2061/3083 (67) | 2167/3083 (70) | 1860 (17.5) | 1.05 (1.04-1.06) |
| **Length of stay** | | | | |
| 1-2 days | 354/708 (50) | 393/708 (56) | 801 (20.5) | 1.11 (1.08-1.15) |
| 3-6 days | 698/1143 (61) | 740/1143 (65) | 700 (16.7) | 1.06 (1.04-1.08) |
| ≥ 7 days | 1547/1922 (80) | 1591/1922 (83) | 703 (16.0) | 1.03 (1.02-1.04) |
| **Comorbidity** | | | | |
| Chronic pulmonary disease | 922/1187 (78) | 944/1187 (80) | 466 (21.2) | 1.02 (1.01-1.03) |
| Congestive heart failure | 745/939 (79) | 772/939 (82) | 294 (10.9) | 1.04 (1.02-1.05) |
| Dementia | 570/647 (88) | 575/647 (89) | 173 (34.6) | 1.01 (1.00-1.02) |
| Metastatic cancer | 634/938 (68) | 670/938 (71) | 177 (4.9) | 1.06 (1.04-1.08) |
| Any of the above | 1999/2657 (75) | 2071/2657 (78) | 874 (12.1) | 1.04 (1.03-1.05) |
| None of the above | 600/1116 (54) | 653/1116 (59) | 1330 (25.1) | 1.09 (1.07-1.12) |
| **Overall** | **2599/3773 (69)** | **2724/3773 (72)** | **2204 (17.6)** | **1.05 (1.04-1.06)** |

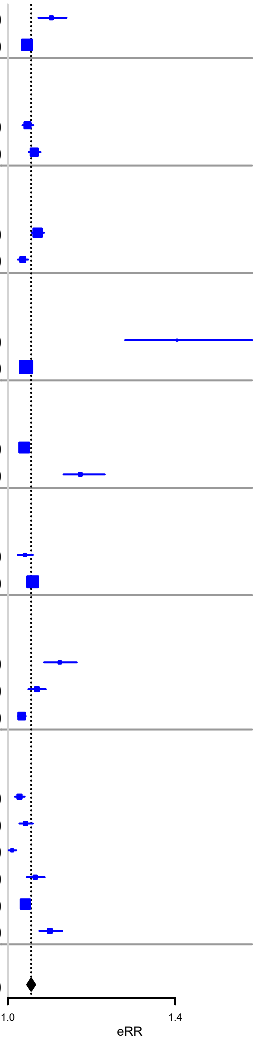

See caption for Figure S2.

**Figure S7. Decision curve analysis with a random hospitalization per patient**

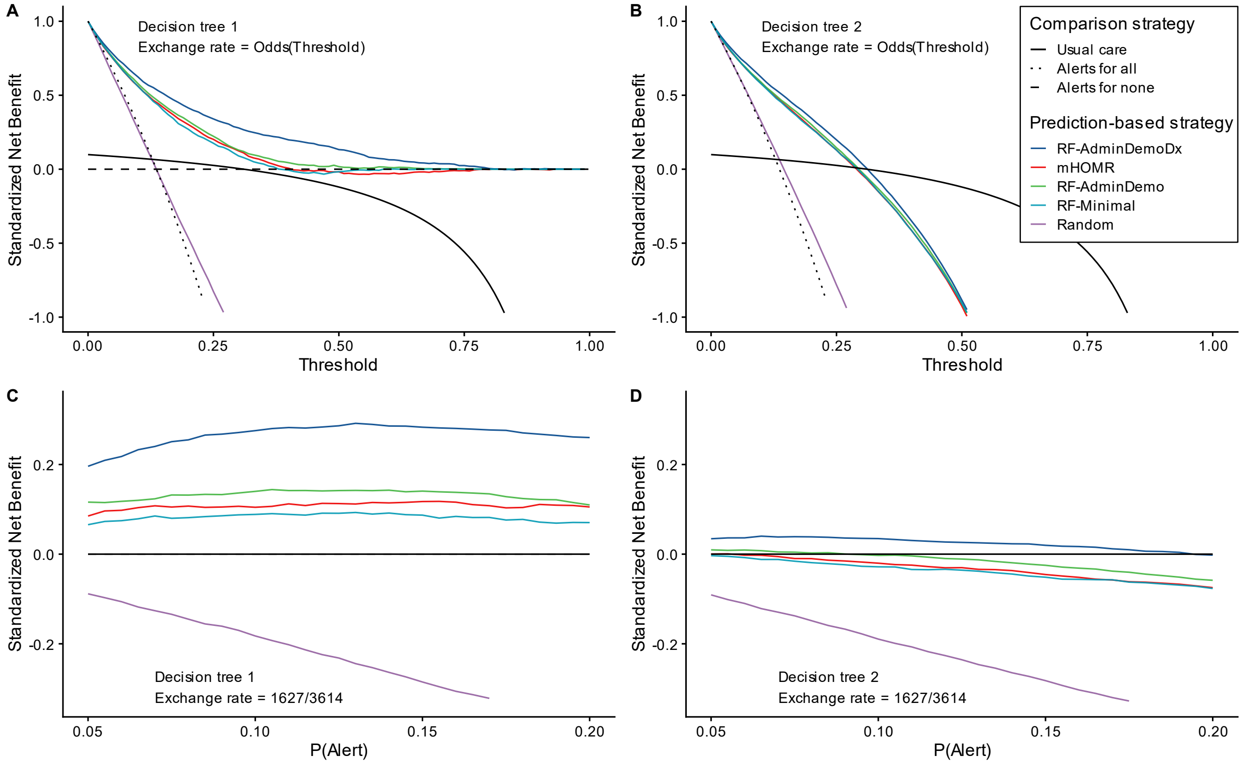

Exchange rate in bottom panels calculated using observed TP and FP rates in included sample (n=16940).

**Figure S8. Decision curve analysis with the first hospitalization per patient**

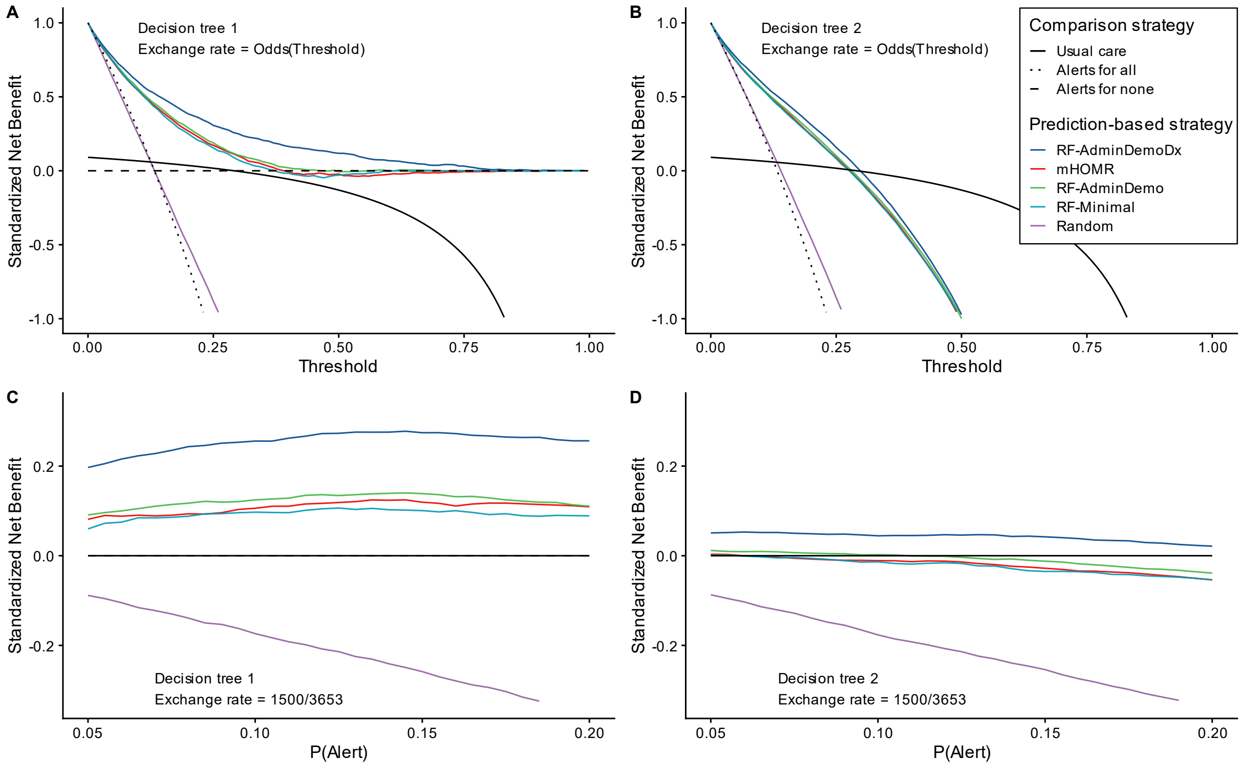

Exchange rate in bottom panels calculated using observed TP and FP rates in included sample (n=16940).

**Figure S9. Decision curve analysis with the last hospitalization per patient**

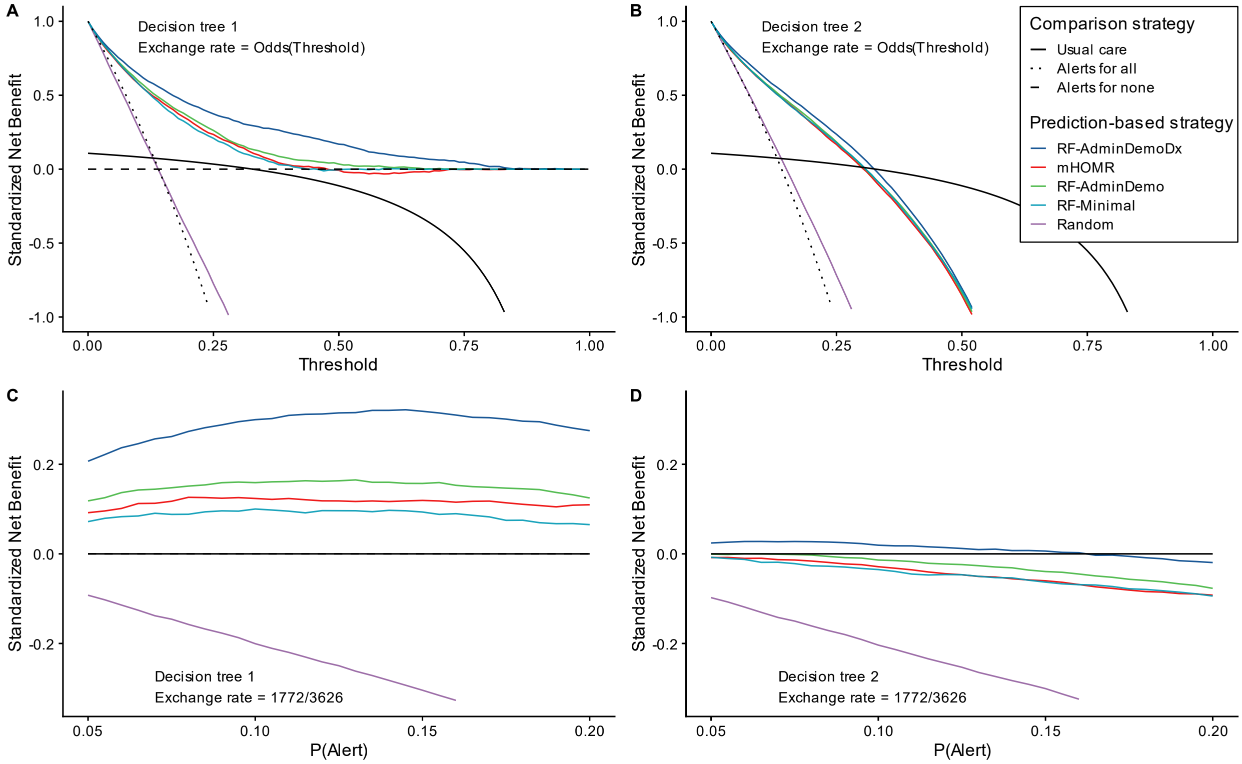

Exchange rate in bottom panels calculated using observed TP and FP rates in included sample (n=16940).

**References**

1. Steyerberg EW, Vickers AJ, Cook NR, et al. Assessing the performance of prediction models: a framework for some traditional and novel measures. *Epidemiol Camb Mass*. 2010;21(1):128-138. doi:10.1097/EDE.0b013e3181c30fb2

2. Steyerberg EW. Evaluation of Clinical Usefulness. *Clin Predict Models*. Published online 2019:309-328. doi:10.1007/978-3-030-16399-0_16

3. Wolff RF, Moons KGM, Riley RD, et al. PROBAST: A Tool to Assess the Risk of Bias and Applicability of Prediction Model Studies. *Ann Intern Med*. 2019;170(1):51-58. doi:10.7326/M18-1376

4. Huber MT, Highland JD, Krishnamoorthi VR, Tang JW-Y. Utilizing the Electronic Health Record to Improve Advance Care Planning: A Systematic Review. *Am J Hosp Palliat Med*. 2018;35(3):532-541. doi:10.1177/1049909117715217

5. Bush RA, Pérez A, Baum T, Etland C, Connelly CD. A systematic review of the use of the electronic health record for patient identification, communication, and clinical support in palliative care. *JAMIA Open*. 2018;1(2):294-303. doi:10.1093/jamiaopen/ooy028

6. Rhodes RL, Kazi S, Xuan L, Amarasingham R, Halm EA. Initial Development of a Computer Algorithm to Identify Patients With Breast and Lung Cancer Having Poor Prognosis in a Safety Net Hospital. *Am J Hosp Palliat Care*. 2016;33(7):678-683. doi:10.1177/1049909115591499

7. Mason B, Boyd K, Murray SA, et al. Developing a computerised search to help UK General Practices identify more patients for palliative care planning: a feasibility study. *BMC Fam Pract*. 2015;16:99. doi:10.1186/s12875-015-0312-z

8. Cox CE, Jones DM, Reagan W, et al. Palliative Care Planner: A Pilot Study to Evaluate Acceptability and Usability of an Electronic Health Records System-integrated, Needs-targeted App Platform. *Ann Am Thorac Soc*. 2018;15(1):59-68. doi:10.1513/AnnalsATS.201706-500OC

9. Ernecoff NC, Wessell KL, Gabriel S, Carey TS, Hanson LC. A Novel Screening Method to Identify Late-Stage Dementia Patients for Palliative Care Research and Practice. *J Pain Symptom Manage*. 2018;55(4):1152-1158.e1. doi:10.1016/j.jpainsymman.2017.12.480

10. Tan A, Durbin M, Chung FR, et al. Design and implementation of a clinical decision support tool for primary palliative Care for Emergency Medicine (PRIM-ER). *BMC Med Inform Decis Mak*. 2020;20(1):13. doi:10.1186/s12911-020-1021-7

11. Hua MS, Ma X, Li G, Wunsch H. Derivation of data-driven triggers for palliative care consultation in critically ill patients. *J Crit Care*. 2018;46:79-83. doi:10.1016/j.jcrc.2018.04.014

12. Makar M, Ghassemi M, Cutler DM, Obermeyer Z. Short-term Mortality Prediction for Elderly Patients Using Medicare Claims Data. *Int J Mach Learn Comput*. 2015;5(3):192-197. doi:10.7763/IJMLC.2015.V5.506

13. Berg GD, Gurley VF. Development and validation of 15-month mortality prediction models: a retrospective observational comparison of machine-learning techniques in a national sample of Medicare recipients. *BMJ Open*. 2019;9(7):e022935. doi:10.1136/bmjopen-2018-022935

14. Cary MP, Zhuang F, Draelos RL, et al. Machine Learning Algorithms to Predict Mortality and Allocate Palliative Care for Older Patients With Hip Fracture. *J Am Med Dir Assoc*. 2021;22(2):291-296. doi:10.1016/j.jamda.2020.09.025

15. Reardon PM, Parimbelli E, Wilk S, et al. Incorporating Laboratory Values Into a Machine Learning Model Improves In-Hospital Mortality Predictions After Rapid Response Team Call. *Crit Care Explor*. 2019;1(7):e0023. doi:10.1097/CCE.0000000000000023

16. Deschepper M, Waegeman W, Vogelaers D, Eeckloo K. Using structured pathology data to predict hospital-wide mortality at admission. *PLOS ONE*. 2020;15(6):e0235117. doi:10.1371/journal.pone.0235117

17. Jain SS, Sarkar IN, Stey PC, Anand RS, Biron DR, Chen ES. Using Demographic Factors and Comorbidities to Develop a Predictive Model for ICU Mortality in Patients with Acute Exacerbation COPD. *AMIA Annu Symp Proc AMIA Symp*. 2018;2018:1319-1328.

18. Cao Y, Bass GA, Ahl R, et al. The statistical importance of P-POSSUM scores for predicting mortality after emergency laparotomy in geriatric patients. *BMC Med Inform Decis Mak*. 2020;20(1):86. doi:10.1186/s12911-020-1100-9

19. Wegier P, Koo E, Ansari S, et al. mHOMR: a feasibility study of an automated system for identifying inpatients having an elevated risk of 1-year mortality. *BMJ Qual Saf*. Published online June 28, 2019:bmjqs-2018-009285. doi:10.1136/bmjqs-2018-009285

20. Courtright KR, Chivers C, Becker M, et al. Electronic Health Record Mortality Prediction Model for Targeted Palliative Care Among Hospitalized Medical Patients: a Pilot Quasi-experimental Study. *J Gen Intern Med*. 2019;34(9):1841-1847. doi:10.1007/s11606-019-05169-2

21. Murphree DH, Wilson PM, Asai SW, et al. Improving the delivery of palliative care through predictive modeling and healthcare informatics. *J Am Med Inform Assoc*. 2021;(ocaa211). doi:10.1093/jamia/ocaa211

22. Major VJ, Aphinyanaphongs Y. Development, implementation, and prospective validation of a model to predict 60-day end-of-life in hospitalized adults upon admission at three sites. *BMC Med Inform Decis Mak*. 2020;20. doi:10.1186/s12911-020-01235-6

23. Manz CR, Chen J, Liu M, et al. Validation of a Machine Learning Algorithm to Predict 180-Day Mortality for Outpatients With Cancer. *JAMA Oncol*. Published online September 24, 2020. doi:10.1001/jamaoncol.2020.4331

24. Uneno Y, Taneishi K, Kanai M, et al. Development and validation of a set of six adaptable prognosis prediction (SAP) models based on time-series real-world big data analysis for patients with cancer receiving chemotherapy: A multicenter case crossover study. *PloS One*. 2017;12(8):e0183291. doi:10.1371/journal.pone.0183291

25. Avati A, Jung K, Harman S, Downing L, Ng A, Shah NH. Improving palliative care with deep learning. *BMC Med Inform Decis Mak*. 2018;18(Suppl 4):122. doi:10.1186/s12911-018-0677-8

26. Sahni N, Simon G, Arora R. Development and Validation of Machine Learning Models for Prediction of 1-Year Mortality Utilizing Electronic Medical Record Data Available at the End of Hospitalization in Multicondition Patients: a Proof-of-Concept Study. *J Gen Intern Med*. 2018;33(6):921-928. doi:10.1007/s11606-018-4316-y

27. Beeksma M, Verberne S, van den Bosch A, Das E, Hendrickx I, Groenewoud S. Predicting life expectancy with a long short-term memory recurrent neural network using electronic medical records. *BMC Med Inform Decis Mak*. 2019;19(1):36. doi:10.1186/s12911-019-0775-2

28. Wang L, Sha L, Lakin JR, et al. Development and Validation of a Deep Learning Algorithm for Mortality Prediction in Selecting Patients With Dementia for Earlier Palliative Care Interventions. *JAMA Netw Open*. 2019;2(7):e196972-e196972. doi:10.1001/jamanetworkopen.2019.6972

29. Lin Y-J, Chen R-J, Tang J-H, et al. Machine-Learning Monitoring System for Predicting Mortality Among Patients With Noncancer End-Stage Liver Disease: Retrospective Study. *JMIR Med Inform*. 2020;8(10):e24305. doi:10.2196/24305

30. Guo A, Foraker R, White P, Chivers C, Courtright K, Moore N. Using electronic health records and claims data to identify high-risk patients likely to benefit from palliative care. *Am J Manag Care*. 2021;27(1):e7-e15. doi:10.37765/ajmc.2021.88578

31. Blom MC, Ashfaq A, Sant'Anna A, Anderson PD, Lingman M. Training machine learning models to predict 30-day mortality in patients discharged from the emergency department: a retrospective, population-based registry study. *BMJ Open*. 2019;9(8):e028015. doi:10.1136/bmjopen-2018-028015

32. Adelson K, Lee DKK, Velji S, et al. Development of Imminent Mortality Predictor for Advanced Cancer (IMPAC), a Tool to Predict Short-Term Mortality in Hospitalized Patients With Advanced Cancer. *J Oncol Pract*. 2018;14(3):e168-e175. doi:10.1200/JOP.2017.023200

33. Parchure P, Joshi H, Dharmarajan K, et al. Development and validation of a machine learning-based prediction model for near-term in-hospital mortality among patients with COVID-19. *BMJ Support Palliat Care*. Published online September 22, 2020. doi:10.1136/bmjspcare-2020-002602

34. Escobar GJ, Ragins A, Scheirer P, Liu V, Robles J, Kipnis P. Nonelective Rehospitalizations and Postdischarge Mortality: Predictive Models Suitable for Use in Real Time. *Med Care*. 2015;53(11):916-923. doi:10.1097/MLR.0000000000000435

35. Gensheimer MF, Henry AS, Wood DJ, et al. Automated Survival Prediction in Metastatic Cancer Patients Using High-Dimensional Electronic Medical Record Data. *JNCI J Natl Cancer Inst*. 2018;111(6):568-574. doi:10.1093/jnci/djy178

36. Parikh RB, Manz C, Chivers C, et al. Machine Learning Approaches to Predict 6-Month Mortality Among Patients With Cancer. *JAMA Netw Open*. 2019;2(10). doi:10.1001/jamanetworkopen.2019.15997

37. Walraven C van. The Hospital-patient One-year Mortality Risk score accurately predicted long-term death risk in hospitalized patients. *J Clin Epidemiol*. 2014;67(9):1025-1034. doi:10.1016/j.jclinepi.2014.05.003

38. Barash Y, Soffer S, Grossman E, et al. Alerting on mortality among patients discharged from the emergency department: a machine learning model. *Postgrad Med J*. Published online December 3, 2020. doi:10.1136/postgradmedj-2020-138899

39. Leening MJG, Vedder MM, Witteman JCM, Pencina MJ, Steyerberg EW. Net reclassification improvement: computation, interpretation, and controversies: a literature review and clinician's guide. *Ann Intern Med*. 2014;160(2):122-131. doi:10.7326/M13-1522

40. Blanes-Selva V, Ruiz-García V, Tortajada S, Benedí J-M, Valdivieso B, García-Gómez JM. Design of 1-year mortality forecast at hospital admission: A machine learning approach. *Health Informatics J*. 2021;27(1):1460458220987580. doi:10.1177/1460458220987580

41. Portraits de la population. Accessed January 11, 2021. https://www.santeestrie.qc.ca/medias-publications/sante-publique/portraits-de-la-population/

42. Walraven C van, McAlister FA, Bakal JA, Hawken S, Donzé J. External validation of the Hospital-patient One-year Mortality Risk (HOMR) model for predicting death within 1 year after hospital admission. *CMAJ*. 2015;187(10):725-733. doi:10.1503/cmaj.150209

43. Groenwold RHH. Informative missingness in electronic health record systems: the curse of knowing. *Diagn Progn Res*. 2020;4(1):8. doi:10.1186/s41512-020-00077-0

44. Field CA, Welsh AH. Bootstrapping clustered data. *J R Stat Soc Ser B Stat Methodol*. 2007;69(3):369-390. doi:10.1111/j.1467-9868.2007.00593.x

45. Quan H, Li B, Couris CM, et al. Updating and Validating the Charlson Comorbidity Index and Score for Risk Adjustment in Hospital Discharge Abstracts Using Data From 6 Countries. *Am J Epidemiol*. 2011;173(6):676-682. doi:10.1093/aje/kwq433

46. Steyerberg EW. Restrictions on Candidate Predictors. *Clin Predict Models*. Published online 2019:191-206. doi:10.1007/978-3-030-16399-0_10

47. Quan H, Sundararajan V, Halfon P, et al. Coding Algorithms for Defining Comorbidities in ICD-9-CM and ICD-10 Administrative Data. *Med Care*. 2005;43(11):1130-1139.

48. Jameson JL, Fauci A, Kasper D, Hauser S, Longo D, Loscalzo J. *Harrison's Principles of Internal Medicine, Twentieth Edition*. 20th edition. McGraw-Hill Education / Medical; 2018.

49. Rajkomar A, Oren E, Chen K, et al. Scalable and accurate deep learning with electronic health records. *Npj Digit Med*. 2018;1(1):1-10. doi:10.1038/s41746-018-0029-1

50. Hastie T, Tibshirani R, Friedman J. Model Assessment and Selection. In: Hastie T, Tibshirani R, Friedman J, eds. *The Elements of Statistical Learning: Data Mining, Inference, and Prediction*. Springer Series in Statistics. Springer; 2009:219-259. doi:10.1007/978-0-387-84858-7_7

51. Lipton ZC. The Mythos of Model Interpretability. *ArXiv160603490 Cs Stat*. Published online June 10, 2016. Accessed August 26, 2019. http://arxiv.org/abs/1606.03490

52. Ahmad MA, Eckert C, Teredesai A, McKelvey G. Interpretable Machine Learning in Healthcare. *IEEE Intell Inform Bull*. 2018;19(1):7.

53. SIGDU - Système d'information de gestion des urgences - Actifs informationnels - Professionnels de la santé - MSSS. Accessed July 12, 2020. https://www.msss.gouv.qc.ca/professionnels/technologies-de-l-information/actifs-informationnels/sigdu/

54. MED-ECHO - Sources de données et métadonnées - Professionnels de la santé - MSSS. Accessed July 12, 2020. https://www.msss.gouv.qc.ca/professionnels/documentation-sources-de-donnees-et-indicateurs/sources-de-donnees-et-metadonnees/med-echo/

55. Breiman L. Random Forests. *Mach Learn*. 2001;45(1):5-32. doi:10.1023/A:1010933404324

56. Malley JD, Kruppa J, Dasgupta A, Malley KG, Ziegler A. Probability Machines: Consistent Probability Estimation Using Nonparametric Learning Machines. *Methods Inf Med*. 2012;51(1):74-81. doi:10.3414/ME00-01-0052

57. Hastie T, Tibshirani R, Friedman J. *The Elements of Statistical Learning: Data Mining, Inference, and Prediction, Second Edition*. 2 edition. Springer; 2009.

58. Assel M, Sjoberg DD, Vickers AJ. The Brier score does not evaluate the clinical utility of diagnostic tests or prediction models. *Diagn Progn Res*. 2017;1(1):19. doi:10.1186/s41512-017-0020-3

59. Major VJ, Jethani N, Aphinyanaphongs Y. Estimating real-world performance of a predictive model: a case-study in predicting mortality. *JAMIA Open*. 2020;3(2):243-251. doi:10.1093/jamiaopen/ooaa008

60. DiCiccio TJ, Efron B. Bootstrap confidence intervals. *Stat Sci*. 1996;11(3):189-228. doi:10.1214/ss/1032280214

61. Bernacki RE, Block SD, American College of Physicians High Value Care Task Force. Communication about serious illness care goals: a review and synthesis of best practices. *JAMA Intern Med*. 2014;174(12):1994-2003. doi:10.1001/jamainternmed.2014.5271

62. Allison TA, Sudore RL. Disregard of patients' preferences is a medical error: comment on "Failure to engage hospitalized elderly patients and their families in advance care planning." *JAMA Intern Med*. 2013;173(9):787. doi:10.1001/jamainternmed.2013.203

63. Hunink MGM, Weinstein MC, Wittenberg E, et al. *Decision Making in Health and Medicine: Integrating Evidence and Values*. 2nd ed. Cambridge University Press; 2014. doi:10.1017/CBO9781139506779

64. Kerr KF, Brown MD, Zhu K, Janes H. Assessing the Clinical Impact of Risk Prediction Models With Decision Curves: Guidance for Correct Interpretation and Appropriate Use. *J Clin Oncol Off J Am Soc Clin Oncol*. 2016;34(21):2534-2540. doi:10.1200/JCO.2015.65.5654

65. Vickers AJ, Calster BV, Steyerberg EW. Net benefit approaches to the evaluation of prediction models, molecular markers, and diagnostic tests. *BMJ*. 2016;352:i6. doi:10.1136/bmj.i6

66. Fitzgerald M, Saville BR, Lewis RJ. Decision curve analysis. *JAMA*. 2015;313(4):409-410. doi:10.1001/jama.2015.37

67. Localio AR, Goodman S. Beyond the Usual Prediction Accuracy Metrics: Reporting Results for Clinical Decision Making. *Ann Intern Med*. 2012;157(4):294-295. doi:10.7326/0003-4819-157-4-201208210-00014

68. Vickers AJ, Elkin EB. Decision curve analysis: a novel method for evaluating prediction models. *Med Decis Mak Int J Soc Med Decis Mak*. 2006;26(6):565-574. doi:10.1177/0272989X06295361

69. Van Calster B, Wynants L, Verbeek JFM, et al. Reporting and Interpreting Decision Curve Analysis: A Guide for Investigators. *Eur Urol*. 2018;74(6):796-804. doi:10.1016/j.eururo.2018.08.038

70. Vickers AJ, van Calster B, Steyerberg EW. A simple, step-by-step guide to interpreting decision curve analysis. *Diagn Progn Res*. 2019;3(1):18. doi:10.1186/s41512-019-0064-7

71. Wickham H, Averick M, Bryan J, et al. Welcome to the tidyverse. *J Open Source Softw*. 2019;4(43):1686. doi:10.21105/joss.01686

72. Wright MN, Ziegler A. ranger: A Fast Implementation of Random Forests for High Dimensional Data in C++ and R. *J Stat Softw*. 2017;77(1):1-17. doi:10.18637/jss.v077.i01

73. Eddelbuettel D, Francois R. Rcpp: Seamless R and C++ Integration. *J Stat Softw*. 2011;40(1):1-18. doi:10.18637/jss.v040.i08

74. Tingley D, Yamamoto T, Hirose K, Keele L, Imai K. mediation: R Package for Causal Mediation Analysis. *J Stat Softw*. 2014;59(1):1-38. doi:10.18637/jss.v059.i05

75. Wickham H. *Ggplot2: Elegant Graphics for Data Analysis*. 2nd ed. Springer International Publishing; 2016. doi:10.1007/978-3-319-24277-4

76. Claus Wilke, Spencer J Fox, Tim Bates, et al. *Wilkelab/Cowplot: 1.1.1*. Zenodo; 2021. doi:10.5281/zenodo.4411966

77. Gordon M, Lumley T. *Forestplot: Advanced Forest Plot Using "grid" Graphics*.; 2020. Accessed January 28, 2021. https://CRAN.R-project.org/package=forestplot
